# ‘Shaking the Ladder’ reveals how analytic choices can influence associations in nutrition epidemiology: beef intake and coronary heart disease as a case study

**DOI:** 10.1101/2023.12.05.23299578

**Authors:** Colby J. Vorland, Lauren E. O’Connor, Beate Henschel, Cuiqiong Huo, James M. Shikany, Carlos A. Serrano, Robert Henschel, Stephanie L. Dickinson, Keisuke Ejima, Aurelian Bidulescu, David B. Allison, Andrew W. Brown

## Abstract

**Background:** Many analytic decisions are made when analyzing an observational dataset, such as how to define an exposure or which covariates to include and how to configure them. Modelling the distribution of results for many analytic decisions may illuminate how instrumental decisions are on conclusions in nutrition epidemiology.

**Objective:** We explored how associations between self-reported dietary intake and a health outcome depend on different analytical decisions, using self-reported beef intake from a food frequency questionnaire and incident coronary heart disease as a case study.

**Design:** We used REasons for Geographic and Racial Differences in Stroke (REGARDS) data, and various selected covariates and their configurations from published literature to recapitulate common models used to assess associations between meat intake and health outcomes. We designed three model sets: in the first and second sets (self-reported beef intake modeled as continuous and quintile-defined, respectively), we randomly sampled 1,000,000 model specifications informed by choices used in the published literature, all sharing a consistent covariate base set. The third model set directly emulated existing covariate combinations.

**Results:** Few models (<1%) were statistically significant at p<0.05. More hazard ratio (HR) point estimates were >1 when beef was polychotomized via quintiles (95% of models) vs. continuous intake (79% of models). When covariates related to race or multivitamin use were included in models, HRs tended to be shifted towards the null with similar confidence interval widths compared to when they were not included. Models emulating existing published associations were all above HR of 1.

**Conclusions:** We quantitatively illustrated the impact that analytical decisions can have on HR distribution of nutrition-related exposure/outcome associations. For our case study, exposure configuration resulted in substantially different HR distributions, with inclusion or exclusion of some covariates being associated with higher or lower HRs.

This project was registered at OSF: https://doi.org/10.17605/OSF.IO/UE457

## Introduction

‘Shaking the Ladder’ is a phrase borrowed from Sam Savage who wrote: “The last thing you do before climbing on a ladder to paint the side of your house is to give it a good shake. By bombarding it with random physical forces, you simulate how stable the ladder will be when you climb on it. You can then adjust it accordingly so as to minimize the risk that it falls down with you on it.” (2) Following Savage’s analogy, just as we would shake a ladder to test its stability before trusting it, we must rigorously evaluate how our analytical choices influence our conclusions in nutritional epidemiology.

Investigating the associations of foods and nutrients with chronic disease endpoints is a challenging line of scientific inquiry. One of these challenges is the many reasonable decisions that investigators face when defining their exposure and outcome, and numerous analytical decisions such as how to configure the exposure, covariates, and model selections. For instance, covariates could be included or excluded, or defined as continuous, categorical, ordinal, or other ways (what we will refer to as covariate configuration). With each decision point, the combinations of defensible analytical decisions increase exponentially. Several studies demonstrated that if different sets of investigators were asked to analyze the same data set, their analysis approaches can vary substantially, sometimes resulting in vastly different conclusions (3–8).

Flexibility in analytical choices has been described as a garden of forking paths (9), or investigator degrees of freedom (10), among other names. Different analysis approaches may be responsible for some inconsistency in results in nutritional epidemiology research, although this has not been explored as extensively as other fields. Given that a set of decisions represents one of many reasonable potential approaches to analyzing the data, one analysis may lead to a conclusion that is represented by a minority of those approaches. Many foods and nutrients have both positive and negative associations with disease outcomes in the published literature (11), thus it is paramount to explore the degree to which this may be explained by analysis strategies.

One option to explore this phenomenon is to run many models with defensible analytic choices and report the distribution of results. This ‘multiverse’-style (12, 13) approach (similar to “specification curve analysis” (14), or “vibration of effects” (15)) can be used to explore the distribution of association estimates between an exposure and an outcome for many analytical paths, in turn allowing us to assess what influence the analytical decisions have on estimating the associations. The concept has been applied to several nutritional questions (15, 16) that focus on covariate inclusion and exclusion; however, additional choices such as the configuration of the nutritional exposure and covariates add additional flexibility. There is poor reporting in nutritional epidemiology for how covariate selection and configuration are decided (17), which raises questions about whether these methods are being systematically employed or if the selection and configuration processes are somewhat arbitrary and other choices also defensible.

Our objective was to evaluate to what degree associations between self-reported nutritional intake and health outcomes depend on different analytical decisions (e.g., exposure configuration, covariate inclusion and configuration, subject inclusion and exclusion criteria). Because covariate inclusion and configuration are not well reported in nutrition epidemiology, we aimed to evaluate the consequences of not carefully considering these. In contrast to previous approaches (e.g., specification curve, multiverse analysis), in which models are selected based on theory to explore the robustness of results for a particular research question, we randomly selected models based on existing published variable choices, and therefore the research questions represented by each model may change in subtle ways. We specifically use the case study of beef consumption and incident coronary heart disease (CHD). The beef-CHD relation is particularly appropriate for this approach because there is significant disagreement in the literature on the relationship between red meat intake and CHD (18–27); thus, analytical flexibility may be one explanation for this disagreement. Our analysis serves as a case study for how this approach can test the influence of analytical decisions on diet-outcome associations in nutrition specifically, and in observational association studies generally.

## Subjects and Methods

### Study Sample

We used data from the REasons for Geographic and Racial Differences in Stroke (REGARDS) prospective cohort (28). REGARDS is a national, longitudinal cohort of 30,239 Black and White women and men ages 45 and older, who were recruited from 2003-2007. After excluding 56 participants with data anomalies, we utilized data from 30,183 participants. Participants’ CHD status was last updated in 2018. Participants with a history of CHD or cancer at baseline were excluded from our analyses. **Figure 1** and **Supplemental Table 2** describe additional participant exclusions, such as those based on self-reported energy intake cutoffs (varying methods to exclude extreme/implausible data).

**Figure 1.**
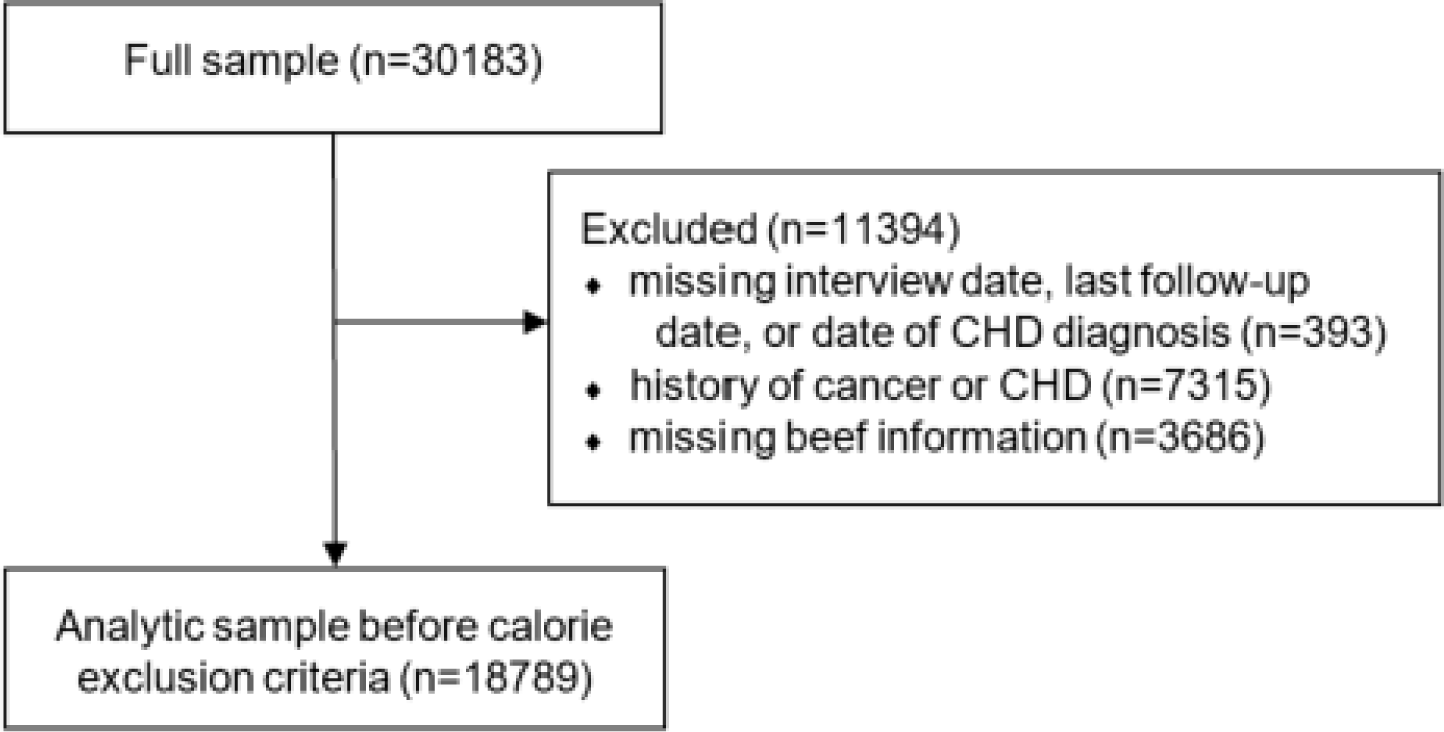
Sample size flow chart. The number of participants in the full sample, number after participant exclusions and before energy intake exclusions, and reasons for exclusion.

### Exposure, Outcome, and Covariate Selection Process

Self-reported beef consumption was originally estimated via the Block 98 food frequency questionnaire (FFQ). We defined beef intake using gram weight estimates (using the variables ‘hamburger’, ‘beefroast’ ‘beeffattrimmed’, ‘beeffatnotrim’ based on the FFQ items “hamburgers, cheeseburgers, meat loaf, at home or in a restaurant” and “beef steaks”). Values in the hamburger variable were multiplied by a proportion of 0.59 to refine the estimation of beef content. This proportion was determined from the Food and Nutrient Database for Dietary Studies 2017-2018 data (29). The outcome of incident CHD was defined as myocardial infarction event or acute CHD death.

Our inclusion of covariates and their configurations was informed by prior literature, allowing us to indirectly crowdsource expert choices in covariate inclusion and configuration that had also passed peer review. The prior analyses were identified from 1) a previous systematic review of prospective cohort studies of red meat and CVD outcomes (30), 2) a selection of observational studies assessing red meat or beef and CVD outcomes known to coauthors or identified through literature searching, and 3) previous analyses using the REGARDS dataset by a coauthor (JS). All references are listed in documents attached to our preregistration: https://doi.org/10.17605/OSF.IO/UE457. Covariates and their configurations identified from studies not using the REGARDS data were matched as closely as possible to REGARDS variables. History of chronic obstructive pulmonary disease and sleep outcomes in the literature sampling did not have a close match within the REGARDS dataset and were not included in models. Configurations included categorical, continuous, or ordinal via quintiles or sex-specific median. A complete list of included variables, their configurations, as well as their corresponding REGARDS variable names, and variables unable to be matched to REGARDS variables, is available in the following repository: https://doi.org/10.17605/OSF.IO/SY96K.

Three sets of models were developed. First, we created a random sample of 1,000,000 model combinations, based on variables that appear in previous literature, where self-reported beef intake was defined as either continuous (model set 1) or in quintile defined categories (model set 2). Then, we emulated prior literature to try to reproduce existing variable choice combinations exactly as they have been previously published as specific, expert, pre-specified analytical examples in the REGARDS dataset (model set 3); these models were reproduced in the REGARDS set (i.e., they were not randomly sampled). In the first and second model set, age, sex, energy intake, size of census tract, and REGARDS region were included in all models, consistent with the prior literature; thus, we decided it would be unreasonable that expert analysts would define a model without them. For food- and nutrient-related variables, their inclusion was randomly varied in models, but when included we used the same configuration for all such included variables because we did not believe analysts would consider models reasonable if configurations differed among these variables. For other variables, we randomly varied their inclusion and configuration. Direct comparisons between hazard ratios (HRs) derived from continuous versus quantile exposure definitions can be difficult; therefore, we decided to express continuous beef intake per 50g unit increase, which was comparable to the difference in mean reported intakes in the highest 20% versus lowest 20% of participants (50.01g). Covariate inclusion and configurations for these models are described in the ‘statistical analysis and visualization’ section. **Table 1** summarizes which variables were kept constant in all models, and which were varied.

**Table 1.** Model sets.

|  | Model sets |  |  |
| --- | --- | --- | --- |
| Variable choices | Model set 1 | Model set 2 | Model set 3 |
| Outcome | CHD |  |  |
| Exclusion criteria | History of CHD or cancer |  |  |
| Base model <sup>1</sup> | 'Basic' |  |  |
| Measure of association | HR |  |  |
| Exposure definition | Beef intake <sup>2</sup> |  |  |
| Exposure configuration | Continuous | Quintiles | Quintiles and Continuous |
| Energy cutoff | Vary cutoffs per **Supplemental Table 2** |  | According to published papers <sup>3</sup> |
| Covariates | Inclusion/Exclusion and configuration when included |  | According to published papers <sup>3</sup> |

Grey = Consistent across model sets; White = Varies across model sets for the exposure configuration; Blue = Varies within each model set. <sup>1</sup> 'Base model' covariates that were always included were: age (3 different configurations, one at a time), gender, calorie intake (3 different configurations, one at a time), size of census tract, and REGARDS region. <sup>2</sup> Beef was defined using gram weight estimates (using the following variables: 'hamburger', 'beefroast' 'beeffattrimmed', 'beeffatnotrim'; based on the FFQ items "hamburgers, cheeseburgers, meat loaf, at home or in a restaurant" and "beef steaks"); values in the hamburger variable were multiplied by a proportion of 0.59 to refine the estimation of beef content. This proportion was determined from the Food and Nutrient Database for Dietary Studies 2017-2018 data (29). <sup>3</sup> We used the energy cutoff that aligned closest to one of our 7 different configurations for energy cutoffs. <sup>3</sup> We used covariate configurations that aligned closest with one of our covariate configurations.

### Statistical Analysis and Visualization

Cox proportional hazards regression models were used, with time from enrollment as the underlying time metric within each of the analyses, censoring date of CHD diagnosis, date of death, date of withdrawal, or date of last follow-up. Sample size was allowed to vary on a complete-case basis depending on which covariates were included in each model. Missing data were not imputed. This approach was consistent with the sampled prior literature. The proportion of missingness for any given covariate is shown in **Supplemental Table 3**. The total number of possible combinations of covariates and configurations was far beyond computational capabilities (see results); thus, model sets 1 and 2 randomly sampled 1,000,000 variable combinations total (500,436 for beef as continuous and 499,564 for beef as quintiles). Covariates were first sampled for inclusion or exclusion; if the covariate was included, the configurations were equally sampled. For example, in the case of a food variable such as dairy intake, it had a 50% chance of being included; if it was included, then each of the three configurations (continuous, sex-specific median or quintile) were sampled with equal probability (1 out of 3 conditional on being included). For model set 3, all models were computed as closely as possible to emulate the prior literature (see https://osf.io/sy96k for the models).

Code to run the analysis was developed and tested on a small scale and later parallelized for the full 1,000,000 model runs. Briefly, code consisted of a ‘for loop’ iterating through model runs and saving the model output. After a loop dependency analysis, we found that the loop did not depend on any other “outside” data including dependencies between models, so we parallelized by modifying the code to run small subsets of the total models (500 models) and run this code multiple times so that these subsets were run in parallel. Lastly, all subsets of results were combined using a Linux shell script. Parallel code was run on Carbonate, which is Indiana University’s large-memory computer cluster, designed to support data-intensive computing (31).

Model results were visualized and further analyzed in different ways. Distribution of HRs, z-scores and p-values were plotted in histograms. To visualize the impact of variable configuration on beef hazard ratios we created specification curve plots. In the first step, we did this for the beef variable itself, but then also for all other covariates included in the model. Specification curve plots show the distribution of the estimates of the association of interest and the impact of analytic decisions on those estimates by showing the distribution of estimates for each analytic decision. Bivariate scatterplots of the beef HRs and its 95% confidence interval (CI) widths were created to show possible relationships of variable inclusion/exclusion on estimates and their precision. Additionally, to show data density, we plotted bivariate KDE curves. Model meta-information such as sample size, number of covariates, and number of CHD events were plotted showing their density by significance of the beef HR. Descriptive statistics of the beef HR, its z-scores, its 95% CI, and its p-values were calculated overall and by beef configuration for model set 1 and 2 and the models from the literature. Impact of covariate configuration (including exclusion) on the beef HR was assessed in multivariate logistic regression models that adjusted for all included covariates and their configurations at the same time. Odds ratios and 95% CI are presented. Lastly, a series of two-sample Kolmogorov-Smirnov statistics (e.g., D statistics) were calculated to quantify the distance between the cumulative distributions of HR by inclusion/exclusion of covariates. Significance tests with p-values were not used for the K-S test because the samples are not independent and identically distributed. Cross-correlation coefficient and likeness measures between KDEs were calculated as defined in (32). Higher values for both indicate higher overlap between both KDEs. For all covariate specific plots, we show the results for four selected covariates in the main text; the remaining plots for all covariates can be found in supplemental materials.

SAS [version 9.4] was used to prepare the dataset for analysis, R [version 4.1.1] was used on a x86 64-bit Linux cluster for the models, and R [version 4.2.3] and RStudio [version 2023.03.0] were used for analyses and to produce visualizations.

### Ethics

This study was approved by the Indiana University Institutional Review Board (#11227). The REGARDS study was previously approved by all associated institutional review boards (33).

### Power calculation

We estimated that we would have sufficient power to detect most associations with HR > 1.1 with a sample size of 20,000 or lower (**Supplemental Figure 1**) using incident CVD from the lowest quartile of estimated red meat consumption from Zhong et al. (24) as the reference hazard. Thus, REGARDS provided a sufficiently large sample for small HRs.

### Inference Criteria

We used p < 0.05 as a threshold of statistical significance within any given analysis, consistent with standard practice in the prior nutritional epidemiology literature. We did not correct for multiple comparisons, because each analysis represents one theoretical independent choice of many that an analyst could make.

### Changes after Preregistration

Our project was preregistered at OSF: https://doi.org/10.17605/OSF.IO/UE457. We describe changes post-registration and our reasoning in **Supplemental Table 1**.

## Results

### Calculation of Model Possibilities

For model sets 1 and 2, with 2 beef configurations, 7 exclusion criteria configurations based on self-reported energy intake cutoffs (**Supplemental Table 2**), and 34 covariates with 117 configurations, we calculated over 4.16 quadrillion total possible model combinations. Running all possible combinations would not have been feasible even with the use of parallel high-performance computing resources available to the study team. Additionally, storing, analyzing, and presenting model results would have been challenging if not impossible.

Comparing results (e.g., distribution of HRs and covariate associations) from an initial test run using 10,000 models and the results shown herein of 1,000,000 models, we are not convinced that more insights will be gleaned from an even greater number of samples.

### Model Summaries

**Supplemental Table 3** shows covariates and their configurations as defined using REGARDS data, along with the number of missing values for each. **Table 2** shows the mean, median, 5^th^ to 95^th^ percentile range, and min and max values for HRs, p-values and z-scores in model sets 1 and 2. **Figure 2** shows distributions of HRs, significant HRs (with p<0.05), z-scores, and p-values by beef configuration.

**Figure 2.**
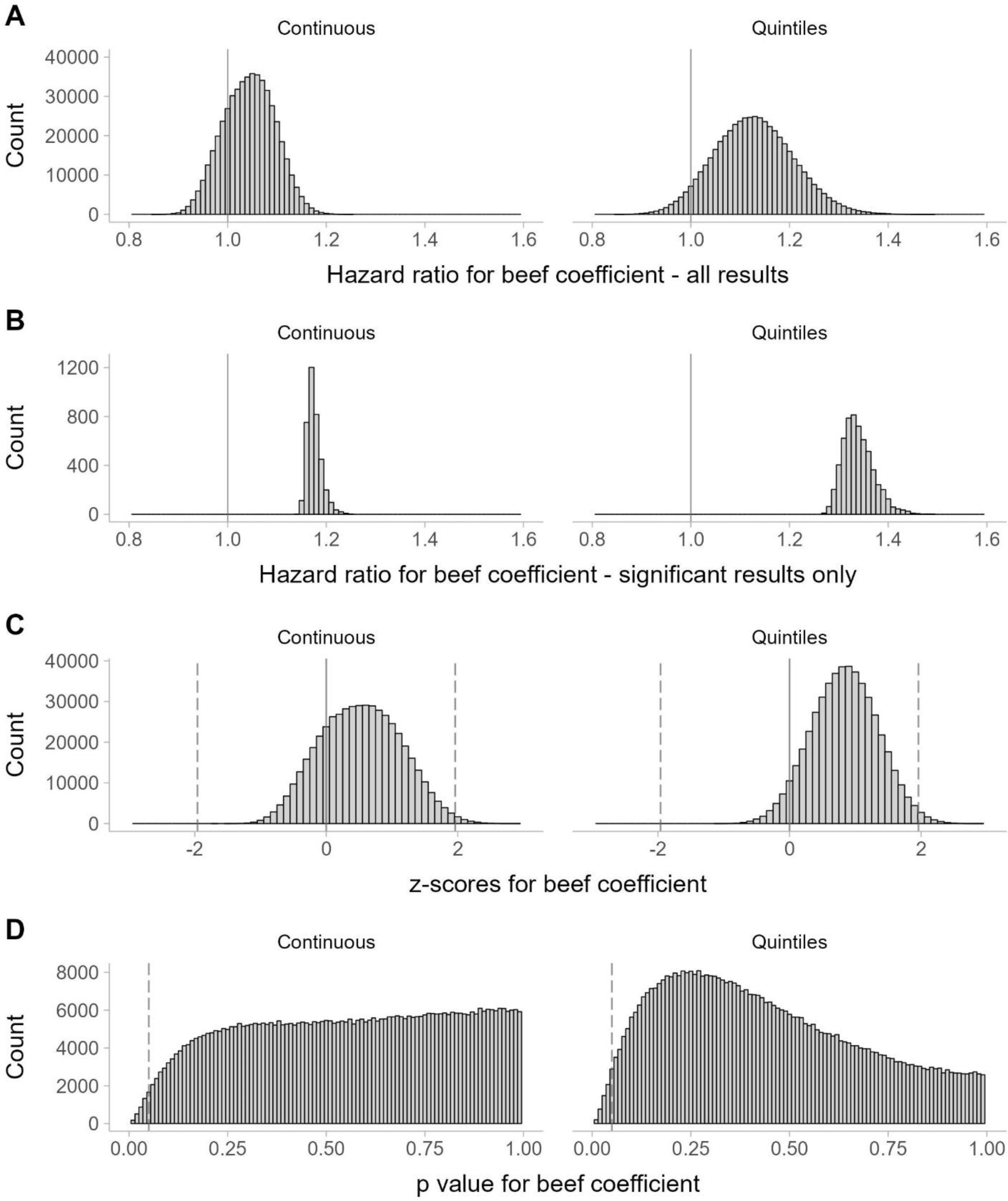
Frequency of Hazard ratios all model results (row A), Hazard ratios from significant model results (p<0.05, row B), z scores from all model results (row C), and p-values from all model results (row D) for coefficient for beef intake for model set 1 when beef is expressed as continuous (left; per 50g), and model set 2 when beef is expressed as quintiles (right; highest vs. lowest). y-axis shows the number of models, the scale in the rows B and D is smaller to better show the distribution for significant hazard ratios and p-values. Vertical dashed lines represent z=|1.96|, values outside the ±1.96 range are considered significant at p<0.05 (row C), p=0.05 (row D).

**Table 2.**
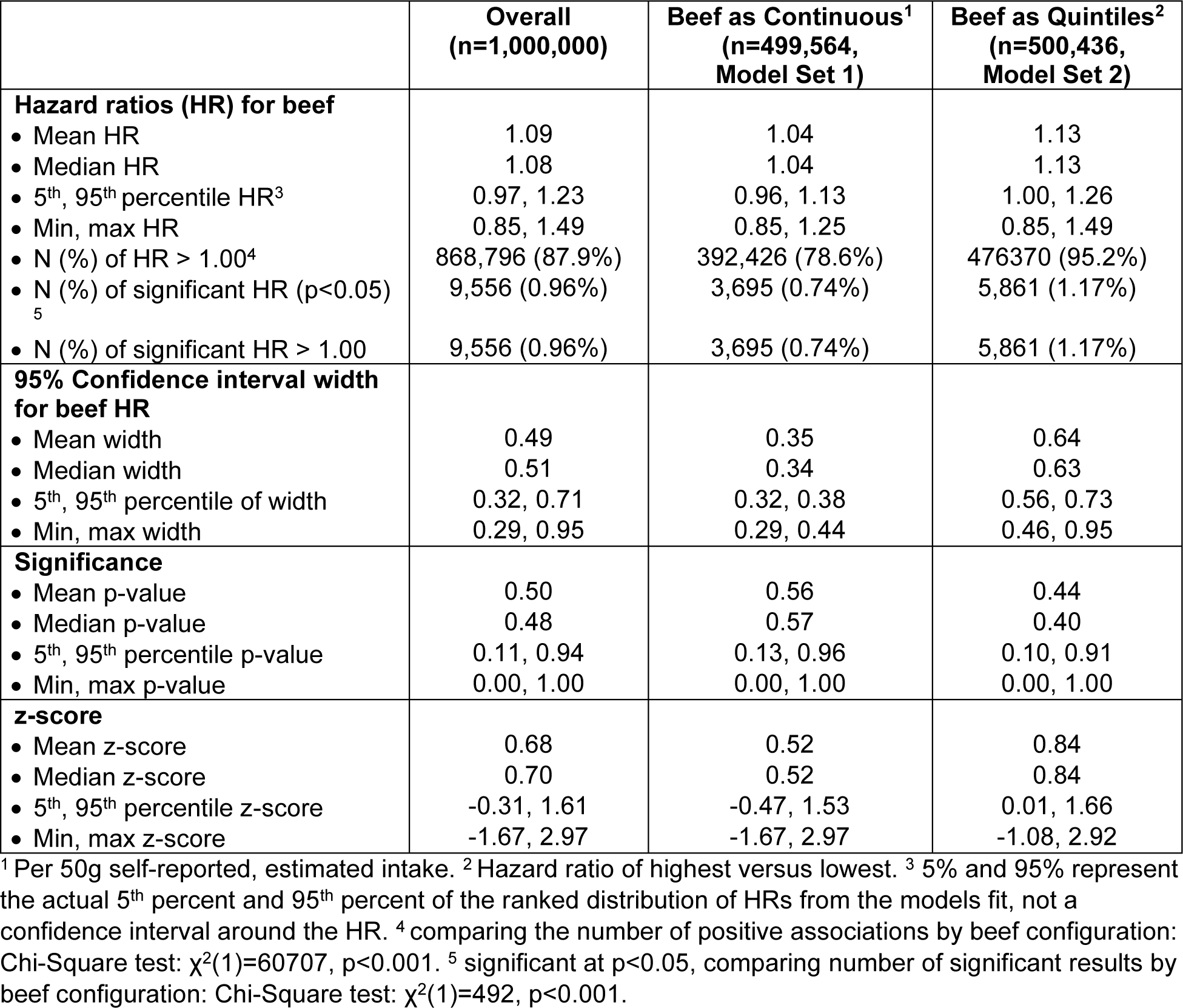
Descriptive statistics for model sets 1 and 2.

|  | **Overall**<br>**(n=1,000,000)** | **Beef as Continuous<sup>1</sup>**<br>**(n=499,564,**<br>**Model Set 1)** | **Beef as Quintiles<sup>2</sup>**<br>**(n=500,436,**<br>**Model Set 2)** |
| --- | --- | --- | --- |
| **Hazard ratios (HR) for beef** |  |  |  |
| • Mean HR | 1.09 | 1.04 | 1.13 |
| • Median HR | 1.08 | 1.04 | 1.13 |
| • 5<sup>th</sup>, 95<sup>th</sup> percentile HR<sup>3</sup> | 0.97, 1.23 | 0.96, 1.13 | 1.00, 1.26 |
| • Min, max HR | 0.85, 1.49 | 0.85, 1.25 | 0.85, 1.49 |
| • N (%) of HR > 1.00<sup>4</sup> | 868,796 (87.9%) | 392,426 (78.6%) | 476,370 (95.2%) |
| • N (%) of significant HR (p<0.05)<br><sup>5</sup> | 9,556 (0.96%) | 3,695 (0.74%) | 5,861 (1.17%) |
| • N (%) of significant HR > 1.00 | 9,556 (0.96%) | 3,695 (0.74%) | 5,861 (1.17%) |
| **95% Confidence interval width**<br>**for beef HR** |  |  |  |
| • Mean width | 0.49 | 0.35 | 0.64 |
| • Median width | 0.51 | 0.34 | 0.63 |
| • 5<sup>th</sup>, 95<sup>th</sup> percentile of width | 0.32, 0.71 | 0.32, 0.38 | 0.56, 0.73 |
| • Min, max width | 0.29, 0.95 | 0.29, 0.44 | 0.46, 0.95 |
| **Significance** |  |  |  |
| • Mean p-value | 0.50 | 0.56 | 0.44 |
| • Median p-value | 0.48 | 0.57 | 0.40 |
| • 5<sup>th</sup>, 95<sup>th</sup> percentile p-value | 0.11, 0.94 | 0.13, 0.96 | 0.10, 0.91 |
| • Min, max p-value | 0.00, 1.00 | 0.00, 1.00 | 0.00, 1.00 |
| **z-score** |  |  |  |
| • Mean z-score | 0.68 | 0.52 | 0.84 |
| • Median z-score | 0.70 | 0.52 | 0.84 |
| • 5<sup>th</sup>, 95<sup>th</sup> percentile z-score | -0.31, 1.61 | -0.47, 1.53 | 0.01, 1.66 |
| • Min, max z-score | -1.67, 2.97 | -1.67, 2.97 | -1.08, 2.92 |

<sup>1</sup> Per 50g self-reported, estimated intake. <sup>2</sup> Hazard ratio of highest versus lowest. <sup>3</sup> 5% and 95% represent the actual 5<sup>th</sup> percent and 95<sup>th</sup> percent of the ranked distribution of HRs from the models fit, not a confidence interval around the HR. <sup>4</sup> comparing the number of positive associations by beef configuration: Chi-Square test: $\chi^2(1)=60707$ , $p<0.001$ . <sup>5</sup> significant at $p<0.05$ , comparing number of significant results by beef configuration: Chi-Square test: $\chi^2(1)=492$ , $p<0.001$ .

As shown in **Figure 2** (top panel) and **Table 2**, the proportion of models with HR greater than 1.0 was much higher when beef intake was expressed in quintile defined categories (right, 95.2%) compared to expressed as a continuous variable (per 50g intake; left, 78.6%). Of the 9556 significant beef HRs, only 38.7% (3695) came from models using beef as a continuous variable while the other 61.3% (5861) came from the quintile models. This is further illustrated in the specification curve (**Figure 3**) in which HRs are ranked from lowest to highest; the vertical dashed line (in top plot) shows that 131,205 of 1,000,000 models were less than an HR of 1. The bottom plot of **Figure 3** shows 1) the distributions of the ranked HR with the associated exposure configurations: continuous or quintile beef intake, and 2) that the statistically significant (p < 0.05) HRs appear lower along the ranking for continuous beef intake compared to beef intake in quintile defined categories. Despite these differences, overall, very few models produced statistically significant associations in either approach (9556/1000000 models=0.96% of all models), and these significant associations were associated with higher HRs (all were above 1.0).

**Figure 3.**
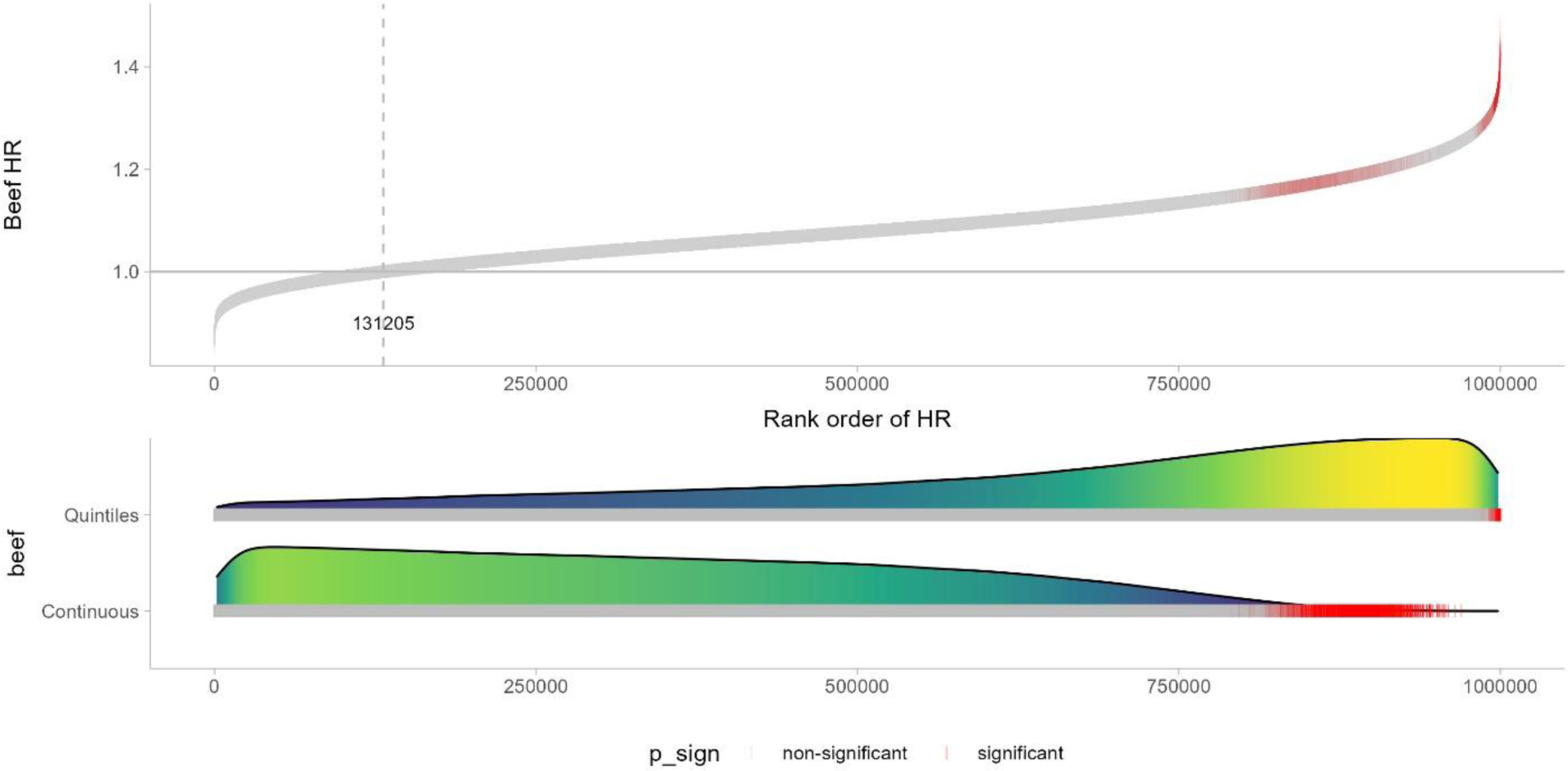
Specification curve that shows the distribution of HRs for the association between self-reported beef consumption and CHD for model sets 1 and 2. Each of the 1,000,000 model combinations is represented by a thin vertical bar in either gray color (if p=>0.05) or red (if p<0.05). Curves in the bottom plot show distribution of HR by beef intake configuration, with the same gray or red thin vertical bars as above. A bar in the top plot can be traced down to the bottom plot. Color represents the density in models along the HR distribution (yellow=more models, dark blue=fewer models).

### Influence of Covariates on HRs and Precision

We used the same HRs that were plotted in Figures 2 and 3 to generate additional plots to highlight the influence of the covariate selection and configuration. **Figure 4** shows the ranked HRs for continuous beef (left) and beef in quintile defined categories (right) by four selected covariates (race, income, education, and multivitamin use). We selected these variables as examples to highlight because their inclusion/exclusion and configuration showed a range of strong to weak influences on HRs. HRs that came from model specifications that adjusted for race or years of multivitamin use tended towards smaller HRs while HRs from models not adjusting for race tended towards higher HRs. Note that these shifts reflect the impact on the beef coefficient with inclusion or configuration of the covariate, not the coefficient for the covariate itself (e.g., the influence of a specific race or multivitamin use on the HR). For income, we observed that either not adjusting for income or adjusting for income using four categories appears to have not much impact on the size of the HR but adjusting for income as a continuous measure tended towards higher HRs at the right tail. Lastly, the covariate education is an example where the distributions of beef HRs do not appear to differ much when adjusting or not adjusting in the analysis.

**Figure 4.**
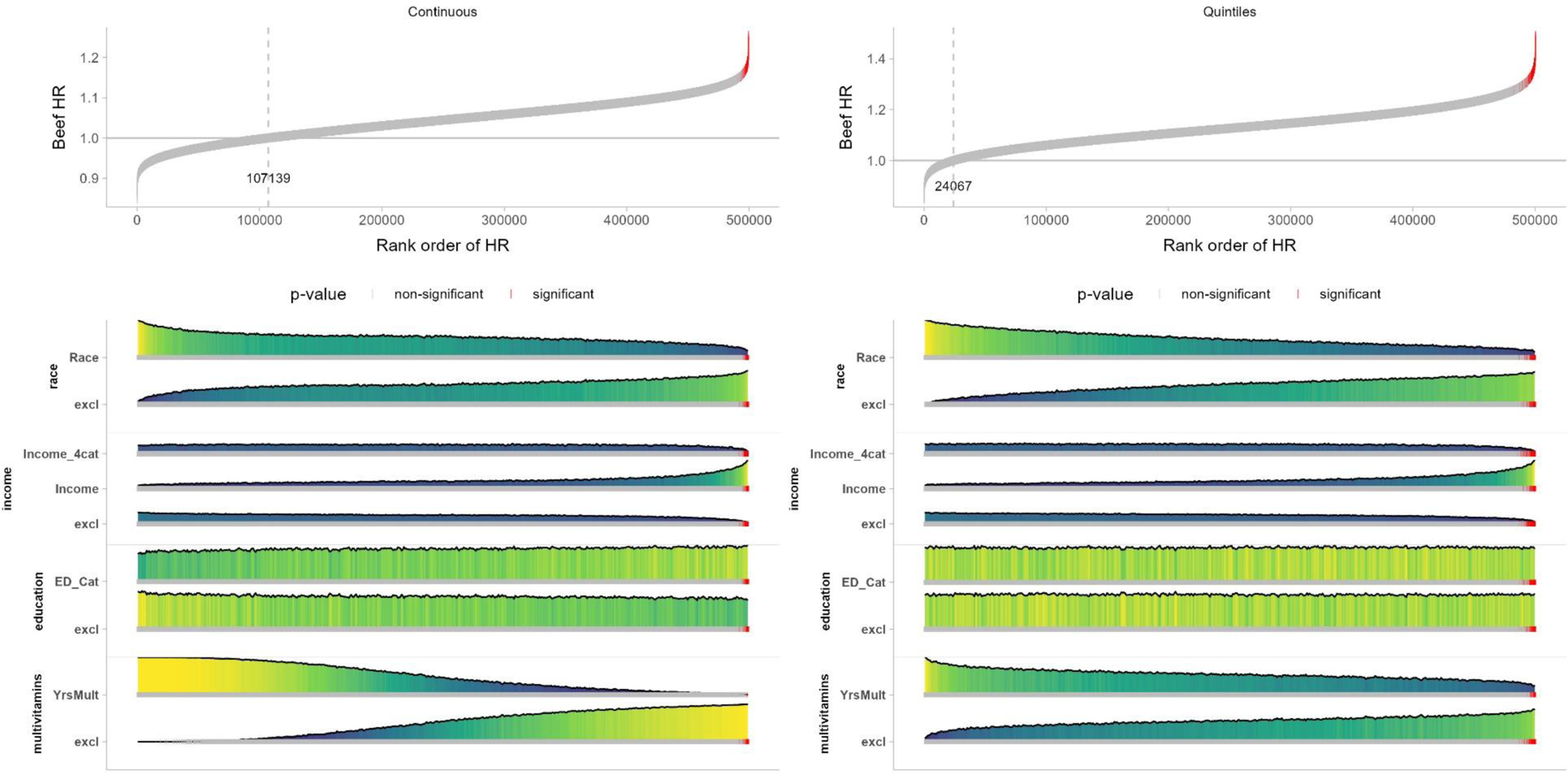
Specification curves that show the distribution of HRs for the association between self-reported beef consumption and CHD, when beef is expressed as a continuous variable (left), and as quintiles of intake (right). Each combination of covariates is represented if a vertical line were traced from any point on the curve on top down through each variable underneath. Red lines represent models that were p < 0.05. Variables: race (White; Black); income (‘Income_4cat’: <$20K, $20K-$35K, $35K-75K, $75K+, Refused; ‘Income’: 1 (<5K), 2 (5-10K), 3 (10-15K), 4 (15-20K), 5 (20-25K), 6 (25-35K), 7 (35-50K), 8 (50-75K), 9 (75-150K), 10 (>150K)); education (‘ED_Cat’: <HS, HS, Some College, College+); multivitamins (‘YrsMult’: Years took multivitamins (0=No vitamins taken in past year, 1=Less than 1 year, 2=1 Year, 3=2 Years, 4=3-4 Years, 5=5-9 Years, 6=10+ Years)). Color represents the density in models along the HR distribution (yellow=more models, dark blue=fewer models).

We explored the results for these four covariates further in **Figure 5** by plotting HRs across the x-axis and the 95% confidence interval width of the HRs across the y-axis, the latter representing precision of the estimate, for the same set of covariates. When multivitamin use (bottom right) was excluded from the beef as continuous models, HRs and confidence interval width tended to be shifted above 1 compared to inclusion, but with similar precision, whereas inclusion (i.e., adjusting for years of multivitamin use) tended to be centered around 1. The trends were similar in the quintile approach, though inclusion or exclusion models remained with densities higher than an HR of 1 and less precision (i.e., wider confidence intervals) as compared to the continuous beef models. For race (top left), the results are much more diffuse for inclusion and exclusion for both continuous and quintile beef models. For income (top right), the density curves for exclusion and categorical income adjustment (labeled ‘Income_4cat’) are almost identical and on top of each other, while the curve for continuous income adjustment (labeled ‘Income’) is shifted away from the null with less precision. Lastly, for education, the density curves for inclusion vs exclusion do not show visual differences. The plots for all 34 covariates are shown in **Supplemental File 1**. In order to analytically assess differences in the distributions of HRs by inclusion/exclusion of covariates, we ran a series of Kolmogorov-Smirnov tests (**Table 4**). With the high number of different models we ran, we had large sample sizes, so these tests had high power to detect even minor differences. All beef hazard ratio distributions were significantly different when including a covariate compared to excluding a covariate. When using a Bonferroni-corrected p-value of 0.0017 (0.05/29 tests), inclusion vs exclusion of two covariates (“grains” and “monofat”) was no longer significantly associated with the HR distributions in the continuous beef configuration, while all other tests remained significant. Visually inspecting the distributions of beef HRs by configurations of covariates, we observed the largest shifts for the following covariates in the beef as quintile models: race, income, multivitamin use, history of diabetes, history of stroke, physical activity, fiber, and fruit intake (**Supplemental File 3**).

**Figure 5.**
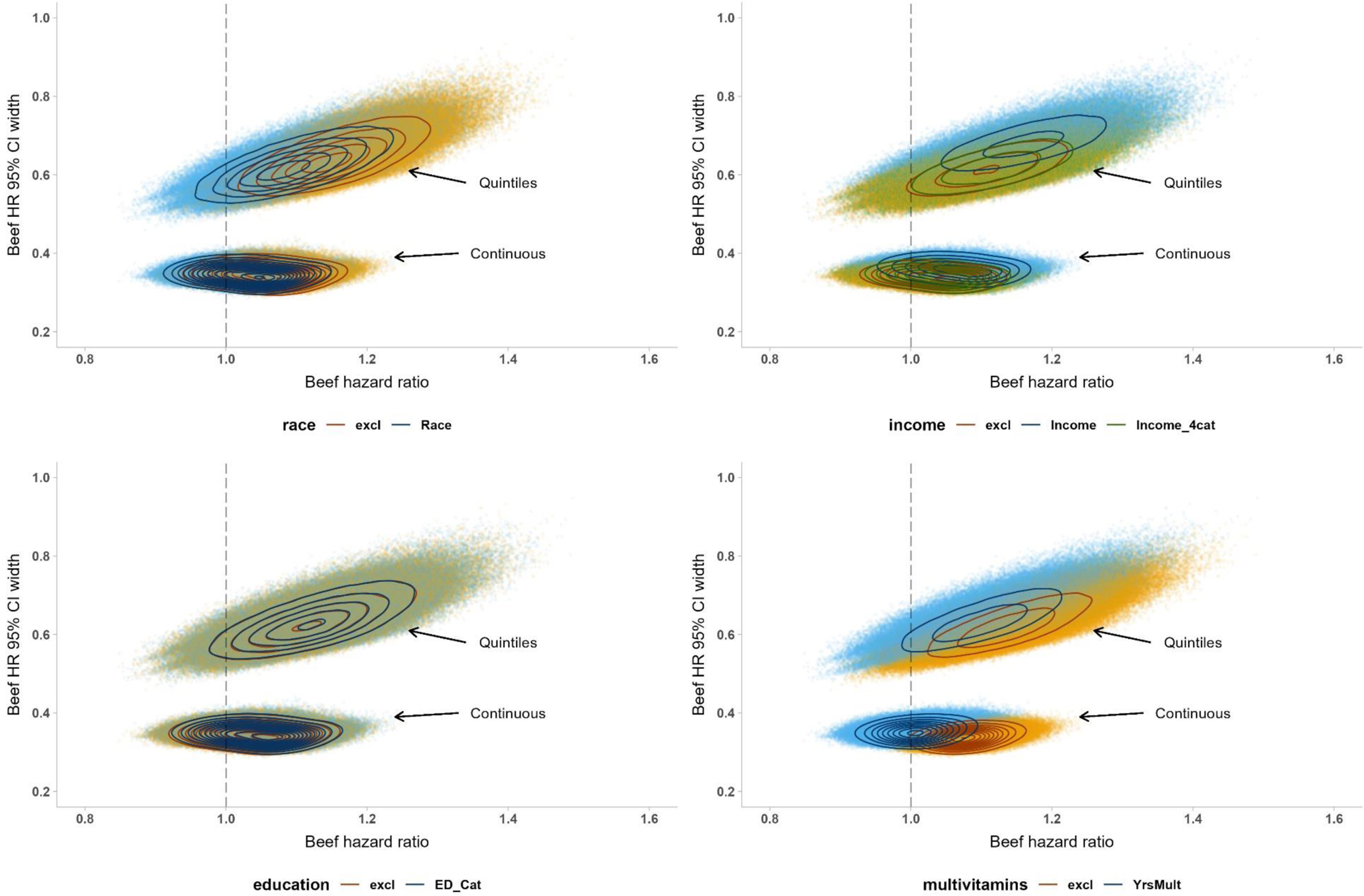
Plots that show the distribution of HRs vs. 95% CI width for four selected covariates (top left: race; top right: income, bottom left: education, bottom right: years of multivitamin use) when inclusion/exclusion and configuration is varied, when self-reported beef consumption is expressed as continuous and as quintiles of intake. Lines are contour lines from a kernel density estimation using a normal distribution kernel; kernel smoothing was done over 200 grid points. Kernel density estimates were made using the MASS package with the kde2d function, as described (50).

Although overall only about 1% of the models resulted in statistically significant HRs, we tested the influence that inclusion and configuration of covariates had on the statistical significance of the HRs using separate multivariable logistic models for the two beef configurations. **Supplemental Table 4** displays the results showing the proportion of significant HRs for each configuration, an odds ratio (OR) for significant HRs using one of the configurations as the reference group (in most cases: covariate exclusion), and the p-value for the OR. We observed significant associations for all covariates except education (type 3 p-value = 0.061) and history of PAD (p = 0.129), indicating that all other covariates had some influence whether the association between self-reported beef and CHD was statistically significant. Notably high ORs were found for the continuous configuration of income (OR=85.42 (95% CI: 73.05, 99.88) for continuous beef, OR=28.91 (95% CI: 26.04, 32.09) for quintile beef intake), meaning that when continuous income was included in models, there were higher odds of a significant HR for the beef-CHD association. In contrast, including the years of multivitamin use in the model resulted in far fewer significant HRs for beef (OR<0.01 (95% CI: <0.01, <0.01) for continuous beef, OR=0.02 (95% CI: 0.02, 0.03) for quintile beef) compared to excluding multivitamin use.

Figure 6 shows pairwise density plots for the number of CHD events and sample size depending on the number of covariates in the model. Models with significant beef HR (p<0.05) show density curves in red and those not significant (p>=0.05) in black. There was a tendency that as more covariates were included in the model, the sample size was smaller. Given that we let the sample size vary depending on the complete case of the model specification, this result is expected. With higher sample sizes, the number of CHD events tended to be higher (Figure 6b), and models with significant HRs appear to have a higher number of CHD events (Figure 6b). Finally, models with significant HRs tended to have a lower number of covariates (Figure 6c). Specification curves showing the distribution of HRs are shown for all 32 covariates in **Supplemental File 3** (beef as continuous) and **Supplemental File 4** (beef as quintiles).

**Figure 6:**
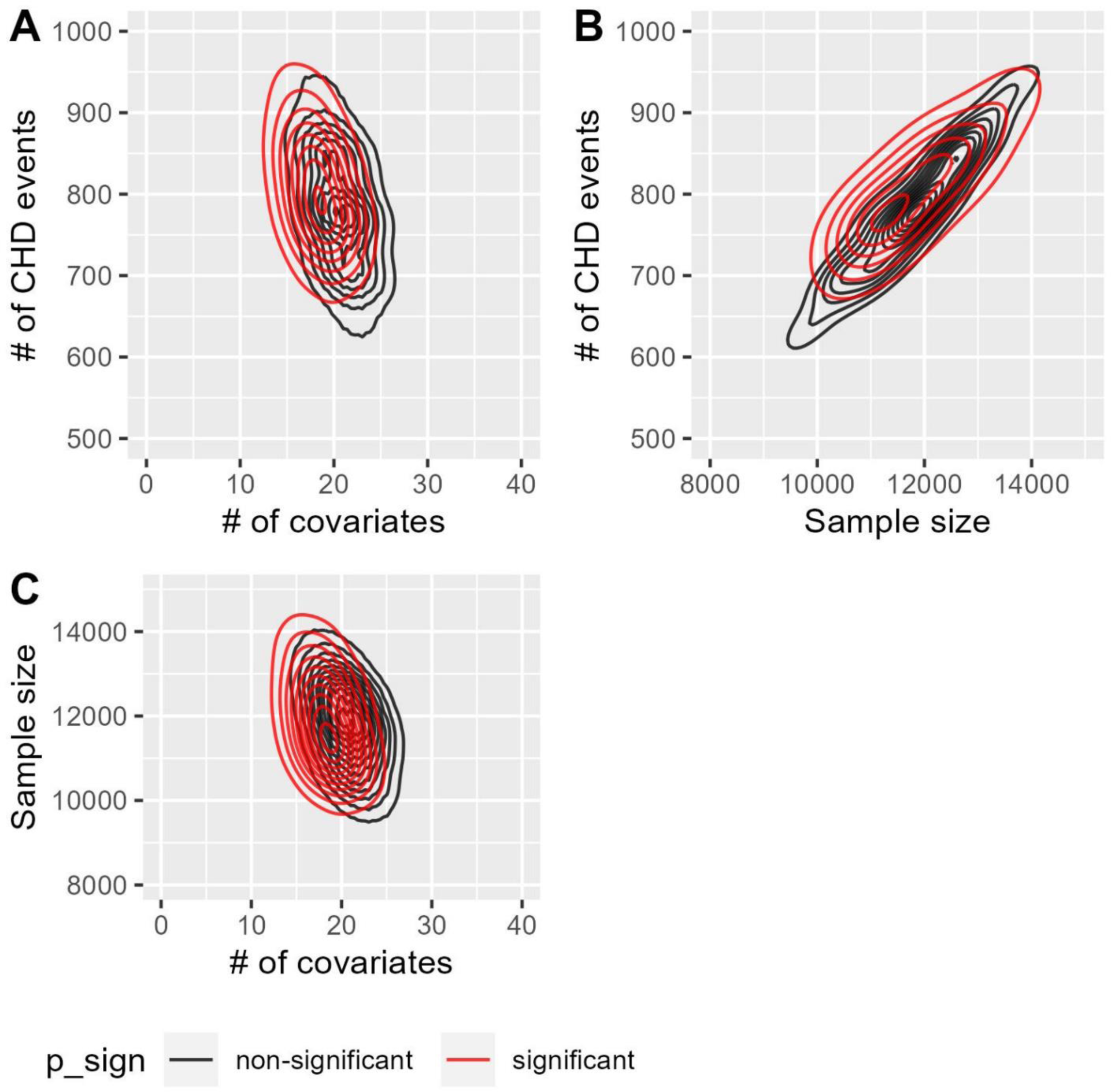
Pairwise density plots of number of CHD events (A, B), sample size (B, C), and number of covariates in the model (A, C). Density curves in red color represent models with significant coefficients for beef (p<0.05).

### Emulating existing literature

To benchmark our agnostic, random sampling approach against expert-chosen models, we reproduced 20 models from the literature (see references in the preregistration). Figure 7 shows that the frequency of HRs for these models were all greater than 1.0, and, from visual inspection, tended to have higher HRs and lower precision when beef was expressed as quintiles of intake. Two of 20 models were statistically significant; both of them when beef was expressed as quintiles of intake (**Table 3**). Figure 8 shows the cumulative distributions of the HR from the 1,000,000 models from model sets 1 and 2 combined, and the 20 models that we emulated as they appear in the existing literature. We see that the empirical cumulative distribution function (ECDF) for the existing literature is below the one for model sets 1 and 2, which suggests that the results from the existing literature are shifted towards higher HRs overall. The strongest divergence between the two distributions (D=0.438) can be observed for HRs below the median (ECDF=0.50), with a higher concentration of HR between 1.05 and 1.10 for the results from existing literature.

**Figure 7.**
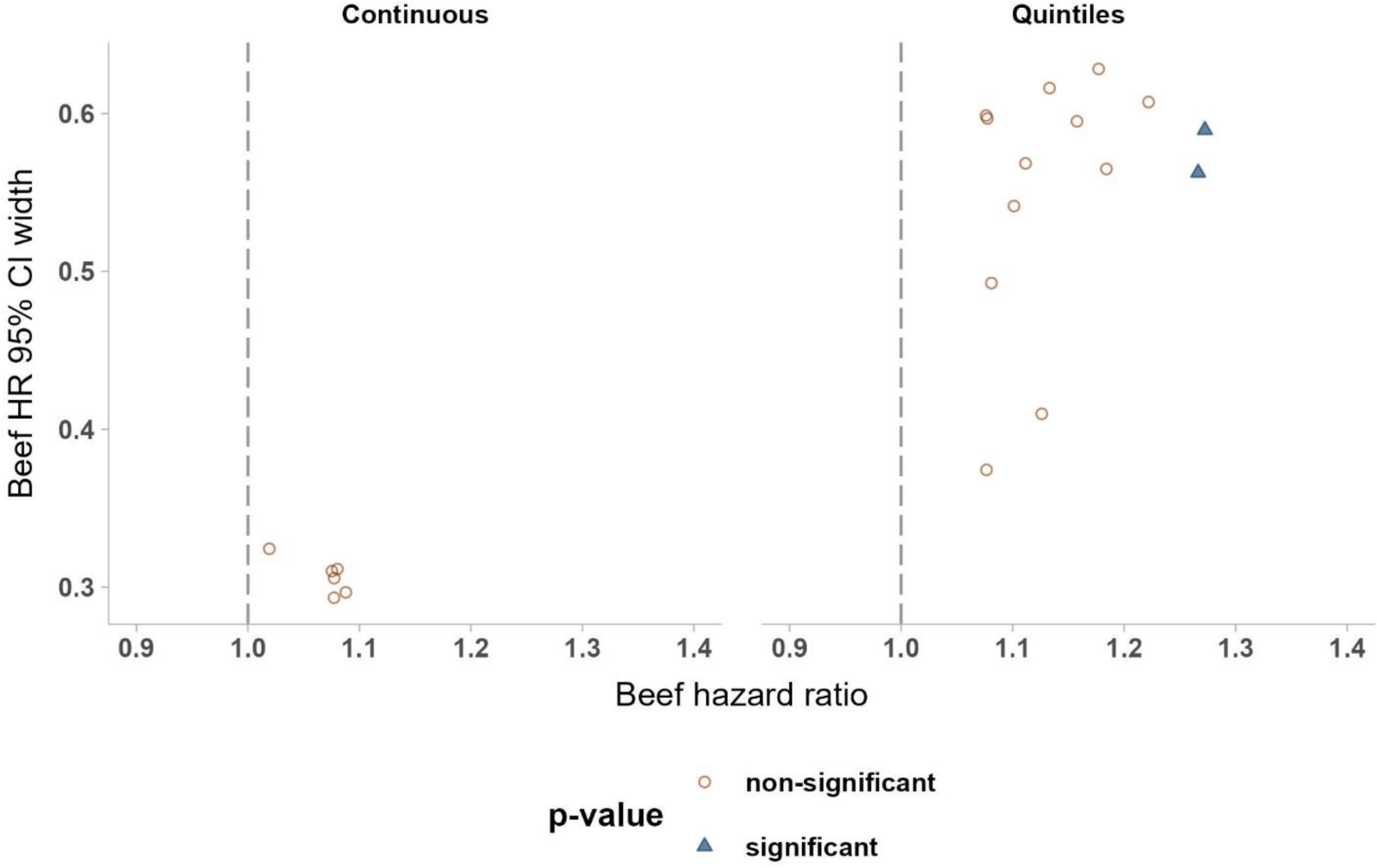
Scatterplot of HRs vs. 95% CI width for when self-reported beef consumption is expressed as continuous (left), and as quintiles of intake (right) for models emulating existing literature. Different symbols represent statistical significance at p<0.05 (filled triangle) versus non-significance (p≥0.05, circles)

**Figure 8.**
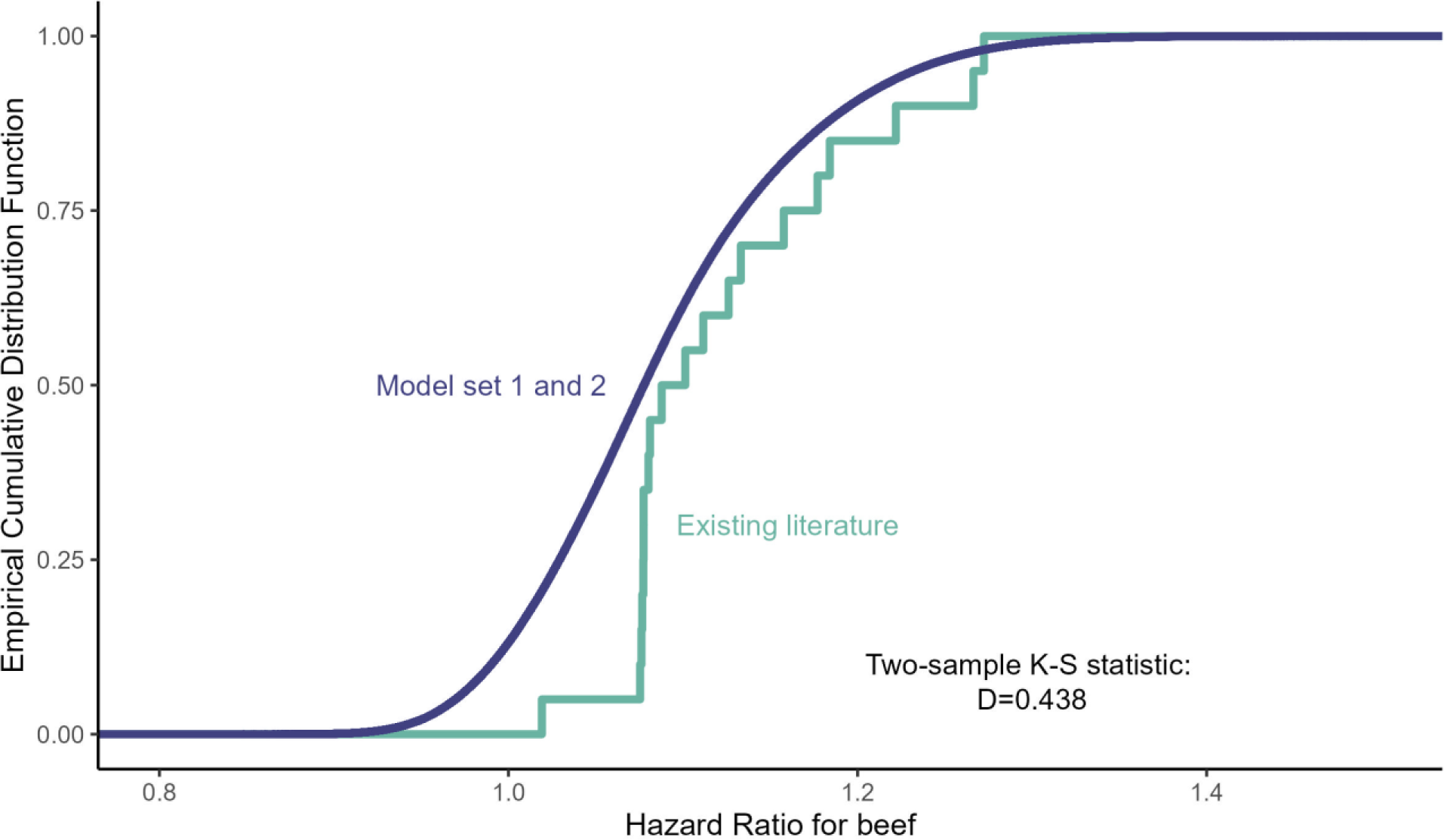
Comparison of the cumulative distribution between HRs from model sets 1 and 2 (combined) with HRs from models emulating existing literature. K-S statistic: Kolmogorov-Smirnov statistic.

**Table 3:** Descriptive statistics for models emulating existing literature.

|  | **Overall**<br>**(n=20)** | **Beef as Continuous<sup>1</sup>**<br>**(n=6)** | **Beef as Quintiles<sup>2</sup>**<br>**(n=14)** |
| --- | --- | --- | --- |
| **Hazard ratios (HR) for beef** |  |  |  |
| • Mean HR | 1.12 | 1.07 | 1.15 |
| • Median HR | 1.09 | 1.08 | 1.13 |
| • 5<sup>th</sup>, 95<sup>th</sup> percentile HR<sup>3</sup> | 1.05, 1.27 | 1.02, 1.09 | 1.08, 1.27 |
| • Min, max HR | 1.02, 1.27 | 1.02, 1.09 | 1.08, 1.27 |
| • N (%) of HR > 1.00<sup>4</sup> | 20 (100%) | 6 (100%) | 14 (100%) |
| • N (%) of significant HR (p<0.05) <sup>5</sup> | 2 (10%) | 0 (0%) | 2 (14%) |
| • N (%) of significant HR > 1.00 | 2 (10%) | 0 (0%) | 2 (14%) |
| **95% Confidence Interval width for beef HR** |  |  |  |
| • Mean width | 0.48 | 0.31 | 0.55 |
| • Median width | 0.55 | 0.30 | 0.58 |
| • 5<sup>th</sup>, 95<sup>th</sup> percentile of width | 0.29, 0.62 | 0.29, 0.32 | 0.37, 0.63 |
| • Min, max width | 0.29, 0.63 | 0.29, 0.32 | 0.37, 0.63 |
| **Significance** |  |  |  |
| • Mean p-value | 0.33 | 0.37 | 0.31 |
| • Median p-value | 0.30 | 0.30 | 0.31 |
| • 5<sup>th</sup>, 95<sup>th</sup> percentile p-value | 0.04, 0.71 | 0.23, 0.81 | 0.04, 0.60 |
| • Min, max p-value | 0.04, 0.81 | 0.23, 0.81 | 0.04, 0.60 |
| **Z-score** |  |  |  |
| • Mean z-score | 1.07 | 0.93 | 1.13 |
| • Median z-score | 1.04 | 1.04 | 1.02 |
| • 5<sup>th</sup>, 95<sup>th</sup> percentile z-score | 0.38, 2.08 | 0.24, 1.21 | 0.52, 2.10 |
| • Min, max z-score | 0.24, 2.10 | 0.24, 1.21 | 0.52, 2.10 |

<sup>1</sup> Per 50g self-reported, estimated intake. <sup>2</sup> Hazard ratio of highest versus lowest. <sup>3</sup> 5% and 95% represent the actual 5<sup>th</sup> percent and 95<sup>th</sup> percent of the ranked distribution of HRs from the models fit, not a confidence interval around the HR. <sup>4</sup> comparing the number of positive associations by beef configuration: Chi-Square test: $\chi^2(1)=60707$ , $p<0.001$ . <sup>5</sup> significant at $p<0.05$ , comparing number of significant results by beef configuration: Chi-Square test: $\chi^2(1)=492$ , $p<0.001$ .

**Table 4:**
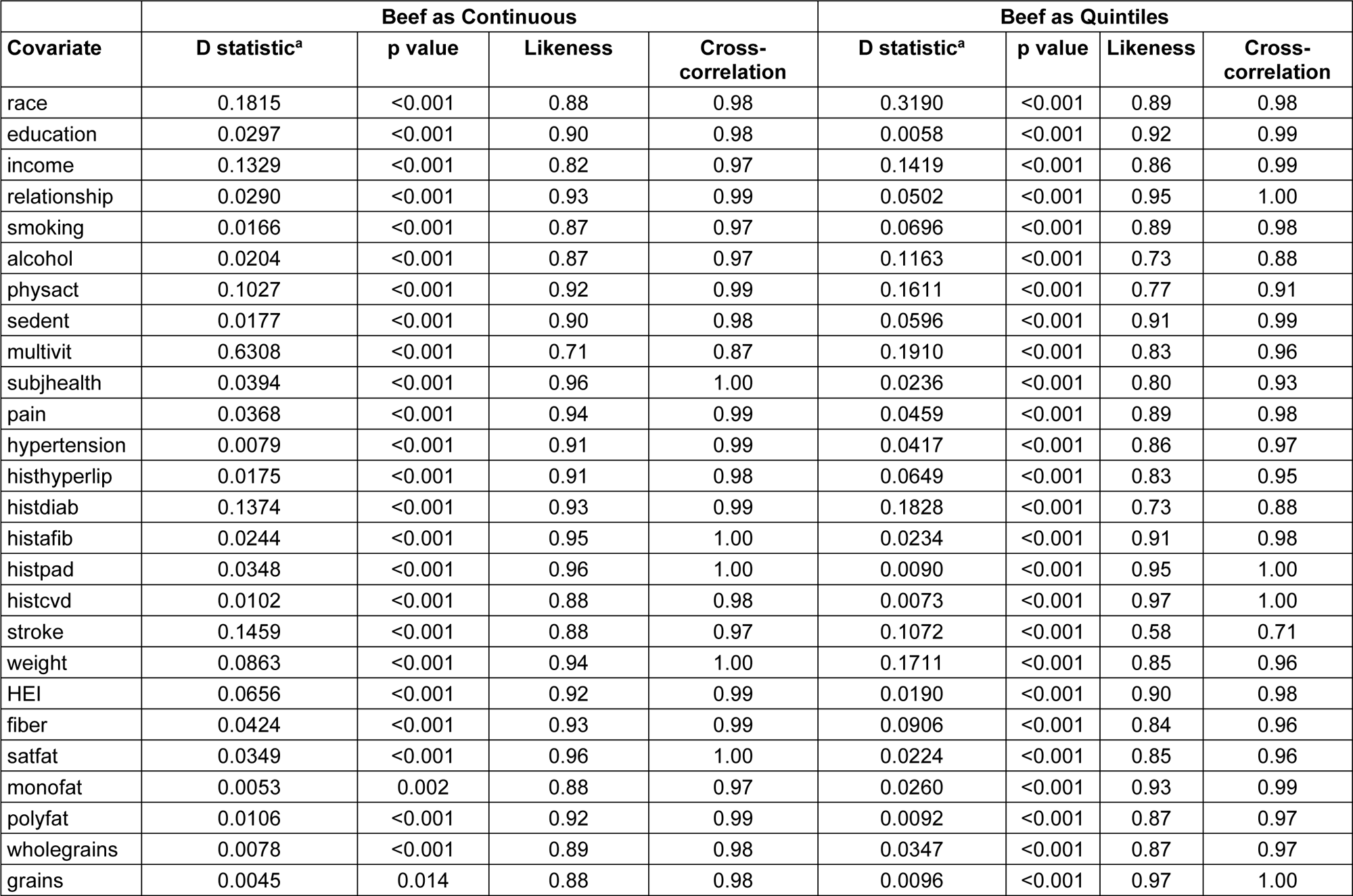
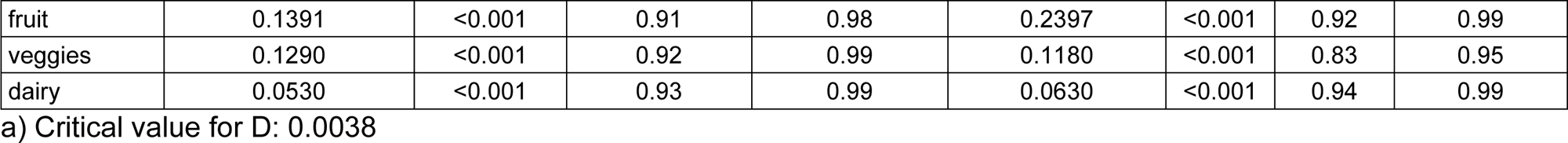
Results from Kolmogorov-Smirnov test comparing the distributions of HR for beef when including and excluding specific covariate.

## Discussion

Many analytic choices are needed when analyzing data from observational cohort studies. Historically, it was only feasible to analyze and report a handful of models, which represent only a small fraction of possible combinations. Indeed, by identifying covariates that have been used in the literature, we calculated over four quadrillion models that could be run to test the association between self-reported beef intake and incident CHD using the REGARDS dataset. Random sampling from these showed that HRs varied around the null of 1, and few models were statistically significant.

The results from our approach pose challenges to interpretation. Overall, the point estimates of the HRs were disproportionately above the null (87.9% overall; Table 2); however, a sizeable proportion of HRs remained less than the null (12.1%). Furthermore, less than 1% of individual models reached classical statistical significance thresholds of p<0.05, which is less than expected if results were derived by random chance. Yet, those that did were all in the deleterious to health direction. Also, all of the models are dependent (that is, they are based on the same underlying data), and thus benchmarking the number of statistically significant findings against traditional type I error metrics may be inappropriate. Therefore, the approach overall leaves some ambiguity regarding the association (let alone the causal relation) between beef and CHD from these data.

A qualitative inspection of our figures suggested that two variables had the greatest influence on results: years of multivitamin use and race. When each was excluded from models, HRs tended to be higher and model standard errors smaller. Multivitamin use is considered among health-related behaviors (34), and race is often considered with socioeconomic status (SES) (35). Not adjusting for these particular covariates, which indirectly capture concepts related to health consciousness and socioeconomic status, may produce more extreme results because of confounding. This raises the possibility that, even if one has an appropriate data generating process for selecting covariates to include based on a causal structure, covariate concepts may or may not be measured among different cohorts, or may be operationalized differently. For instance, SES is difficult to measure, so correlated indirect measures like race, income, and education may be used, but are still subject to unmeasured confounding. None of those measures alone fully capture SES, while adjusting for all of them results in multicollinearity; yet, choosing only one leads to measurement error and potentially high residual confounding. Thus, results may differ among models not because of any nefarious action by an epidemiologist, but because of which variables are available in a dataset and how they are operationalized or measured.

Given the inherent limitations of observational study designs (36, 37), the choice of covariates significantly influences the derived results. Notably, when we emulated models from published literature on meat-heart disease associations, all HRs were above 1, suggesting that those particular covariate choices tend to produce larger HRs than if one takes our agnostic approach. This discordance between our approach and the replications of other investigators’ models may or may not indicate the presence of publication or selection bias (38, 39) that drives the observation of published models exhibiting high effect sizes. Another possibility is that the effect size distribution from the replicated research better represents the underlying relation between beef and CHD. Without knowledge of the data-generating process, it is impossible to discern between these two scenarios or others.

Hypothesized data-generating processes (and thus any hypothesized causal structure) are rarely explicitly articulated in the choice of exposures, outcomes, and covariates in published literature. This leads to a crucial oversight in causal inference. The generation of a model should ideally be based on a robust mechanistic theory that justifies why a particular variable is a confounder, mediator, or collider (13). In this context, biases like collider bias, among others, are of significant concern, particularly when adjusting for measures such as energy intake (40). The practice of adhering to norms, such as including covariates for adjustment without a well-founded theoretical basis, might not be sufficient to account for these biases. We chose our approach because we generally do not observe that published articles on food- or nutrient-disease associations explicitly include a causal model with their analysis, and thus we wanted to evaluate the potential consequences of model selection in a way that emulates the current state of the literature. Indeed, a sample of 150 nutritional epidemiology studies found that 94% did not report *a priori* covariate selection, and only 20% reported the selection criteria for all covariates (17). Simulations have shown that flexibility in covariate selection can increase the chance of achieving statistical significance (10, 41–44). Together, the lack of a theoretical framework for any of our varied models raises the question of where on the HR distribution a true causal association may reside.

Including more covariates tended to decrease HRs in our models; this is consistent with accounting for more confounding. Yet, adding more covariates risks misspecification that could potentially bias results toward the null; however, such misspecification could also induce spuriously inflated associations, and we intuitively (though without empirical claim) find it unlikely that additional covariates would systematically bias toward versus away from the null in our permuted models. Thus, accounting for more covariates seems to weaken the argument for a causal association between beef and CHD in these models. Regardless, our approach does not necessarily resolve unmeasured confounders that systematically bias associations in either direction. For example, in cohorts from the U.S., higher self-reported consumers of red meat are more likely to self-report being less physically active, smoking, drinking alcohol, having higher body weight, and poorer diet quality compared to those who self-report lower red meat consumption (45–47).

Other studies have observed substantial variability in conclusions when different analysis strategies are used, such as asking different research teams to analyze the same data set (3, 4, 7, 8). Other methods have approached analysis strategy variability more systematically to evaluate the robustness of statistical findings to changes in model specification (specification curve analysis (14), multiverse analysis (12, 13), or vibration of effects (15)). The latter concept has been applied to nutritional questions to explore how including and excluding covariates influence the association between alpha-tocopherol and mortality, calcium and femur density, carrots and eyesight, and vitamin D level and COVID-19 (15, 16).

Our analysis is distinct from these approaches in that we varied both covariate inclusion and exclusion and covariate configuration, as well as exposure configuration. Importantly, our data generating process to select covariates was done agnostically, at random, which is not the intention of multiverse-style approaches that should carefully consider the causal structure of the research question to examine the robustness of the question to analytic decisions (13). We adapted some visualization methods developed for specification curve and vibration of effects analyses. Because we allowed our sample size to vary among models (consistent with a common complete-case approach in nutritional epidemiology), and our research question was not strictly held constant by nature of allowing model choice to vary, we chose not to compute an average p-value of all models (12), nor use a bootstrap technique (14). Future work is needed to improve quantitative interpretations when exploring many models and tease out analytic decisions that have a higher relative influence on associations.

There are limitations to our work that may be resolved in future research using these methods. For one, not all variables that we identified or their configurations in the literature could be exactly matched to REGARDS variables. Additional publications identified using different search strategies may identify additional variables or configurations to include in the analyses. Likewise, not all our modeling choices can be translated to different datasets to look at the same question. Many variables, including beef as our exposure of interest, were self-reported, and it is not clear how accurately intake is captured (36). In addition, we could not identify sufficient existing literature on beef *per se* and cardiovascular outcomes, so we used those on red meat more broadly, with the assumption that modeling choices would not differ. Further, two covariates used in previous literature, history of chronic obstructive pulmonary disease and sleep outcomes, did not closely match a variable in the REGARDS dataset, and therefore were missing in the distributions of results. Other modeling choices may be made by other analysts that may expand the decision tree even further and are not reflected in our analyses, such as excluding participants with a history of cancer at baseline (our rationale being that stronger associations may have been observed due to cardiotoxicity and cardiovascular deterioration in individuals with cancer); or using the ‘energy adjustment’ method (48) instead of including energy as a covariate. A particular challenge was to identify a dataset that permitted reasonable assessment of beef consumption specifically, rather than confounding the exposure of interest with other red or processed meats. Our estimation of self-reported beef from the FFQ used by REGARDS, using a proportion derived from 2017-2018 data from the Food and Nutrient Database for Dietary Studies was yet another point where various calculations may be considered reasonable and add additional model combinations, as well as the various ways to define beef such as total, unprocessed, processed, etc. (49). Although we used published literature to inform our covariable selection process, this does not necessarily mean that these covariates are those that all epidemiologists would deem as reasonable to include in models. In addition, some model combinations as randomly sampled may be less likely to be selected by epidemiologists than others, and thus our model distributions do not reflect models that would be weighted as more reasonable than others. Yet, because we included a subset of covariates in all of our models that are commonly included in observational studies, we believe that our models are all within the possibility of what qualified analysts might use. Our permutation approach currently has limitations in how many models can be evaluated because of computational limitations. Indeed, we discovered that only a fraction of the total possible models (quadrillions) can be feasibly run with current resources. We therefore leave open future investigations to run more or targeted sampling to refine the distributions or further investigate features of the HR distribution space. Even then, we could have varied more choices in our model and increased the model space exponentially, such as using additional covariates based on different sets of literature, how beef is defined, whether certain variables should be recoded or not, whether each covariate’s chance of being excluded is the same percentage as each included configuration, and so on. Finally, each way to express a model changes the research question in subtle ways, and thus we wish to emphasize that our approach does not necessarily assess the robustness of a particular question, but rather how it may vary when expressed in different ways (13, 40).

When there are not strong theory-based reasons to utilize specific statistical models for nutrition epidemiology questions, the approach we present herein may be useful to increase transparency and assess the distribution of results across many possible models. This approach may be facilitated by incorporating into standard workflows, and improving the availability of datasets used for nutrition epidemiology research questions (36).

## Abbreviations

CHD: Coronary Heart Disease
REGARDS: REasons for Geographic and Racial Differences in Stroke
HR: hazard ratio

## Glossary of terms

Covariate inclusion: Whether a covariate is included in a model.
Covariate configuration: How covariates are defined in a model; for example, as continuous, categorical, original, etc.
Model specification: All choices that go into a model, such as covariate inclusion, covariate configuration, and model type.
Quantile: Cut points that divide a distribution into intervals with equal likelihood.
Quintile: A quantile that divides a distribution into five intervals (i.e., there are four quintiles that make five intervals).
Effect size: “a quantitative reflection of the magnitude of some phenomenon that is used for the purpose of addressing a question of interest.” (1)

## Acknowledgements

The authors acknowledge the Indiana University Pervasive Technology Institute for providing supercomputing, storage, and consulting resources that have contributed to the research results reported within this paper. This research was supported in part by Lilly Endowment, Inc., through its support for the Indiana University Pervasive Technology Institute.

## Authors’ contributions

CJV, LEO, BH, AWB designed research; CJV, LEO, BH, CH, JS, CAS, RH, SLD, KE, AB, DBA, AWB conducted research; BH, CAS, SLD analyzed data or performed statistical analysis; CJV, LEO, BH, AWB wrote paper; AWB had primary responsibility for final content. All authors critically reviewed, edited, and approved the final manuscript.

## Data and Code Availability

Researchers who wish to reproduce our analyses can submit a project proposal to the REGARDS team (28). Code to reproduce our analyses is publicly available: https://osf.io/sy96k/

## Acknowledgements

The REGARDS study is supported by cooperative agreement U01 NS041588 co-funded by the National Institute of Neurological Disorders and Stroke (NINDS) and the National Institute on Aging (NIA), National Institutes of Health, Department of Health and Human Service. Additional funding for REGARDS CHD outcomes was provided by R01HL080477. The content is solely the responsibility of the authors and does not necessarily represent the official views of the NINDS, NHLBI, or the NIA. Representatives of the NINDS were involved in the review of the manuscript but were not directly involved in the collection, management, analysis or interpretation of the data. The authors thank the other investigators, the staff, and the participants of the REGARDS study for their valuable contributions. A full list of participating REGARDS investigators and institutions can be found at: https://www.uab.edu/soph/regardsstudy/

## Supplemental File Descriptions

**Supplemental Tables and Figures.** Supplemental tables and figures referenced within the text.

**Supplemental File 1**. Plots that show the distribution of HRs vs. SE for each covariate when inclusion/exclusion is varied, when self-reported beef consumption is expressed as continuous (left) and as quintiles of intake (right). Lines are percentile contours from a kernel density estimation using a normal distribution kernel.

**Supplemental File 2**. Plots that show the distribution of HRs vs. SE for each covariate when configuration is varied, when self-reported beef consumption is expressed as continuous (left) and as quintiles of intake (right). Lines are percentile contours from a kernel density estimation using a normal distribution kernel.

**Supplemental File 3**. Specification curves that show the distribution of HRs for the association between self-reported beef consumption and CHD, when beef is expressed as a continuous variable. Each combination of covariates is represented if a vertical line were traced from any point on the curve on top down through each variable underneath.

**Supplemental File 4**. Specification curves that show the distribution of HRs for the association between self-reported beef consumption and CHD, when beef is expressed as quintiles of intake. Each combination of covariates is represented if a vertical line were traced from any point on the curve on top down through each variable underneath.

