## Supplemental Tables and Figures for "‘Shaking the Ladder’ reveals how analytic choices can influence associations in nutrition epidemiology: beef intake and coronary heart disease as a case study"

**Supplemental Table 1.** Changes to our actual analysis plan compared to our preregistered analysis plan.

| **Variable concept** | **Reason for change** |
| --- | --- |
| Survey weights | Did not use because none of REGARDS analyses used them. |
| State | Did not use; models wouldn’t converge because too few participants in some of the states. |
| Energy intake | We originally planned to use the nutrient density method, but instead included energy as a covariate in all models. We did so primarily to directly compare beef as continuous intake per 50g unit increase vs. quintiles of intake, which happened to also be 50g. |
| Cancer | It was originally listed as a covariate as well as an exclusion criteria; we dropped it as a covariate. |
| Multivitamin use | ‘YrsMult’ did not list “no multivitamin use” as an option, so we added it. |
| Fruit | Was missing a continuous form, so it was added. |
| Strata for weighted sampling | Was not included. |
| Alcohol: ‘alc_6cat’ | We combined two categories (>2-4 and >4 drinks per day). |
| Physical activity: ‘exercise_cat’ | Originally we only listed “1 to 3 time per week” and “4 or more per week” as options; we added “none/0 times” as an option like other variables. |
| Model set 3 combinations | We included these emulations of existing literature because these analyses represent expert, peer-reviewed model specifications to benchmark against our ‘naïve’ approach of random covariate inclusion and configuration selection. |

**Supplemental Table 2.** Exclusions based on missing data for self-reported energy intake.

| # | Description | Excluded | Analytic sample |
| --- | --- | --- | --- |
| 1 | <500 kcal/day or >4500 kcal/day | n= 41 | 18748 |
| 2 | <600 kcal/day (2.5 MJ/day) or >5,000 kcal/day (20.9 MJ/day) | n=181 | 18608 |
| 3 | < 500 or > 3500kcal/d | n=405 | 18384 |
| 4 | upper and lower 2.5% (688, 2495) | n=814 | 17975 |
| 5 | more than 2 interquartile ranges above the 75th percentile or below the 25th percentile (0, 3858) | n=211 | 18578 |
| 6 | < 500 to > 4000 kcal/d | n=152 | 18637 |
| 7 | Sex-specific cutoff (<800 or >5000 kcal/d in men, <500 or >4500 kcal/d in women) (same as Regards exclusion) | n= 0 | 18789 |

Number excluded for each energy intake definition, which varied as part of model specifications. Exclusion numbers are prior to other exclusions from missing data from other variables.

**Supplemental Table 3.** Covariates and their configurations as defined using REGARDS data, and the number of missing values for each variable.

| **Variable** | **Omission as part of multi-verse** | **Configuration** | **Brief description of configuration** | **Missing values in conf.** | **Number of models with conf.** |
| --- | --- | --- | --- | --- | --- |
| Beef | No | beef2 | continuous (unit=50g) | 0 | 499564 |
|  |  | beef2_Q5 | quintiles | 0 | 500436 |
| Age | No | age_3cat | 45-54, 55-64, >=65 years | 0 | 333204 |
|  |  | age_5cat | 45-49, 50-59, 60-69, 70-79, >=80 y | 0 | 332860 |
|  |  | age | Continuous | 0 | 333936 |
| Gender | No | Gender | Female vs male | 0 | 1000000 |
| Calories | No | calories_MS | Sex-specific median split | 2482 | 333603 |
|  |  | calories_Q5 | Quintiles | 2482 | 333050 |
|  |  | calories | Continuous | 2482 | 333347 |
| Size of census tract | No | Urbangrp | Rural vs Mixed vs Urban | 1817 | 1000000 |
| Regards region | No | REGION | Belt vs Buckle vs Nonbelt | 0 | 1000000 |
| Race | Yes | Race | White vs Black | 0 | 500191 |
|  |  | excluded |  | - | 499809 |
| Education | Yes | ED_Cat | <HS, HS, Some College, College+ | 11 | 499542 |
|  |  | excluded |  | - | 500458 |
| Income | Yes | Income | 1 (<5K), 2 (5-10K), 3 (10-15K), 4 (15-20K), 5 (20-25K), 6 (25-35K), 7 (35-50K), 8 (50-75K), 9 (75-150K), 10 (>150K) | 2298 | 249899 |
|  |  | Income_4cat | <$20K, $20K-$35K, $35K-75K, $75K+, Refused | 0 | 250500 |
|  |  | excluded |  | - | 499601 |
| Marital status | Yes | Relationshipstatus | married, single, divorced, widowed, other | 0 | 499649 |
|  |  | excluded |  | - | 500351 |
| Smoking | Yes | Packyears | Number of packs smoked per year | 539 | 125280 |
|  |  | Smoke | Current, past, never | 67 | 125150 |
|  |  | Smoke_100cigs | Yes, no | 66 | 124619 |
|  |  | Smoke_current | Yes, no | 67 | 125173 |
|  |  | excluded |  | - | 499778 |
| Alcohol | Yes | Alc_Drinks_Wk | Alcoholic drinks per week | 333 | 99790 |
|  |  | Alc_NIAAA | NIAAA: 0=none, 1=moderate (0-7/wk women, 0-14/wk men) 2=heavy (7+/wk women 14+/wk men) | 0 | 100031 |
|  |  | alc_6cat | none, >0-0.5, >0.5-1, >1-2, >2-4, >4 drinks per day (combined last 2) | 333 | 100308 |
|  |  | alc_4cat | none, 1–4, 5–29, 30 + times/month | 333 | 100153 |
|  |  | Alc_Use | Never, Current, Past | 0 | 100059 |
|  |  | excluded |  | - | 499659 |
| Physical Activity | Yes | Exercise_3cat | 1=None, 2=1 to 3 times, 3=4 or more | 268 | 166374 |
|  |  | Exercise_5cat | none, 0 to <2, 2 to <4, 4 to <6, >6 times/week | 268 | 166970 |
|  |  | num_q8_1 | Times per week engage in intense physical activity, enough to work up a sweat | 268 | 166051 |
|  |  | excluded |  | - | 500605 |
| Sedentary Time | Yes | TV_video_3cat | none, <4 hours/d, >=4 hours/day | 1054 | 249302 |
|  |  | TV_video_6cat | none, 1-6 hrs/wk, 1 hr/day, 2 hrs/day, 3 hrs/day, 4+ hrs/day | 1054 | 250063 |
|  |  | excluded |  | - | 500635 |
| Weight | Yes | BMI | Continuous | 122 | 100081 |
|  |  | BMI_4cat | Underweight, normal, overweight, obese | 122 | 100176 |
|  |  | BMI_5cat | <18.5, 18.5 to <25, 25 to <30, 30 to <35, =35 | 122 | 100536 |
|  |  | BMI_Q5 | Quintiles | 122 | 99814 |
|  |  | Waist_cm | Waist circumference (continuous) | 128 | 100374 |
|  |  | excluded |  | - | 499019 |
| Multi­vitamin use | Yes | YrsMult | Years took multivitamins (0=No vitamins taken in past year, 1=Less than 1 year, 2=1 Year, 3=2 Years, 4=3-4 Years, 5=5-9 Years, 6=10+ Years) | 3573 | 500616 |
|  |  | excluded |  | - | 499384 |
| Subjective Health | Yes | Gen_SR_Health | Excellent, very good, good, fair, poor | 30 | 499530 |
|  |  | excluded |  | - | 500470 |
| Aspirin use | Yes | Reg_ASA | Yes, no | 9 | 249406 |
|  |  | Reg_NSAID | Yes, no | 54 | 250424 |
|  |  | excluded |  | - | 500170 |
| Stroke | Yes | Fram_stroke | Framingham Stroke Risk Score: 10 Year Probability of Stroke (%) (among those who self-reported never having a stroke at baseline) | 2203 | 99994 |
|  |  | ARICStroke | ARIC Stroke Risk Score: 10 Year Probability of Ischemic Stroke (%) | 2064 | 100383 |
|  |  | Stroke_Sym_Number | Number of stroke symptoms | 876 | 100247 |
|  |  | Stroke_SR | Participant reported stroke at baseline | 51 | 100078 |
|  |  | Stroke_Sympt | Presence of stroke symptoms | 876 | 100041 |
|  |  | excluded |  | - | 499257 |
| Hyper­tension | Yes | DBP^1^ | Diastolic BP (continuous) | 50 | 99931 |
|  |  | SBP^1^ | Systolic BP (continuous) | 49 | 99931 |
|  |  | Hyper_Meds_SR_now | Yes, no (SR current use of anti-hypertensive medication) | 740 | 99392 |
|  |  | Hyper_SRmeds_BP | Yes, no (Hypertensive if SBP>=140 or DBP>=90 or SR current medication use to control BP) | 42 | 100392 |
|  |  | Hyper_Meds_SR_ever | Yes, no (Self-reported ever having used anti-hypertensive medication) | 196 | 100016 |
|  |  | Hyper_SR | Yes, no (SR hypertension) | 90 | 99770 |
|  |  | excluded |  | - | 500499 |
| Diabetes | Yes | Diab_SRMed_glu | Yes, no (Diabetic if fasting glucose >= 126 / non-fasting glucose >= 200 or pills or insulin) | 640 | 499882 |
|  |  | excluded |  | - | 500118 |
| History of Hyper­lipidemia | Yes | Lipidemia_Meds_Labs | Yes, no (Dyslipidemia: if TC >= 240 or LDL >= 160 or HDL <= 40 or on medication | 664 | 249532 |
|  |  | Lipidemia_SR | Yes, no (SR elevated lipids) | 182 | 249969 |
|  |  | excluded |  |  | 500499 |
| History AFIB | Yes | AFib_ECG | Yes, no (Participant had ECG evidence of atrial fibrillation) | 223 | 166741 |
|  |  | Afib_SR | Yes, no (Q2_2D) | 182 | 166701 |
|  |  | AFib_SR_ECG | Yes, no (Atrial Fibrillation) | 377 | 167097 |
|  |  | excluded |  | - | 499461 |
| History PAD | Yes | PAD_surgery | Yes, no (SR procedure to fix the arteries in legs) | 18 | 499910 |
|  |  | excluded |  | - | 500090 |
| History CVD | Yes | CAD_aneurysm | Yes, no (SR repair of aortic aneurysm) | 28 | 125062 |
|  |  | Q3_2 | Yes, no (Ever had a surgery or procedure on the arteries in neck) | 0 | 125114 |
|  |  | Q3_4 | Yes, no (Ever had a pacemaker implanted) | 0 | 125352 |
|  |  | Q3_7 | Yes, no (Ever had any other heart or blood vessel surgery) | 0 | 125044 |
|  |  | excluded |  | - | 499428 |
| HEI | Yes | HEI_cat | good diet (HEI>=81), needs improvement (51<=HEI<81), poor diet (HEI<51) | 2482 | 125037 |
|  |  | H_Eat | continuous | 2482 | 124962 |
|  |  | H_Eat_MS | Sex-specific median split | 2482 | 125456 |
|  |  | H_Eat_Q5 | Quintiles | 2482 | 124467 |
|  |  | excl |  | - | 500078 |
| Dairy | Yes | DAIRYSRV | continuous | 2482 | 166667 |
|  |  | DAIRYSRV_MS | Sex-specific median split | 2482 | 166671 |
|  |  | DAIRYSRV_Q5 | Quintiles | 2482 | 166666 |
|  |  | excl |  | - | 499996 |
| Vegetables | Yes | GLOBVEG_MS | continuous | 3671 | 166667 |
|  |  | GLOBVEG_Q5 | Sex-specific median split | 3671 | 166664 |
|  |  | GLOBVEG_cont | Quintiles | 3671 | 166662 |
|  |  | excl |  | - | 500007 |
| Fruit | Yes | GLOBFRT_MS | Sex-specific median split | 3361 | 166673 |
|  |  | GLOBFRT_Q5 | Quintiles | 3361 | 166664 |
|  |  | GLOBFRT_cont | continuous | 3361 | 166661 |
|  |  | excl |  | - | 500002 |
| Grains | Yes | GRAINSRV | continuous | 2482 | 166667 |
|  |  | GRAINSRV_MS | Sex-specific median split | 2482 | 166668 |
|  |  | GRAINSRV_Q5 | Quintiles | 2482 | 166666 |
|  |  | excl |  | - | 499999 |
| Whole grains | Yes | Wholegrain | continuous | 2482 | 166664 |
|  |  | Wholegrain_MS | Sex-specific median split | 2482 | 166672 |
|  |  | Wholegrain_Q5 | Quintiles | 2482 | 166665 |
|  |  | excl |  | - | 499999 |
| Poly­un­saturated Fatty Acids | Yes | POLY_FAT | continuous | 2482 | 166668 |
|  |  | POLY_FAT_MS | Sex-specific median split | 2482 | 166668 |
|  |  | POLY_FAT_Q5 | Quintiles | 2482 | 166665 |
|  |  | excl |  | - | 499999 |
| Mono­un­saturated Fatty Acids | Yes | MONO_FAT | continuous | 2482 | 166666 |
|  |  | MONO_FAT_MS | Sex-specific median split | 2482 | 166665 |
|  |  | MONO_FAT_Q5 | Quintiles | 2482 | 166667 |
|  |  | excl |  | - | 500002 |
| Saturated Fats | Yes | SAT_FAT | continuous | 2482 | 166668 |
|  |  | SAT_FAT_MS | Sex-specific median split | 2482 | 166669 |
|  |  | SAT_FAT_Q5 | Quintiles | 2482 | 166666 |
|  |  | excl |  | - | 499997 |
| Fiber | Yes | FIBER | continuous | 2482 | 166664 |
|  |  | FIBER_MS | Sex-specific median split | 2482 | 166671 |
|  |  | FIBER_Q5 | Quintiles | 2482 | 166661 |
|  |  | excl |  | - | 500004 |
| Exclusion criteria | No | cal_excl_1 | <500 kcal/day or >4500 kcal/day | 2482 | 142527 |
|  |  | cal_excl_2 | <600 kcal/day (2.5 MJ/day) or >5,000 kcal/day (20.9 MJ/day) | 2482 | 141981 |
|  |  | cal_excl_3 | < 500 or > 3500kcal/d | 2482 | 143273 |
|  |  | cal_excl_4 | upper and lower 2.5% (688, 2495) | 2482 | 142949 |
|  |  | cal_excl_5 | more than 2 interquartile ranges above the 75th percentile or below the 25th percentile (0, 3858) | 2482 | 143530 |
|  |  | cal_excl_6 | < 500 to > 4000 kcal/d | 2482 | 142631 |
|  |  | cal_excl_7 | Sex-specific cutoff (<800 or >5000 kcal/d in men, <500 or >4500 kcal/d in women) (same as Regards exclusion) | 2482 | 143109 |

Abbreviations: Conf.=configuration. 1 dual component model: if included: systolic and diastolic blood pressure were always both in the model, 2 for dietary variables: if included, all dietary variables had the same configuration.

**Supplemental Table 4**: Results from multivariable logistic regression on the association between the beef coefficient being significant and inclusion/configuration of covariates.

| **Concept** | **Configuration** | **Model set 1 (beef as continuous)** | | | | **Model set 2 (beef as highest quintile)** | | | |
| --- | --- | --- | --- | --- | --- | --- | --- | --- | --- |
|  |  | **% of significant beef HRs** | **OR (95% CI)^1^** | **P value** | **Type 3 p-value** | **% of significant beef HRs** | **OR (95% CI)^1^** | **p-value** | **Type 3 p-value** |
| age | Age | 1.09% | Ref. |  | <.001 | 1.93% | Ref. |  | <.001 |
|  | age_3cat | 0.42% | 0.13 (0.11, 0.14) | <.001 |  | 0.40% | 0.04 (0.04, 0.05) | <.001 |  |
|  | age_5cat | 0.71% | 0.37 (0.33, 0.42) | <.001 |  | 1.17% | 0.37 (0.34, 0.41) | <.001 |  |
| cal_base | CALORIES | 0.80% | Ref. |  | <.001 | 1.26% | Ref. |  | <.001 |
|  | calories_MS | 1.08% | 2.15 (1.93, 2.40) | <.001 |  | 1.40% | 1.37 (1.26, 1.50) | <.001 |  |
|  | calories_Q5 | 0.34% | 0.18 (0.16, 0.21) | <.001 |  | 0.86% | 0.46 (0.41, 0.50) | <.001 |  |
| subset2 | cal_excl_1 | 0.27% | 0.21 (0.16, 0.26) | <.001 |  | 0.76% | 0.44 (0.37, 0.51) | <.001 |  |
|  | cal_excl_2 | 0.62% | 1.03 (0.85, 1.25) | 0.765 |  | 0.75% | 0.43 (0.37, 0.50) | <.001 |  |
|  | cal_excl_3 | 1.52% | 7.65 (6.43, 9.10) | <.001 |  | 1.11% | 0.93 (0.81, 1.07) | 0.298 |  |
|  | cal_excl_4 | 1.74% | 11.61 (9.76, 13.8) | <.001 |  | 3.36% | 11.64 (10.28, 13.19) | <.001 |  |
|  | cal_excl_5 | 0.29% | 0.23 (0.19, 0.30) | <.001 |  | 0.54% | 0.25 (0.21, 0.29) | <.001 |  |
|  | cal_excl_6 | 0.18% | 0.13 (0.10, 0.17) | <.001 |  | 0.56% | 0.25 (0.21, 0.29) | <.001 |  |
|  | cal_excl_7 | 0.56% | Ref. |  | <.001 | 1.11% | Ref. |  | <.001 |
| alcohol | excl | 0.70% | Ref. |  | <.001 | 0.76% | Ref. |  | <.001 |
|  | alc_4cat | 0.71% | 1.23 (1.04, 1.46) | 0.015 |  | 1.93% | 7.10 (6.31, 8.00) | <.001 |  |
|  | alc_6cat | 0.74% | 1.16 (0.98, 1.36) | 0.086 |  | 2.03% | 7.72 (6.86, 8.69) | <.001 |  |
|  | Alc_Drinks_Wk | 0.54% | 0.61 (0.50, 0.73) | <.001 |  | 1.14% | 2.14 (1.87, 2.46) | <.001 |  |
|  | Alc_NIAAA | 1.01% | 2.33 (2.00, 2.72) | <.001 |  | 1.56% | 4.30 (3.79, 4.87) | <.001 |  |
|  | Alc_Use | 0.88% | 1.83 (1.57, 2.14) | <.001 |  | 1.27% | 2.60 (2.28, 2.97) | <.001 |  |
| dairy | excl | 0.83% | Ref. |  | <.001 | 1.33% | Ref. |  | <.001 |
|  | DAIRYSRV | 0.69% | 0.64 (0.54, 0.75) | <.001 |  | 0.77% | 0.75 (0.64, 0.87) | <.001 |  |
|  | DAIRYSRV_MS | 0.85% | 0.57 (0.50, 0.66) | <.001 |  | 1.06% | 0.68 (0.60, 0.77) | <.001 |  |
|  | DAIRYSRV_Q5 | 0.39% | 0.42 (0.34, 0.51) | <.001 |  | 1.20% | 0.36 (0.32, 0.40) | <.001 |  |
| education | excl | 0.67% | Ref. |  | <.001 | 1.18% | Ref. |  | 0.061 |
|  | ED_Cat | 0.81% | 1.40 (1.27, 1.54) | <.001 |  | 1.16% | 0.93 (0.86, 1.00) | 0.061 |  |
| fiber | excl | 0.93% | Ref. |  | <.001 | 1.62% | Ref. |  | <.001 |
|  | FIBER | 0.46% | 0.14 (0.12, 0.17) | <.001 |  | 0.22% | 0.03 (0.03, 0.04) | <.001 |  |
|  | FIBER_MS | 0.78% | 0.45 (0.39, 0.52) | <.001 |  | 0.89% | 0.39 (0.34, 0.44) | <.001 |  |
|  | FIBER_Q5 | 0.40% | 0.46 (0.38, 0.56) | <.001 |  | 1.05% | 0.26 (0.23, 0.30) | <.001 |  |
| fruit | excl | 1.16% | Ref. |  | <.001 | 2.05% | Ref. |  | <.001 |
|  | GLOBFRT_cont | 0.30% | 0.04 (0.03, 0.04) | <.001 |  | 0.15% | 0.01 (0.01, 0.02) | <.001 |  |
|  | GLOBFRT_MS | 0.52% | 0.11 (0.09, 0.13) | <.001 |  | 0.34% | 0.04 (0.03, 0.04) | <.001 |  |
|  | GLOBFRT_Q5 | 0.13% | 0.02 (0.02, 0.03) | <.001 |  | 0.41% | 0.02 (0.02, 0.03) | <.001 |  |
| grains | excl | 0.72% | Ref. |  | <.001 | 1.17% | Ref. |  | <.001 |
|  | GRAINSRV | 0.86% | 1.64 (1.39, 1.92) | <.001 |  | 0.77% | 0.72 (0.62, 0.84) | <.001 |  |
|  | GRAINSRV_MS | 0.96% | 0.98 (0.85, 1.13) | 0.758 |  | 1.15% | 1.08 (0.96, 1.22) | 0.212 |  |
|  | GRAINSRV_Q5 | 0.46% | 0.89 (0.73, 1.07) | 0.212 |  | 1.61% | 1.30 (1.17, 1.45) | <.001 |  |
| HEI | excl | 1.06% | Ref. |  | <.001 | 1.21% | Ref. |  | <.001 |
|  | H_Eat | 0.32% | 0.06 (0.05, 0.08) | <.001 |  | 1.24% | 1.11 (0.99, 1.25) | 0.068 |  |
|  | H_Eat_MS | 0.67% | 0.35 (0.30, 0.40) | <.001 |  | 1.14% | 0.87 (0.77, 0.98) | 0.023 |  |
|  | H_Eat_Q5 | 0.34% | 0.08 (0.06, 0.09) | <.001 |  | 1.17% | 0.88 (0.78, 1.00) | 0.041 |  |
|  | HEI_cat | 0.33% | 0.08 (0.06, 0.10) | <.001 |  | 1.00% | 0.67 (0.6, 0.76) | <.001 |  |
| histafib | excl | 0.65% | Ref. |  | <.001 | 1.27% | Ref. |  | <.001 |
|  | AFib_ECG | 1.17% | 4.42 (3.90, 5.01) | <.001 |  | 1.87% | 2.18 (1.98, 2.39) | <.001 |  |
|  | Afib_SR | 0.45% | 0.43 (0.36, 0.51) | <.001 |  | 0.52% | 0.16 (0.14, 0.18) | <.001 |  |
|  | Afib_SR_ECG | 0.89% | 1.77 (1.55, 2.02) | <.001 |  | 0.82% | 0.42 (0.38, 0.48) | <.001 |  |
| histcvd | excl | 0.78% | Ref. |  | <.001 | 1.21% | Ref. |  | <.001 |
|  | CAD_aneurysm | 1.12% | 2.26 (1.98, 2.58) | <.001 |  | 1.46% | 1.45 (1.30, 1.62) | <.001 |  |
|  | Q3_2 | 0.52% | 0.44 (0.37, 0.53) | <.001 |  | 0.93% | 0.59 (0.52, 0.67) | <.001 |  |
|  | Q3_4 | 0.74% | 0.92 (0.79, 1.07) | 0.272 |  | 1.29% | 1.12 (1.00, 1.25) | 0.058 |  |
|  | Q3_7 | 0.40% | 0.26 (0.21, 0.31) | <.001 |  | 0.85% | 0.44 (0.39, 0.50) | <.001 |  |
| histdiab | excl | 1.24% | Ref. |  | <.001 | 1.96% | Ref. |  | <.001 |
|  | Diab_SRMed_glu | 0.24% | 0.04 (0.03, 0.04) | <.001 |  | 0.38% | 0.04 (0.04, 0.04) | <.001 |  |
| histhyperlip | excl | 0.80% | Ref. |  | <.001 | 1.09% | Ref. |  | <.001 |
|  | Lipidemia_meds_labs | 0.50% | 0.34 (0.30, 0.39) | <.001 |  | 0.83% | 0.60 (0.54, 0.66) | <.001 |  |
|  | Lipidemia_SR | 0.87% | 1.17 (1.05, 1.31) | 0.005 |  | 1.67% | 2.45 (2.25, 2.67) | <.001 |  |
| histpad | excl | 0.63% | Ref. |  | <.001 | 1.15% | Ref. |  | 0.129 |
|  | PAD_surgery | 0.85% | 1.94 (1.76, 2.14) | <.001 |  | 1.19% | 1.06 (0.98, 1.14) | 0.129 |  |
| hypertension | excl | 0.65% | Ref. |  | <.001 | 1.25% | Ref. |  | <.001 |
|  | Hyper_Meds_SR_ever | 0.57% | 0.70 (0.58, 0.84) | <.001 |  | 1.08% | 0.75 (0.65, 0.85) | <.001 |  |
|  | Hyper_Meds_SR_now | 1.90% | 13.66 (11.90, 15.68) | <.001 |  | 1.13% | 0.83 (0.73, 0.95) | 0.007 |  |
|  | Hyper_SR | 0.64% | 0.99 (0.83, 1.18) | 0.871 |  | 1.65% | 1.83 (1.63, 2.06) | <.001 |  |
|  | Hyper_SRmeds_BP | 0.73% | 1.42 (1.20, 1.68) | <.001 |  | 0.89% | 0.53 (0.46, 0.61) | <.001 |  |
|  | SBP + DBP | 0.27% | 0.23 (0.18, 0.29) | <.001 |  | 0.73% | 0.38 (0.32, 0.44) | <.001 |  |
| income | excl | 0.24% | Ref. |  | <.001 | 0.58% | Ref. |  | <.001 |
|  | Income | 2.11% | 85.42 (73.05, 99.88) | <.001 |  | 2.84% | 28.91 (26.04, 32.09) | <.001 |  |
|  | Income_4cat | 0.37% | 1.96 (1.67, 2.31) | <.001 |  | 0.70% | 1.43 (1.27, 1.60) | <.001 |  |
| monofat | excl | 0.79% | Ref. |  | <.001 | 1.04% | Ref. |  | <.001 |
|  | MONO_FAT | 0.73% | 0.87 (0.74, 1.02) | 0.081 |  | 0.86% | 1.08 (0.93, 1.25) | 0.317 |  |
|  | MONO_FAT_MS | 0.94% | 0.98 (0.85, 1.13) | 0.801 |  | 1.23% | 1.17 (1.04, 1.33) | 0.010 |  |
|  | MONO_FAT_Q5 | 0.38% | 0.40 (0.32, 0.48) | <.001 |  | 1.82% | 2.17 (1.94, 2.43) | <.001 |  |
| multivit | excl | 1.48% | Ref. |  | <.001 | 2.05% | Ref. |  | <.001 |
|  | YrsMult | 0.00% | 0.00 (0.00, 0.00) | <.001 |  | 0.29% | 0.02 (0.02, 0.03) | <.001 |  |
| pain | excl | 0.85% | Ref. |  | <.001 | 1.40% | Ref. |  | <.001 |
|  | Reg_Asa | 0.65% | 0.56 (0.50, 0.63) | <.001 |  | 1.05% | 0.55 (0.5, 0.60) | <.001 |  |
|  | Reg_Nsaids | 0.61% | 0.42 (0.37, 0.48) | <.001 |  | 0.83% | 0.3 (0.27, 0.33) | <.001 |  |
| physact | excl | 1.12% | Ref. |  | <.001 | 1.88% | Ref. |  | <.001 |
|  | Exercise_3cat | 0.37% | 0.08 (0.07, 0.10) | <.001 |  | 0.46% | 0.05 (0.05, 0.06) | <.001 |  |
|  | Exercise_5cat | 0.33% | 0.08 (0.07, 0.10) | <.001 |  | 0.54% | 0.07 (0.06, 0.08) | <.001 |  |
|  | num_q8_1 | 0.38% | 0.11 (0.09, 0.13) | <.001 |  | 0.40% | 0.05 (0.04, 0.06) | <.001 |  |
| polyfat | excl | 0.70% | Ref. |  | <.001 | 1.21% | Ref. |  | 0.016 |
|  | POLY_FAT | 0.87% | 1.56 (1.33, 1.83) | <.001 |  | 0.80% | 1.02 (0.88, 1.18) | 0.789 |  |
|  | POLY_FAT_MS | 0.98% | 0.99 (0.85, 1.14) | 0.838 |  | 1.13% | 0.93 (0.82, 1.05) | 0.222 |  |
|  | POLY_FAT_Q5 | 0.47% | 0.99 (0.82, 1.20) | 0.915 |  | 1.45% | 0.84 (0.76, 0.94) | 0.003 |  |
| race | excl | 1.24% | Ref. |  | <.001 | 1.97% | Ref. |  | <.001 |
|  | Race | 0.24% | 0.03 (0.03, 0.04) | <.001 |  | 0.38% | 0.04 (0.03, 0.04) | <.001 |  |
| relationship | excl | 0.68% | Ref. |  | <.001 | 0.96% | Ref. |  | <.001 |
|  | Relationshipstatus | 0.80% | 1.51 (1.37, 1.66) | <.001 |  | 1.38% | 2.29 (2.12, 2.47) | <.001 |  |
| satfat | excl | 0.99% | Ref. |  | <.001 | 1.41% | Ref. |  | <.001 |
|  | SAT_FAT | 0.43% | 0.11 (0.09, 0.13) | <.001 |  | 0.63% | 0.39 (0.33, 0.45) | <.001 |  |
|  | SAT_FAT_MS | 0.78% | 0.46 (0.40, 0.53) | <.001 |  | 0.92% | 0.43 (0.38, 0.49) | <.001 |  |
|  | SAT_FAT_Q5 | 0.26% | 0.10 (0.08, 0.13) | <.001 |  | 1.23% | 0.42 (0.38, 0.47) | <.001 |  |
| sedent | excl | 0.69% | Ref. |  | <.001 | 1.46% | Ref. |  | <.001 |
|  | TV_video_3cat | 0.69% | 1.09 (0.96, 1.23) | 0.189 |  | 0.76% | 0.27 (0.24, 0.30) | <.001 |  |
|  | TV_video_6cat | 0.88% | 1.78 (1.58, 1.99) | <.001 |  | 1.01% | 0.47 (0.43, 0.52) | <.001 |  |
| smoking | excl | 0.75% | Ref. |  | <.001 | 1.33% | Ref. |  | <.001 |
|  | Packyears | 1.30% | 3.68 (3.23, 4.19) | <.001 |  | 2.66% | 4.71 (4.29, 5.17) | <.001 |  |
|  | Smoke | 0.52% | 0.44 (0.37, 0.52) | <.001 |  | 0.34% | 0.09 (0.07, 0.11) | <.001 |  |
|  | Smoke_100cigs | 0.60% | 0.54 (0.46, 0.63) | <.001 |  | 0.59% | 0.21 (0.18, 0.24) | <.001 |  |
|  | Smoke_current | 0.51% | 0.44 (0.37, 0.53) | <.001 |  | 0.46% | 0.13 (0.11, 0.15) | <.001 |  |
| stroke | excl | 1.17% | Ref. |  | <.001 | 1.79% | Ref. |  | <.001 |
|  | ARICStroke | 0.12% | 0.01 (0.01, 0.02) | <.001 |  | 0.13% | 0.01 (0.01, 0.02) | <.001 |  |
|  | Fram_stroke | 0.01% | 0.00 (0.00, 0.00) | <.001 |  | 0.01% | 0.00 (0.00, 0.00) | <.001 |  |
|  | Stroke_SR | 1.00% | 0.63 (0.54, 0.72) | <.001 |  | 1.09% | 0.38 (0.33, 0.43) | <.001 |  |
|  | Stroke_Sym_Number | 0.23% | 0.04 (0.03, 0.05) | <.001 |  | 0.78% | 0.19 (0.17, 0.23) | <.001 |  |
|  | Stroke_Sympt | 0.18% | 0.02 (0.02, 0.03) | <.001 |  | 0.77% | 0.18 (0.16, 0.21) | <.001 |  |
| subjhealth | excl | 0.64% | Ref. |  | <.001 | 1.21% | Ref. |  | <.001 |
|  | Gen_SR_Health | 0.84% | 1.84 (1.67, 2.03) | <.001 |  | 1.13% | 0.79 (0.73, 0.85) | <.001 |  |
| veggies | excl | 0.33% | Ref. |  | <.001 | 1.28% | Ref. |  | <.001 |
|  | GLOBVEG_cont | 1.23% | 15.60 (12.99, 18.73) | <.001 |  | 0.72% | 0.63 (0.55, 0.73) | <.001 |  |
|  | GLOBVEG_MS | 1.44% | 12.00 (10.2, 14.11) | <.001 |  | 0.90% | 0.41 (0.36, 0.46) | <.001 |  |
|  | GLOBVEG_Q5 | 0.77% | 14.24 (11.51, 17.62) | <.001 |  | 1.57% | 1.20 (1.07, 1.34) | 0.002 |  |
| weight | excl | 0.99% | Ref. |  | <.001 | 1.77% | Ref. |  | <.001 |
|  | BMI | 0.49% | 0.21 (0.17, 0.25) | <.001 |  | 0.35% | 0.04 (0.04, 0.05) | <.001 |  |
|  | BMI_4cat | 0.50% | 0.23 (0.19, 0.28) | <.001 |  | 1.45% | 0.60 (0.54, 0.68) | <.001 |  |
|  | BMI_5cat | 0.48% | 0.25 (0.20, 0.30) | <.001 |  | 0.43% | 0.06 (0.05, 0.07) | <.001 |  |
|  | BMI_Q5 | 0.60% | 0.31 (0.26, 0.37) | <.001 |  | 0.54% | 0.09 (0.08, 0.11) | <.001 |  |
|  | Waist_cm | 0.35% | 0.11 (0.09, 0.14) | <.001 |  | 0.13% | 0.01 (0.01, 0.01) | <.001 |  |
| wholegrains | excl | 0.73% | Ref. |  | 0.153 | 1.21% | Ref. |  | <.001 |
|  | Wholegrain | 0.79% | 1.10 (0.93, 1.29) | 0.258 |  | 0.77% | 0.78 (0.68, 0.91) | 0.001 |  |
|  | Wholegrain_MS | 0.97% | 0.90 (0.78, 1.04) | 0.146 |  | 1.10% | 0.73 (0.65, 0.83) | <.001 |  |
|  | Wholegrain_Q5 | 0.48% | 0.88 (0.73, 1.06) | 0.183 |  | 1.53% | 1.09 (0.97, 1.22) | 0.131 |  |

^1^ Results from a multivariable logistic regression model on the association of the beef coefficient being significant (alpha<0.05) using the configurations of all covariates (including exclusion) as predictors, separately for continuous beef intake and beef intake expressed in quintile defined categories.

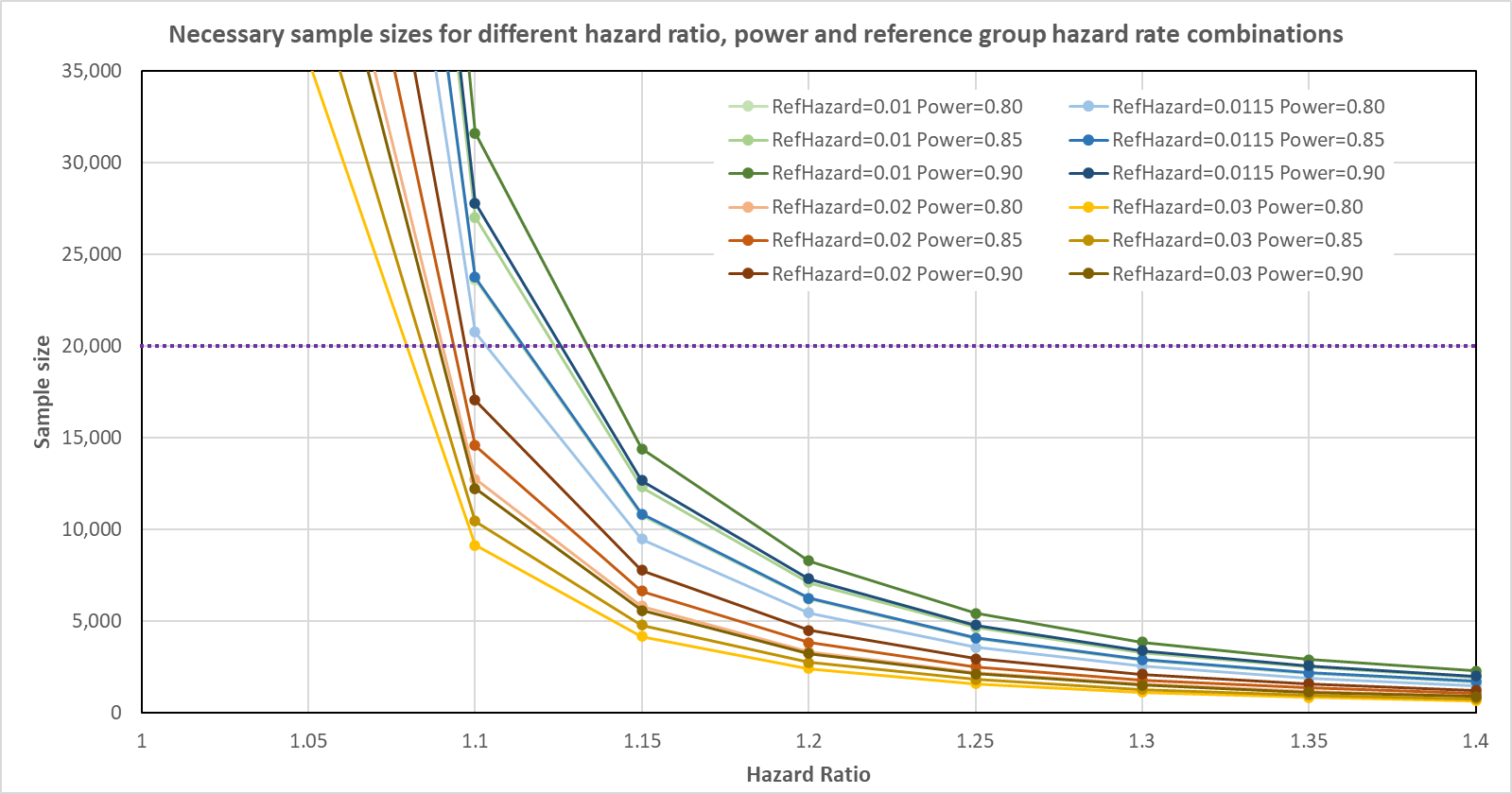

**Supplemental Figure 1**. Power analysis varying hazard ratio and power, using incident cardiovascular disease from the lowest quartile of estimated red meat consumption from Zhong et al. (1) as the reference hazard.
