## Supplemental File 1 for "‘Shaking the Ladder’ reveals how analytic choices can influence associations in nutrition epidemiology: beef intake and coronary heart disease as a case study"

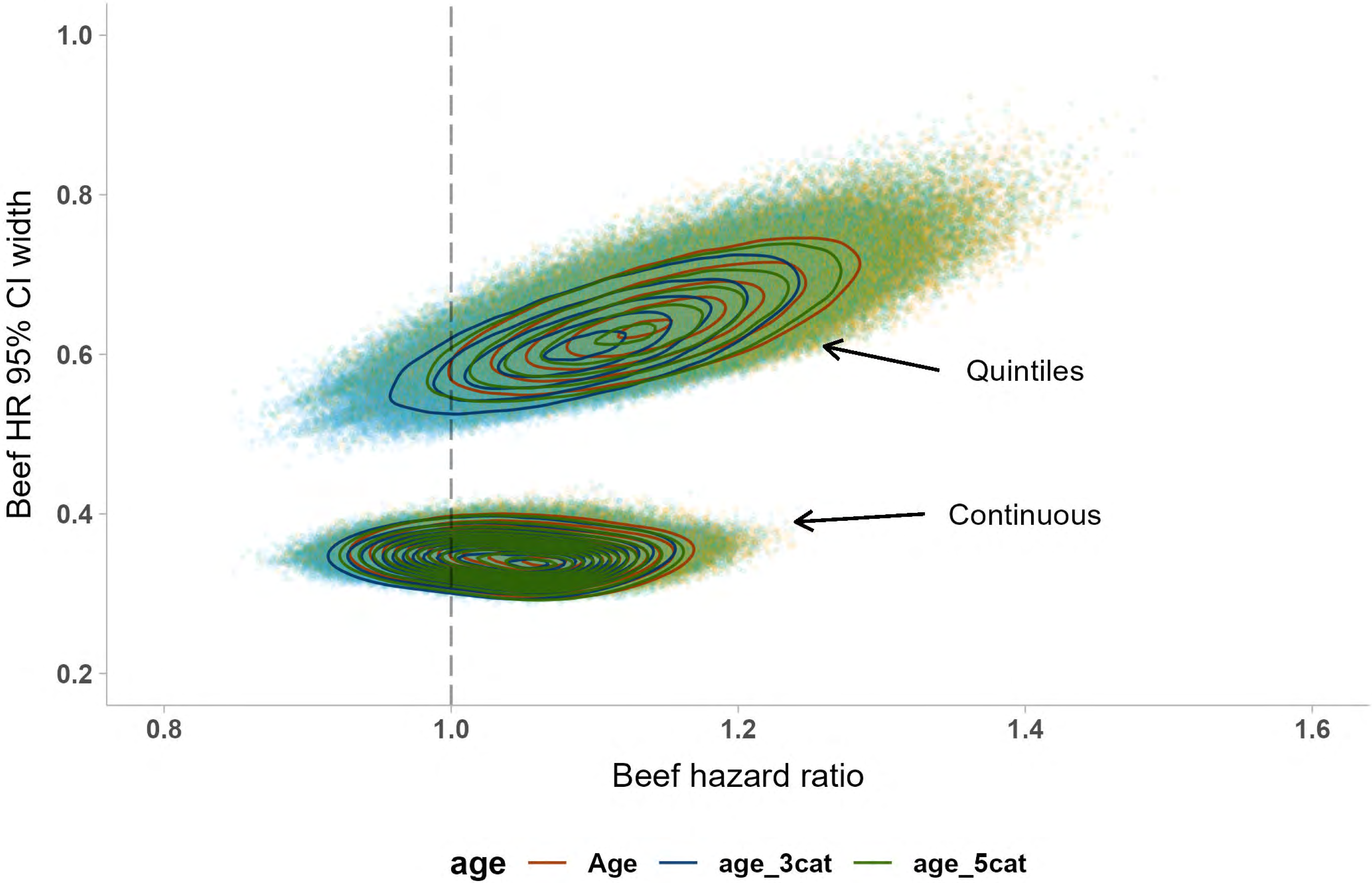

1.0
0.8
0.6
0.4
0.2
Beef HR 95% CI width
Quintiles
Continuous
0.8
1.0
1.2
1.4
1.6
Beef hazard ratio
age
Age
age_3cat
age_5cat

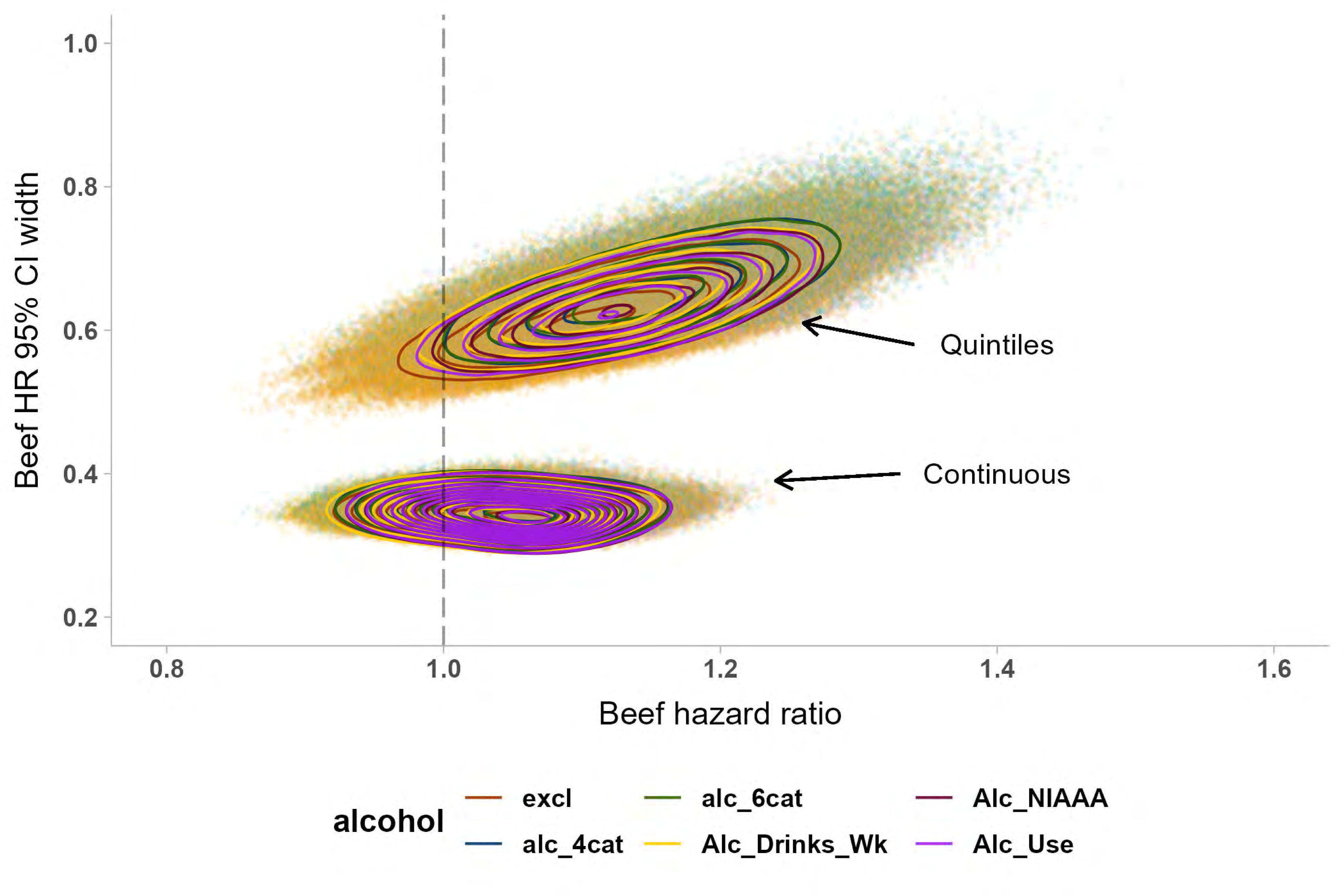

Beef HR 95% CI width
1.0
0.8
0.6
0.4
0.2
Quintiles
Continuous
0.8
1.0
1.2
1.4
1.6
Beef hazard ratio
alcohol
excl
alc_4cat
alc_6cat
Alc_Drinks_Wk
Alc_NIAAA
Alc_Use

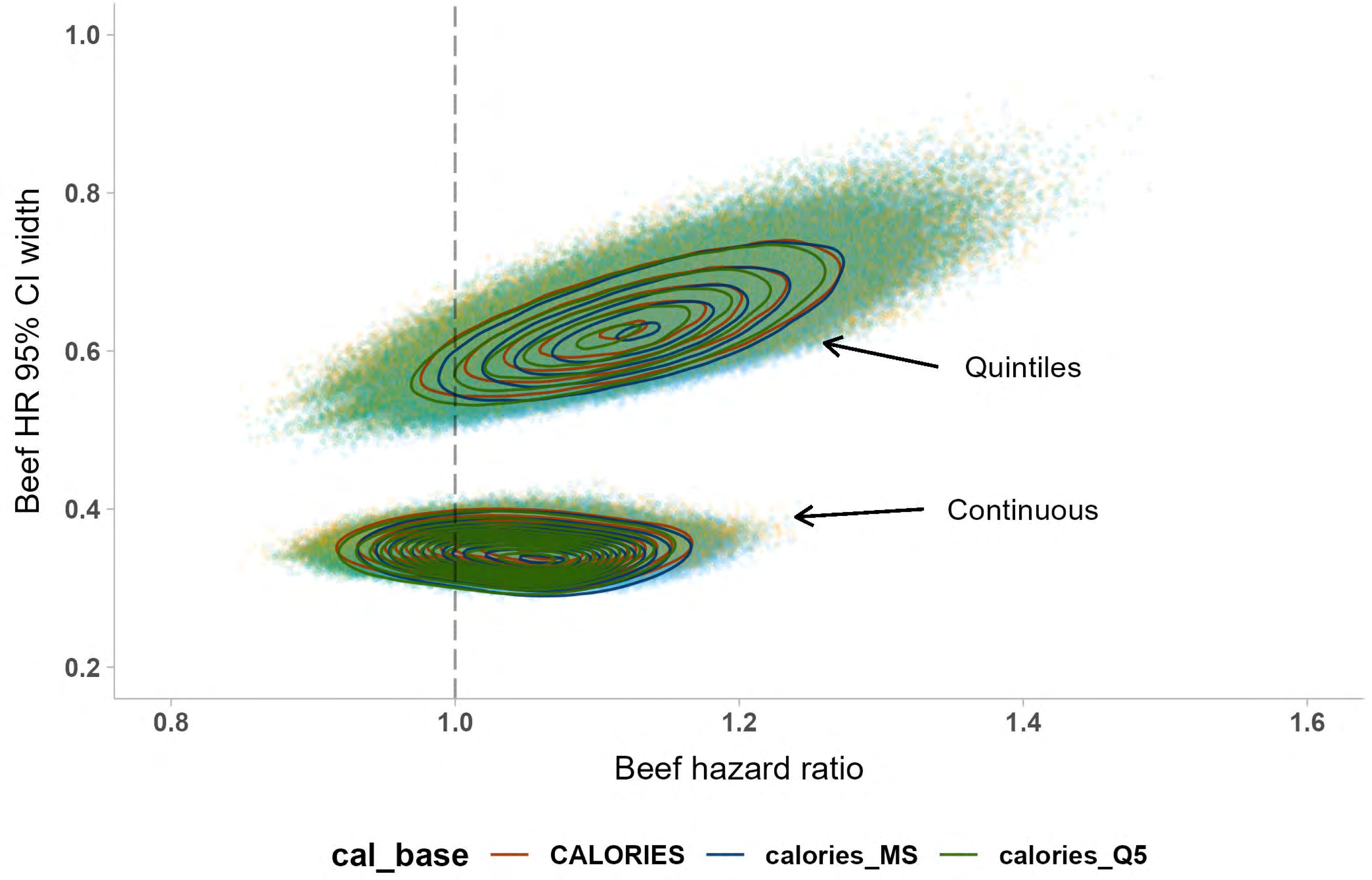

1.0
0.8
0.6
0.4
0.2
Beef HR 95% CI width
Quintiles
Continuous
0.8
1.0
1.2
1.4
1.6
Beef hazard ratio
cal_base
CALORIES
calories_MS
calories_Q5

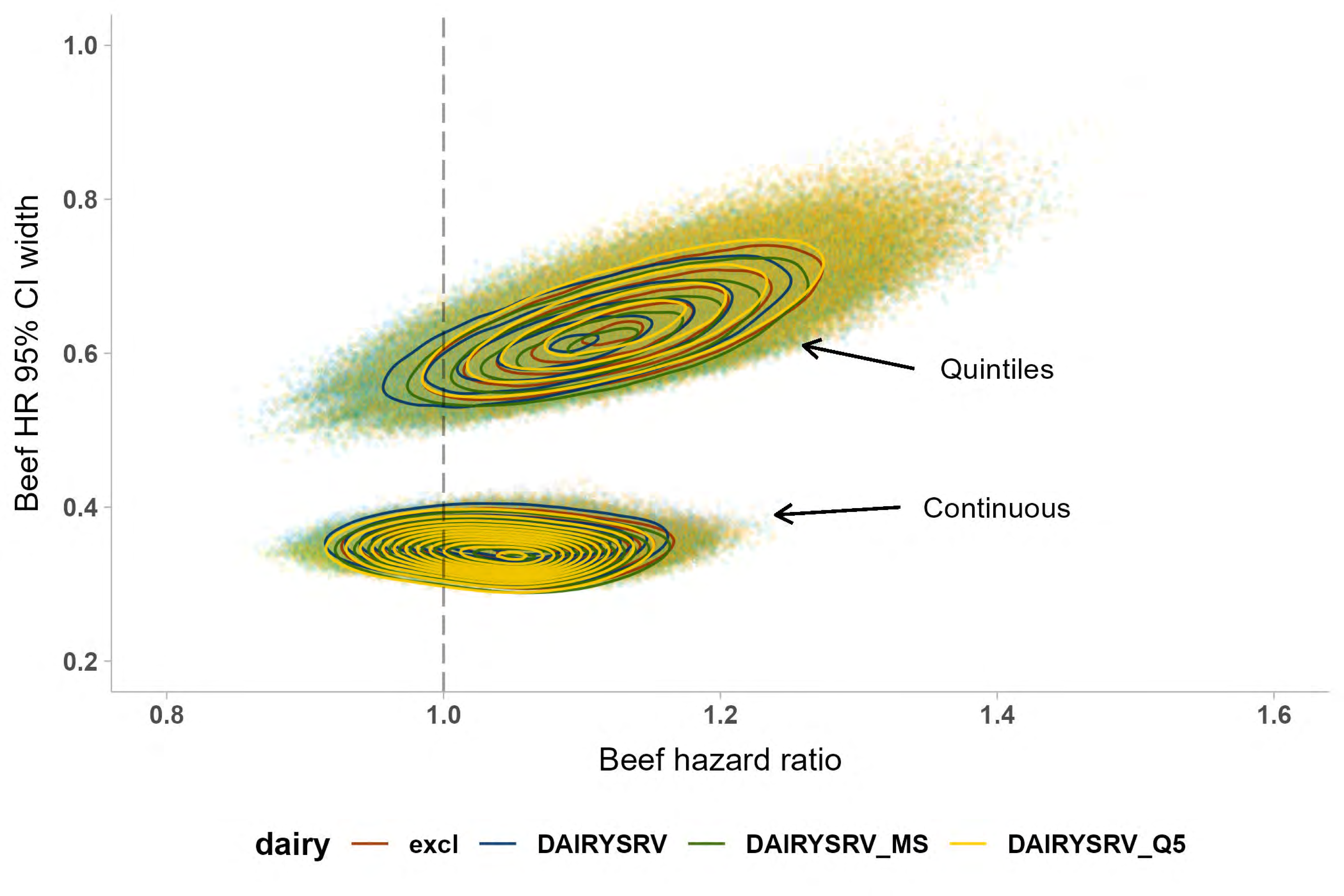
1.0
0.8
0.6
0.4
0.2
Beef HR 95% CI width
Quintiles
Continuous
0.8
1.0
1.2
1.4
1.6
Beef hazard ratio
dairy
excl
DAIRYSRV
DAIRYSRV_MS
DAIRYSRV_Q5

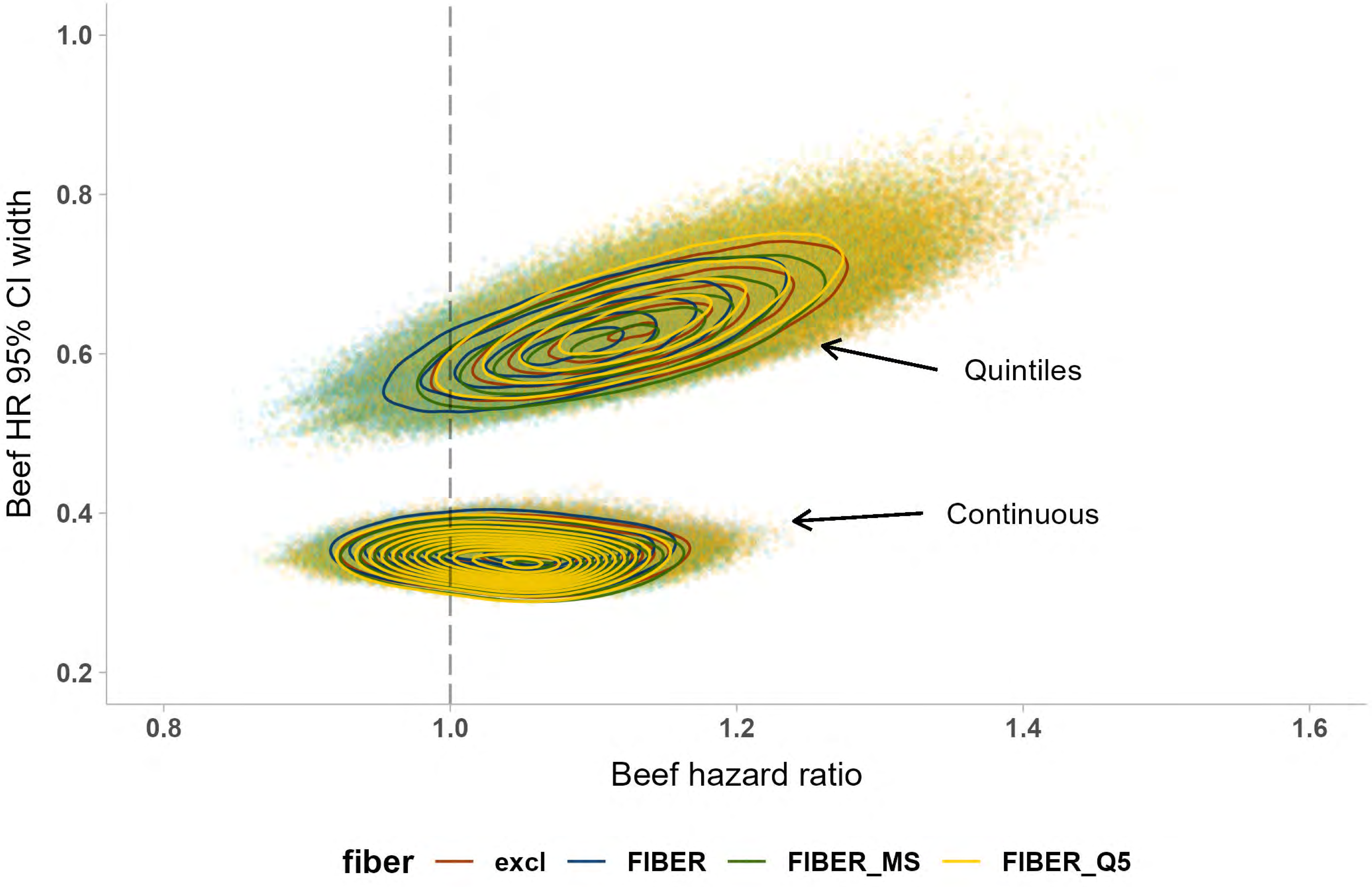

Beef HR 95% CI width
1.0
0.8
0.6
0.4
0.2
0.8
1.0
1.2
1.4
1.6
Beef hazard ratio
Quintiles
Continuous
fiber
excl
FIBER
FIBER_MS
FIBER_Q5

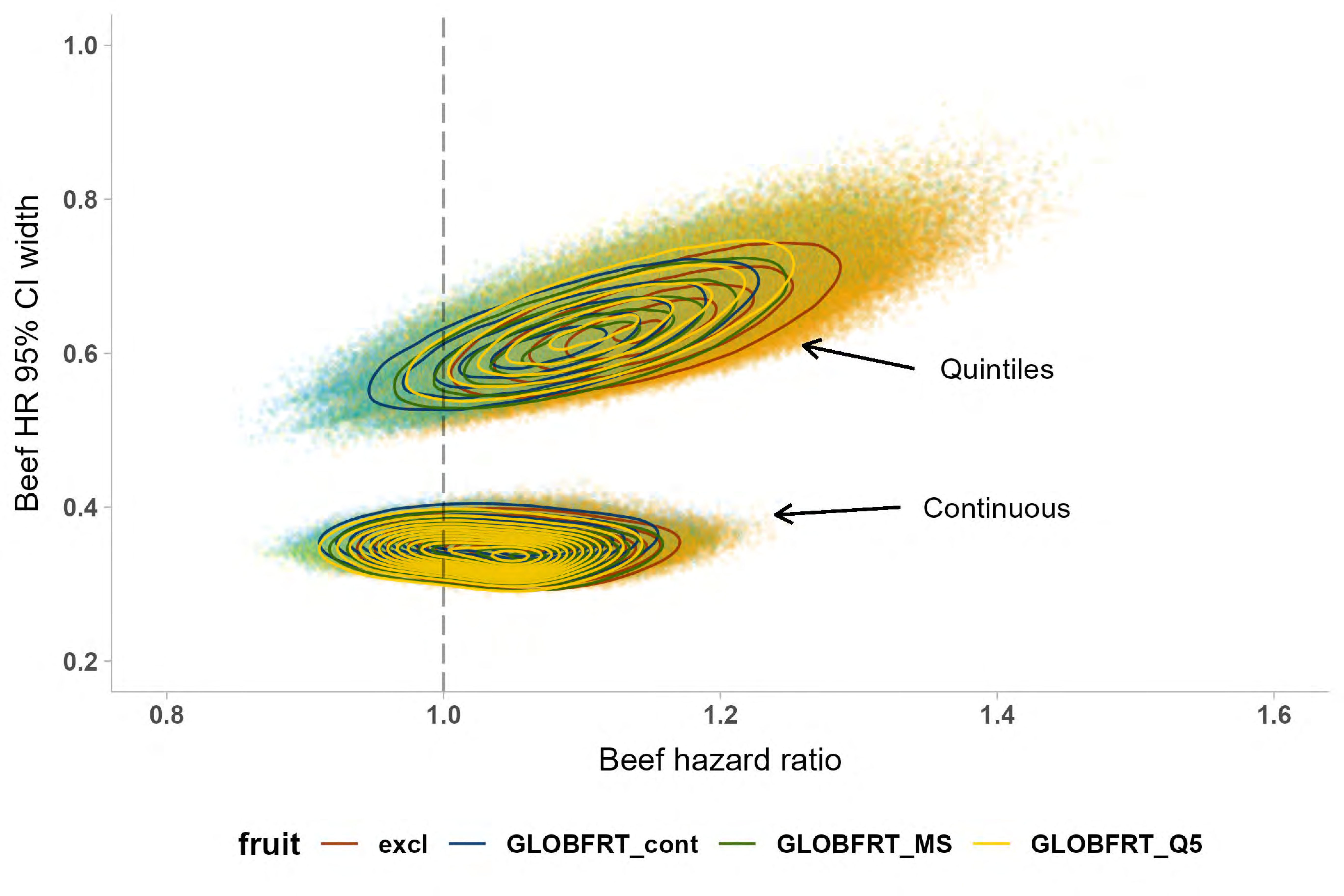

Beef HR 95% CI width
1.0
0.8
0.6
0.4
0.2
0.8
1.0
1.2
1.4
1.6
Beef hazard ratio
Quintiles
Continuous
fruit
excl
GLOBFRT_cont
GLOBFRT_MS
GLOBFRT_Q5

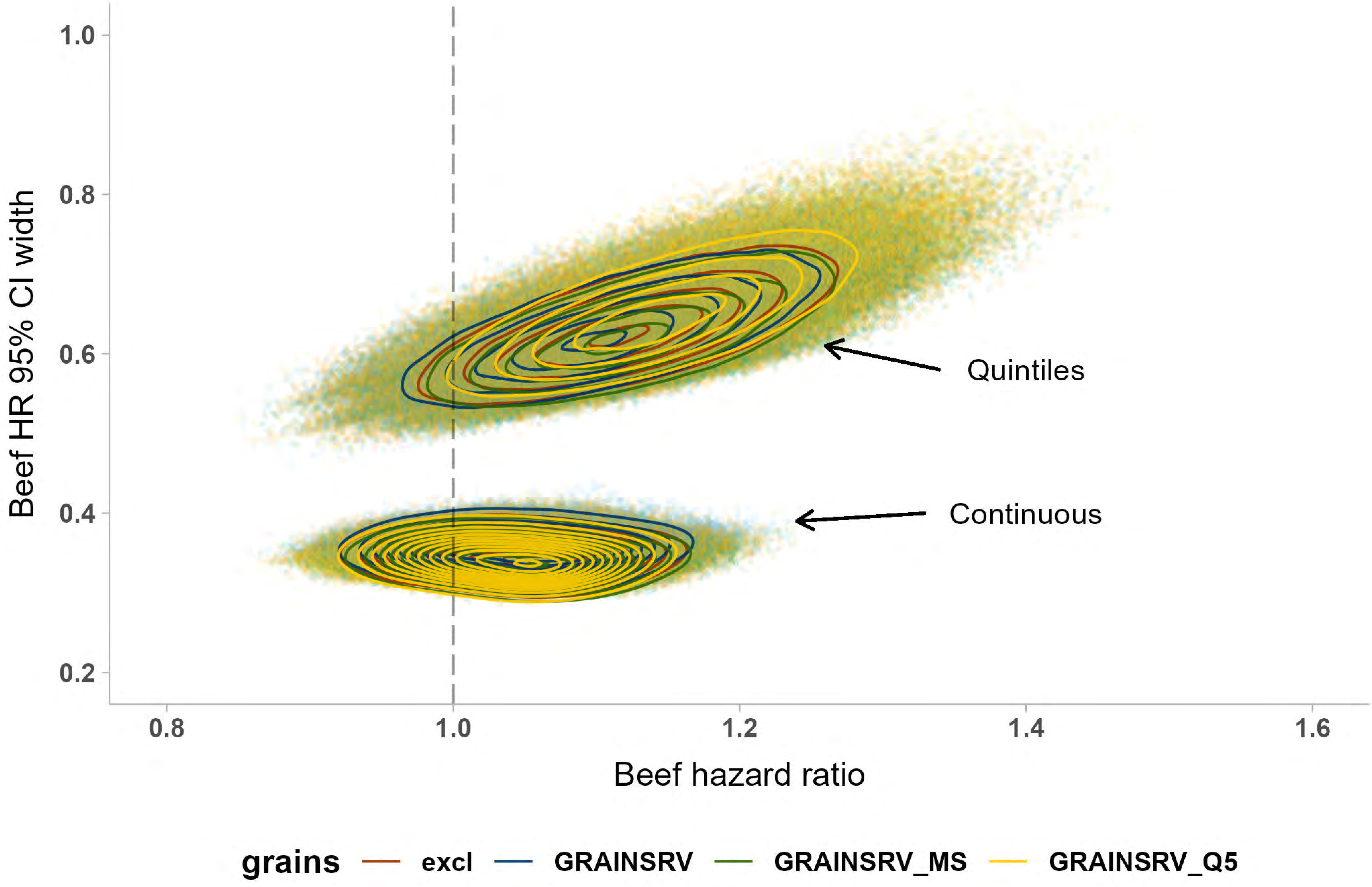

Beef HR 95% CI width
1.0
0.8
0.6
0.4
0.2
0.8
1.0
1.2
1.4
1.6
Beef hazard ratio
Quintiles
Continuous
grains
excl
GRAINSRV
GRAINSRV_MS
GRAINSRV_Q5

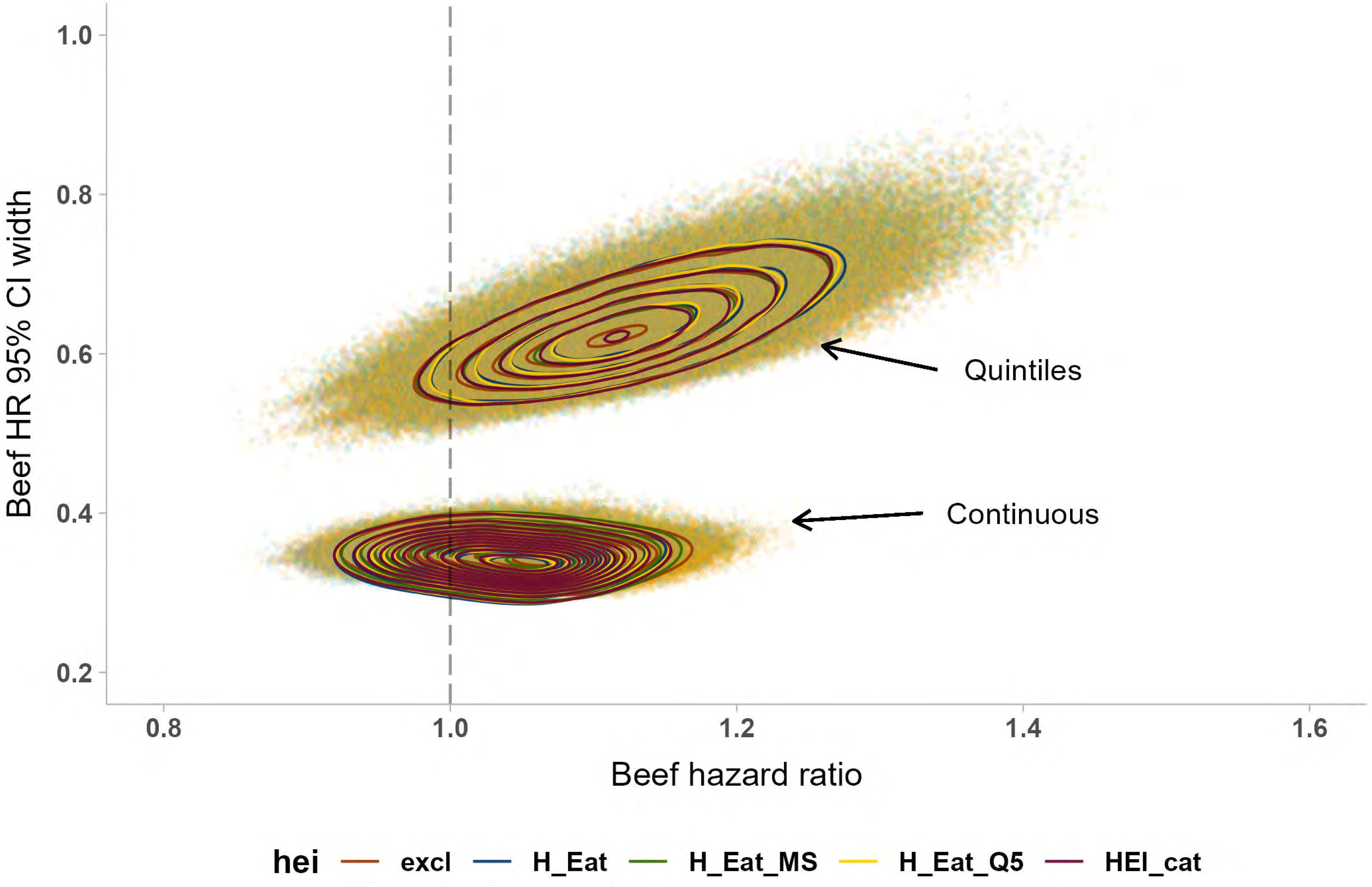
1.0
0.8
0.6
0.4
0.2
Beef HR 95% CI width
Quintiles
Continuous
0.8
1.0
1.2
1.4
1.6
Beef hazard ratio
hei
excl
H_Eat
H_Eat_MS
H_Eat_Q5
HEI_cat

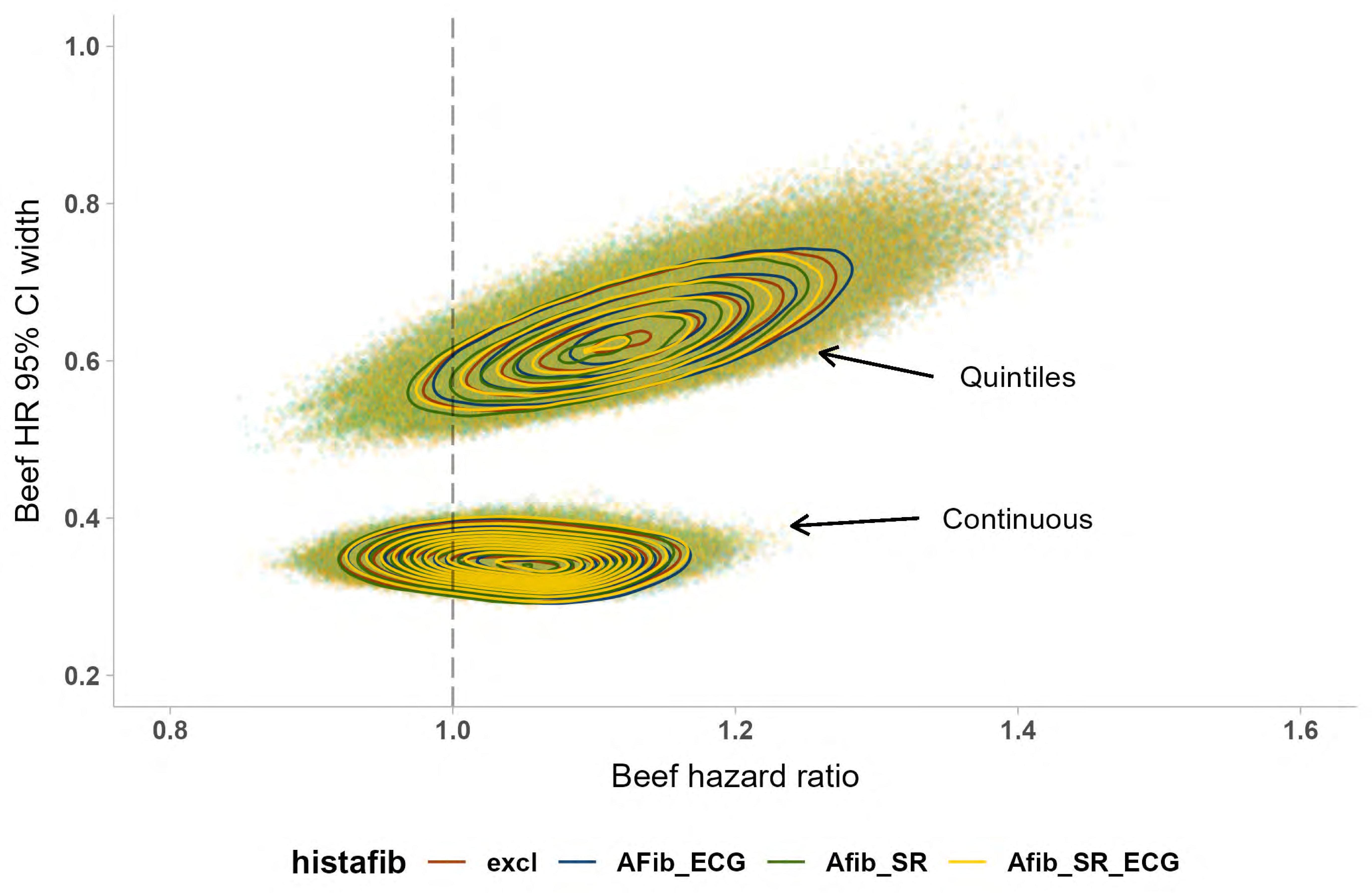

Beef HR 95% CI width
1.0
0.8
0.6
0.4
0.2
Quintiles
Continuous
0.8
1.0
1.2
1.4
1.6
Beef hazard ratio
histafib
excl
AFib_ECG
Afib_SR
Afib_SR_ECG

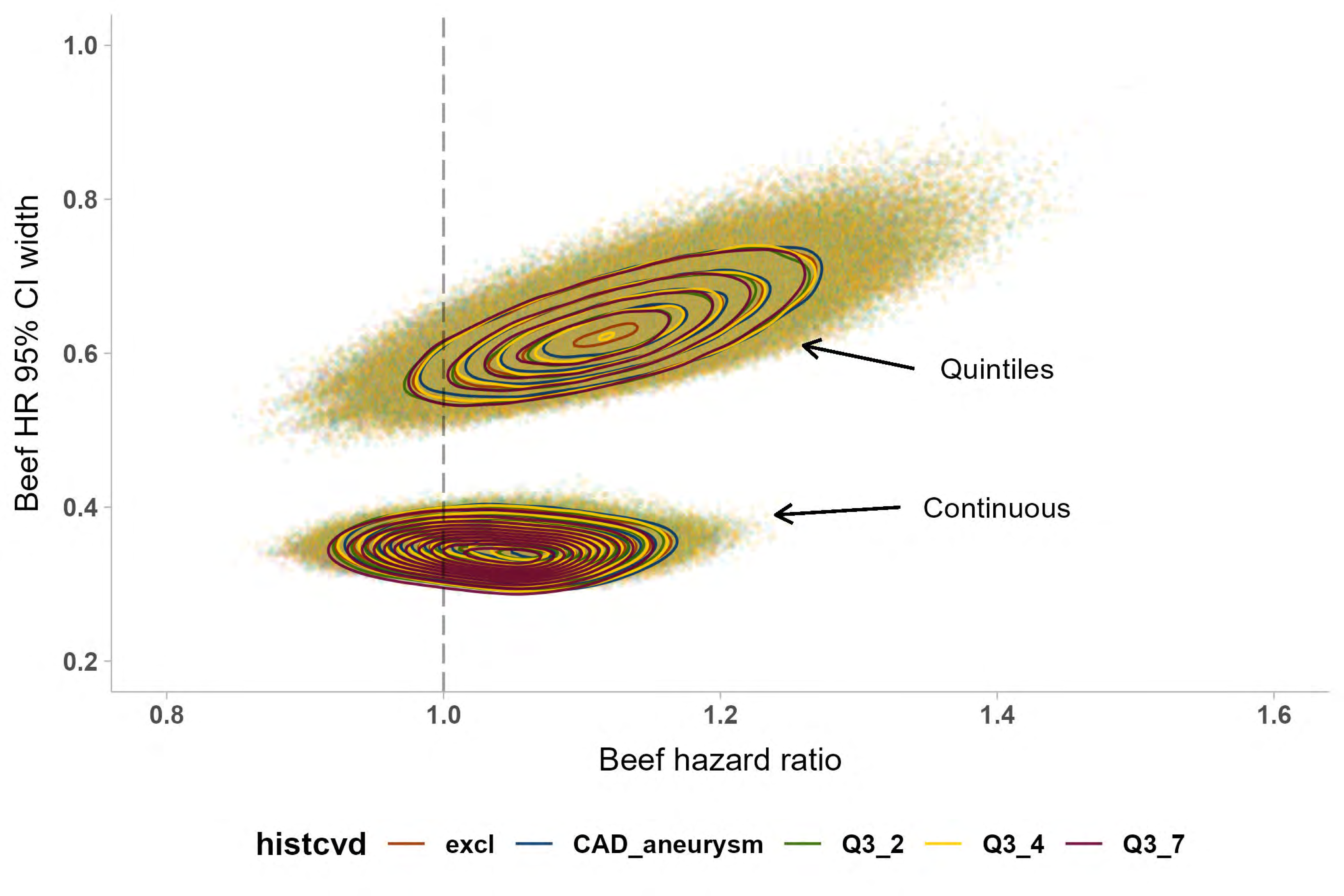

Beef HR 95% CI width
1.0
0.8
0.6
0.4
0.2
Quintiles
Continuous
0.8
1.0
1.2
1.4
1.6
Beef hazard ratio
histcvd
excl
CAD_aneurysm
Q3_2
Q3_4
Q3_7

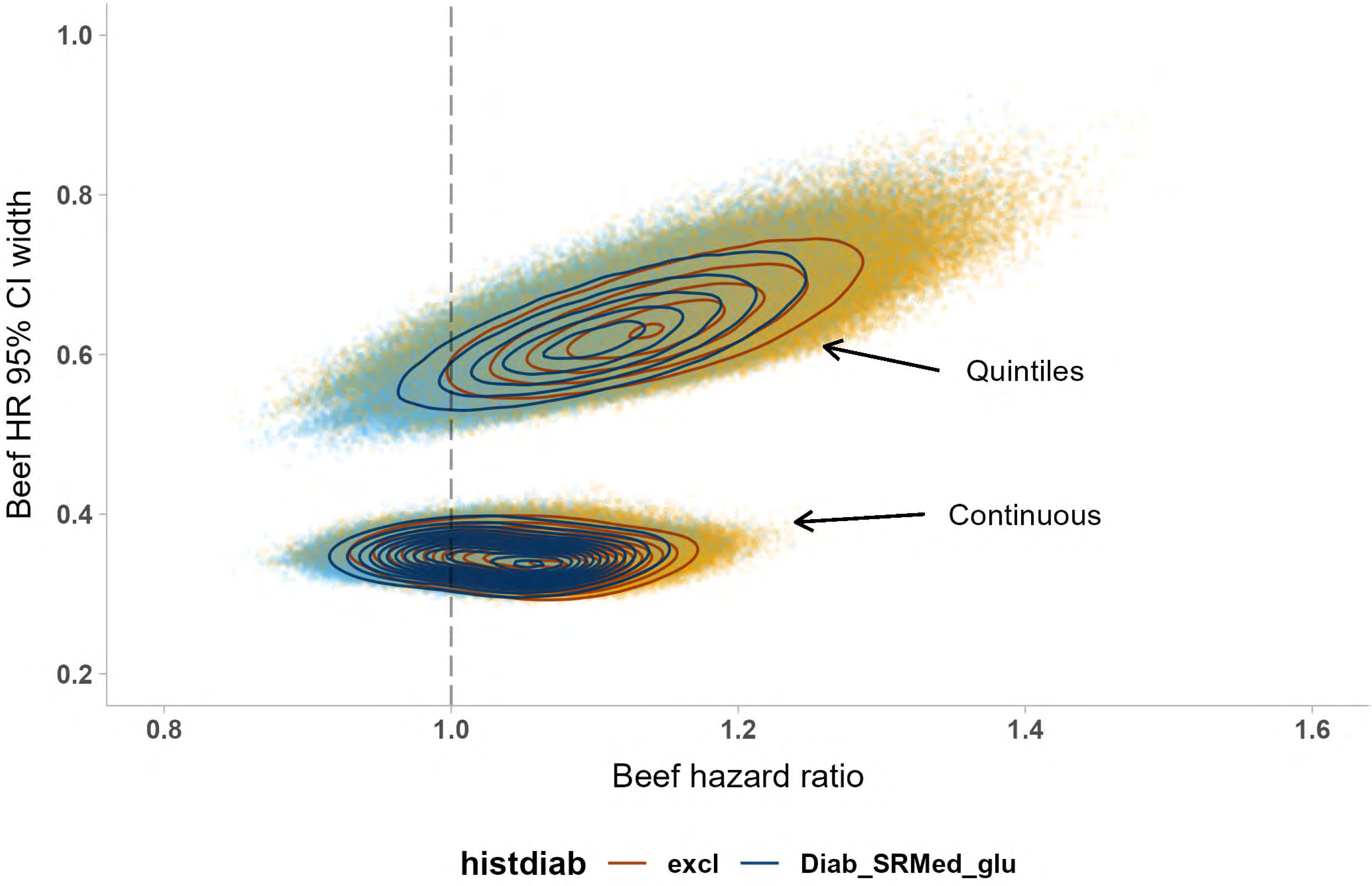

Beef HR 95% CI width
1.0
0.8
0.6
0.4
0.2
Quintiles
Continuous
0.8
1.0
1.2
1.4
1.6
Beef hazard ratio
histdiab
excl
Diab_SRMed_glu

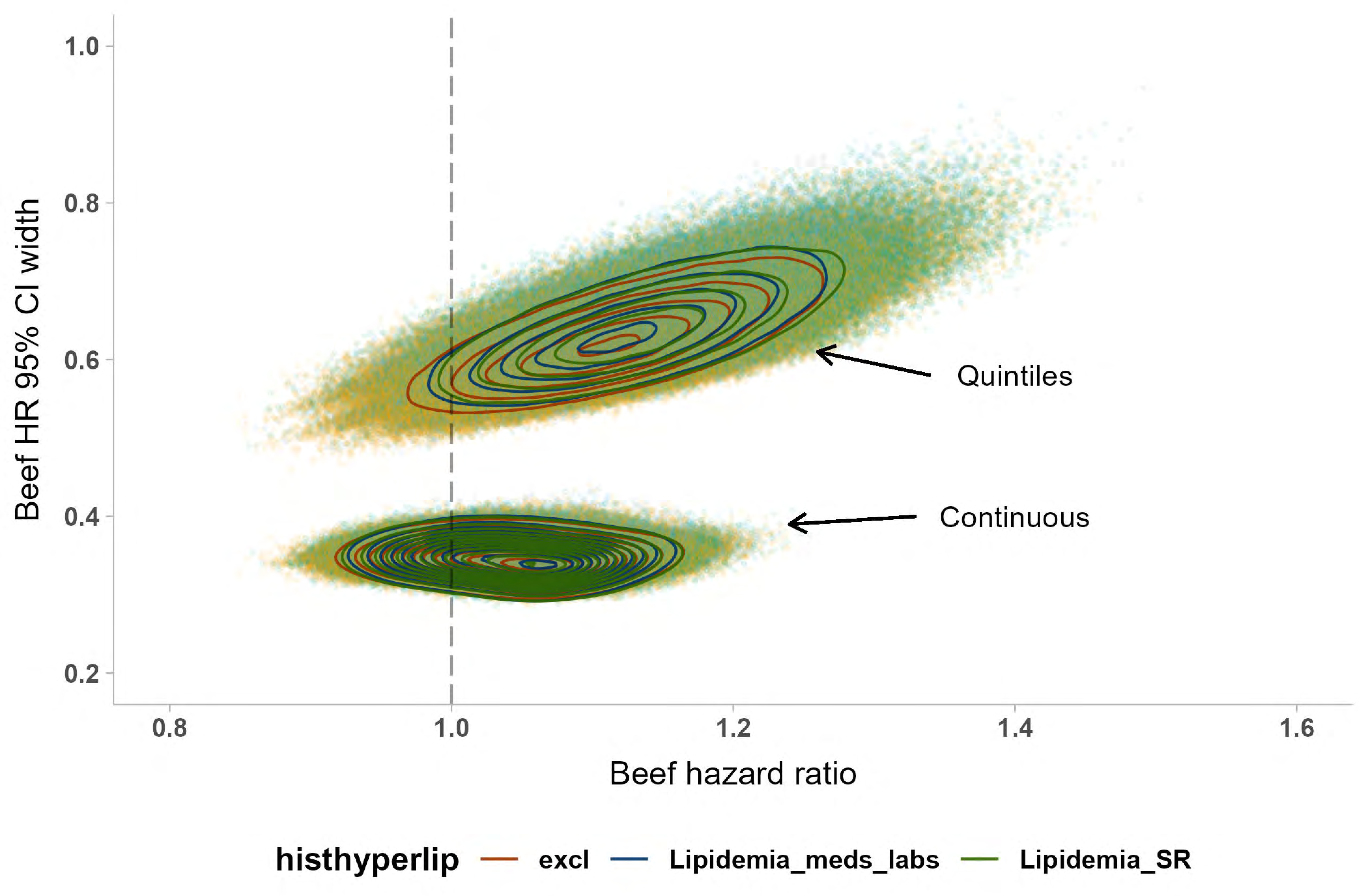

Beef HR 95% CI width
1.0
0.8
0.6
0.4
0.2
0.8
1.0
1.2
1.4
1.6
Beef hazard ratio
Quintiles
Continuous
histhyperlip
excl
Lipidemia_meds_labs
Lipidemia_SR

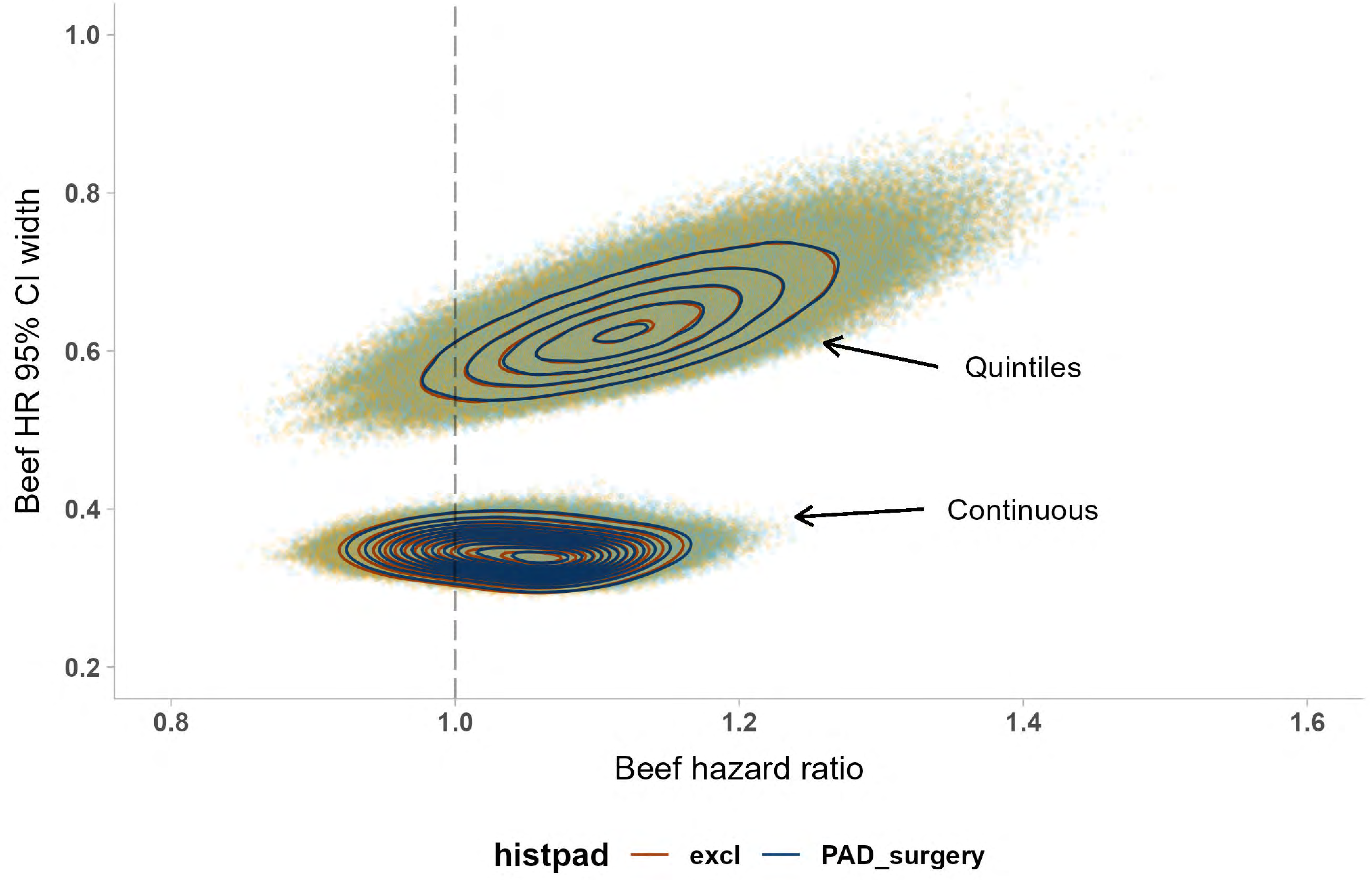

1.0
0.8
0.6
0.4
0.2
Beef HR 95% CI width
0.8
1.0
1.2
1.4
1.6
Beef hazard ratio
Quintiles
Continuous
histpad
excl
PAD_surgery

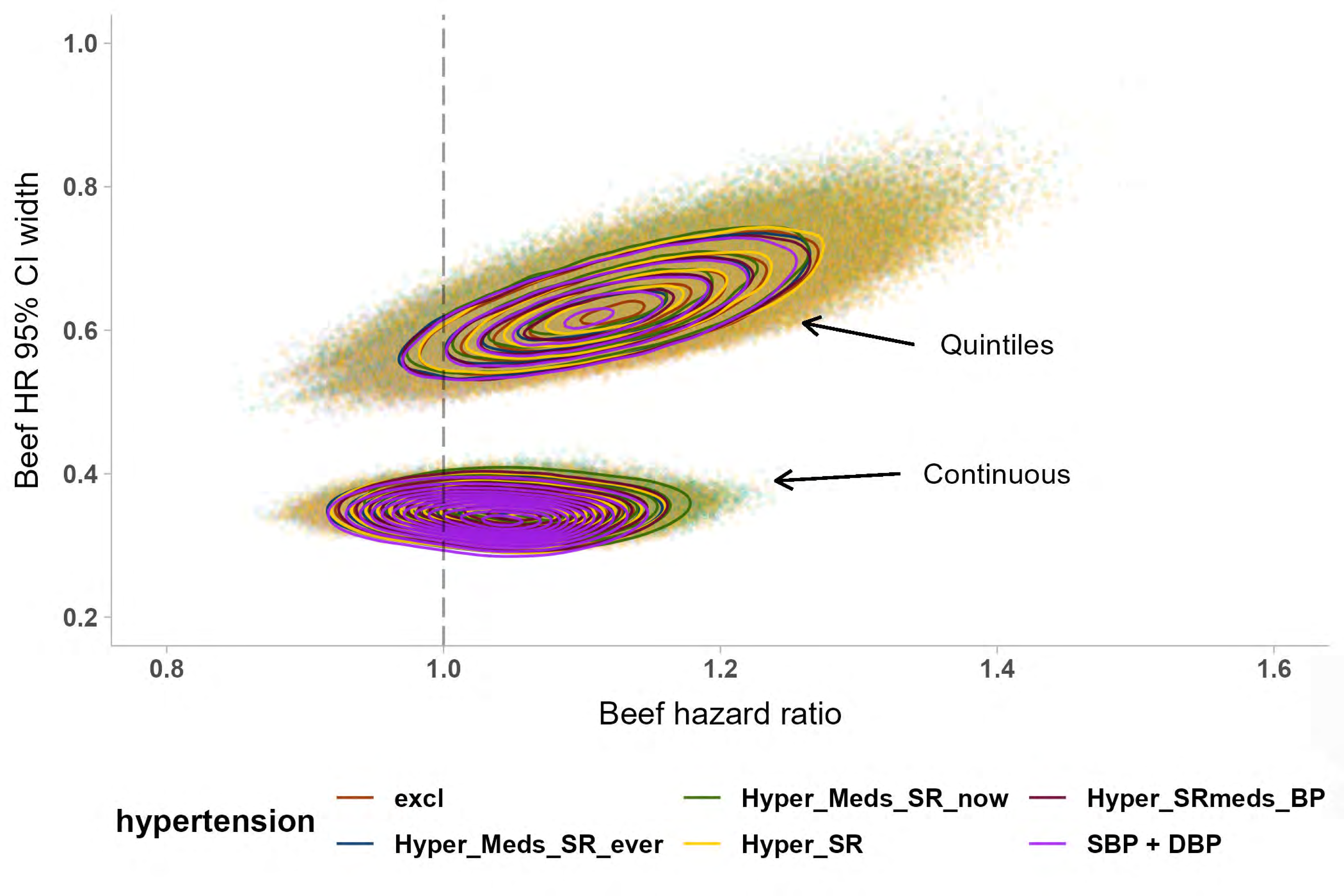

1.0
0.8
0.6
0.4
0.2
Beef HR 95% CI width
0.8
1.0
1.2
1.4
1.6
Beef hazard ratio
Quintiles
Continuous
hypertension
excl
Hyper_Meds_SR_ever
Hyper_Meds_SR_now
Hyper_SR
Hyper_SRmeds_BP
SBP + DBP

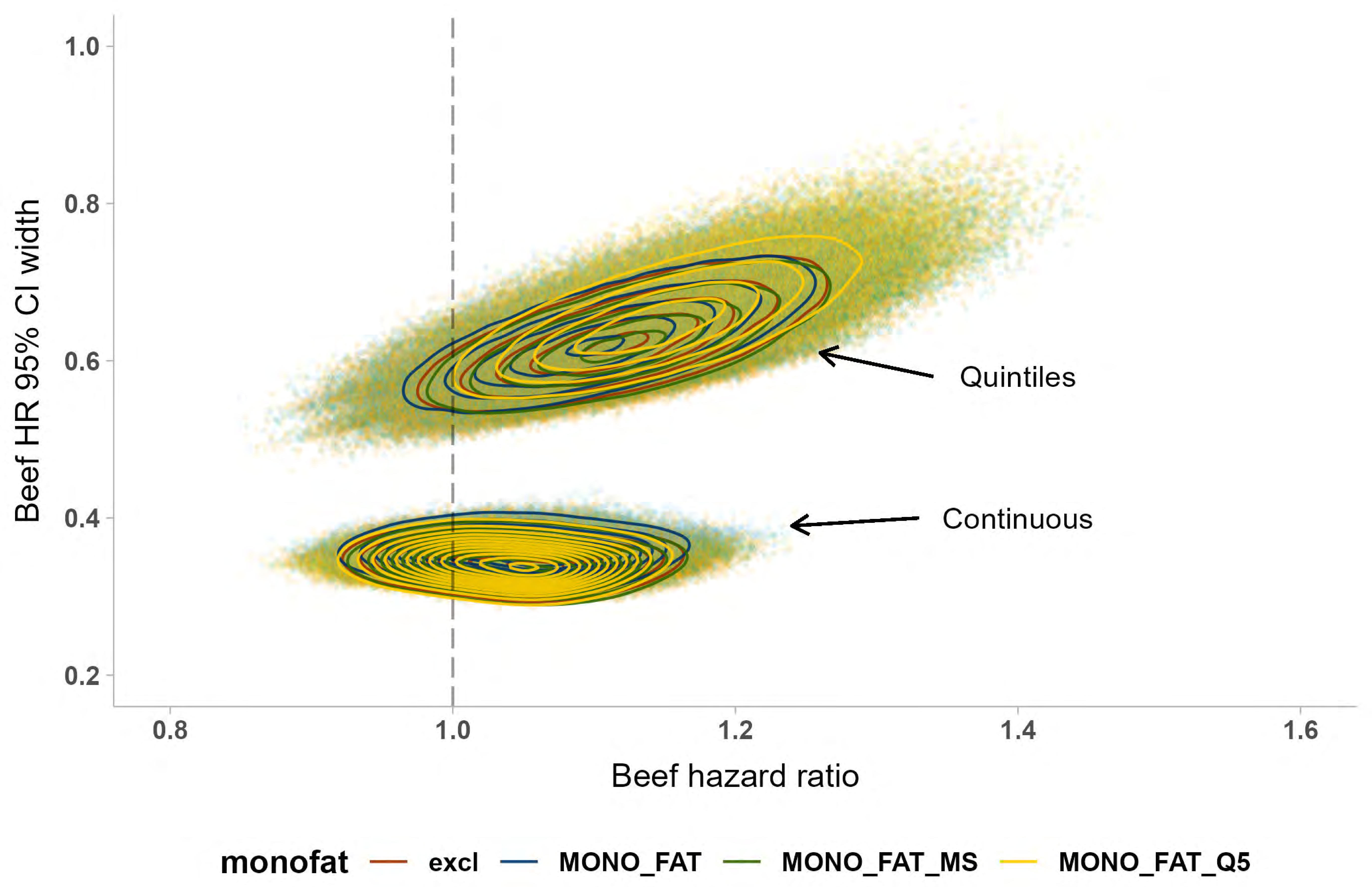

Beef HR 95% CI width
1.0
0.8
0.6
0.4
0.2
0.8
1.0
1.2
1.4
1.6
Beef hazard ratio
Quintiles
Continuous
monofat
excl
MONO_FAT
MONO_FAT_MS
MONO_FAT_Q5

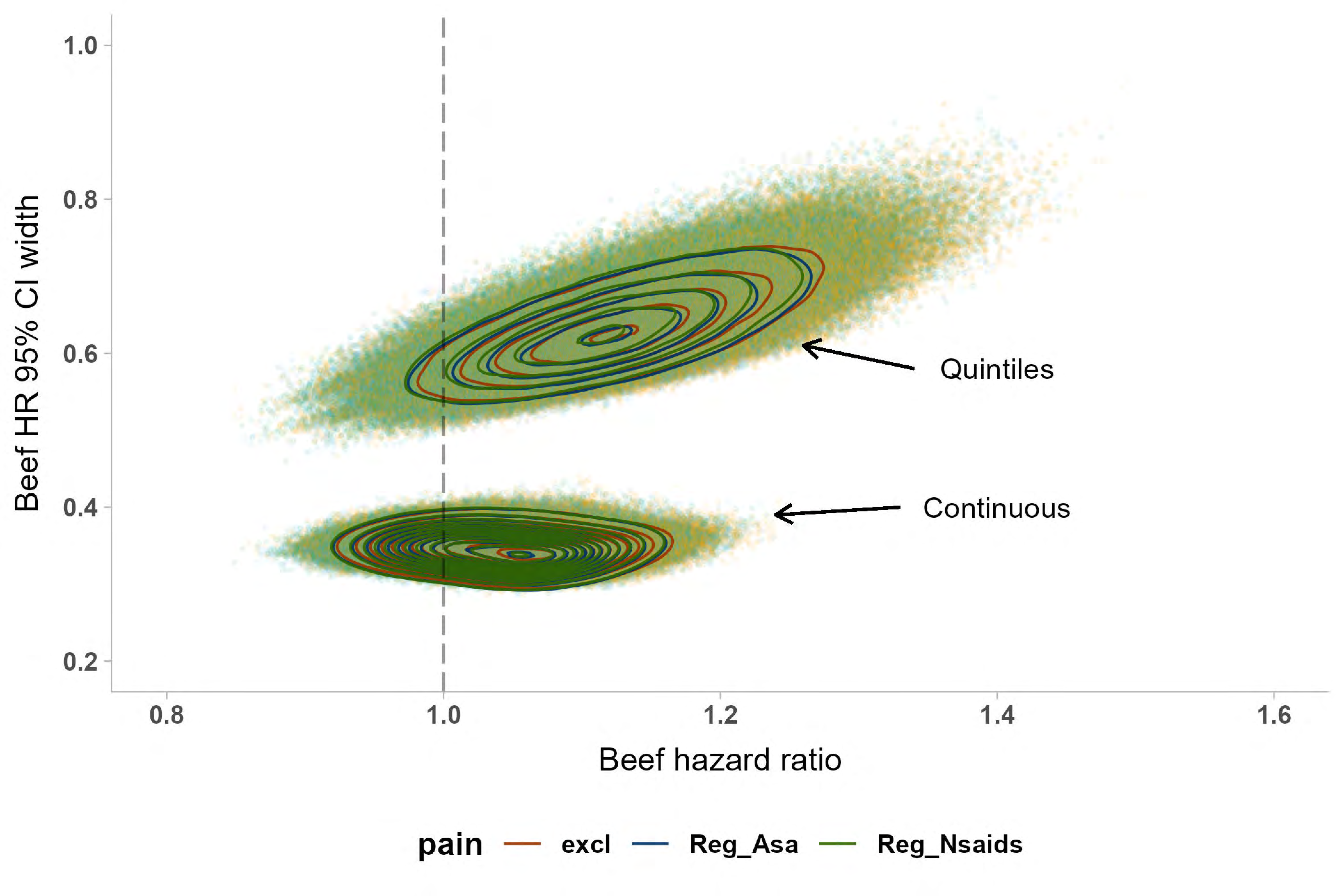

1.0
0.8
0.6
0.4
0.2
Beef HR 95% CI width
0.8
1.0
1.2
1.4
1.6
Beef hazard ratio
Quintiles
Continuous
pain
excl
Reg_Asa
Reg_Nsaids

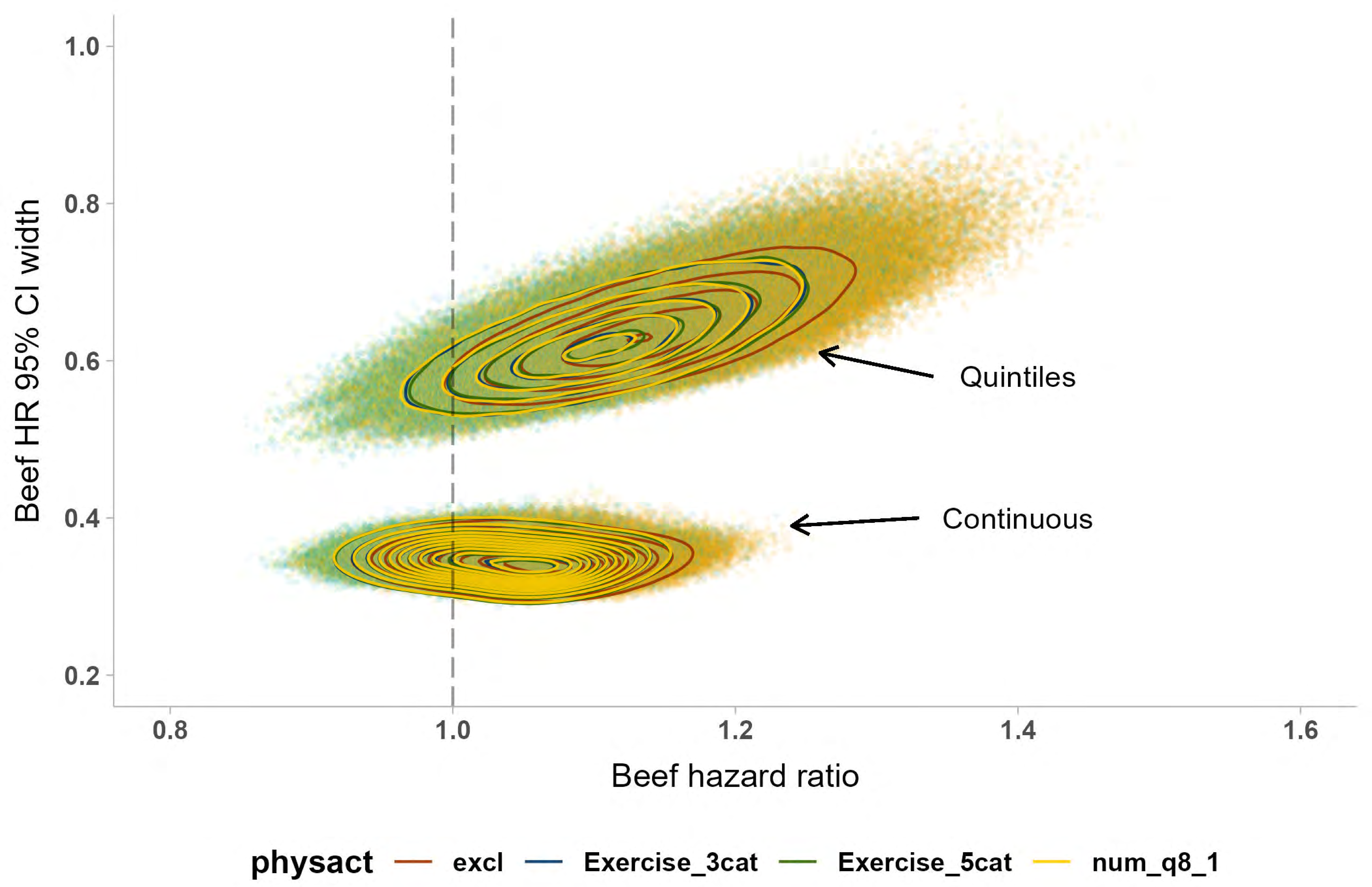
1.0
0.8
0.6
0.4
0.2
Beef HR 95% CI width
Quintiles
Continuous
0.8
1.0
1.2
1.4
1.6
Beef hazard ratio
physact
excl
Exercise_3cat
Exercise_5cat
num_q8_1

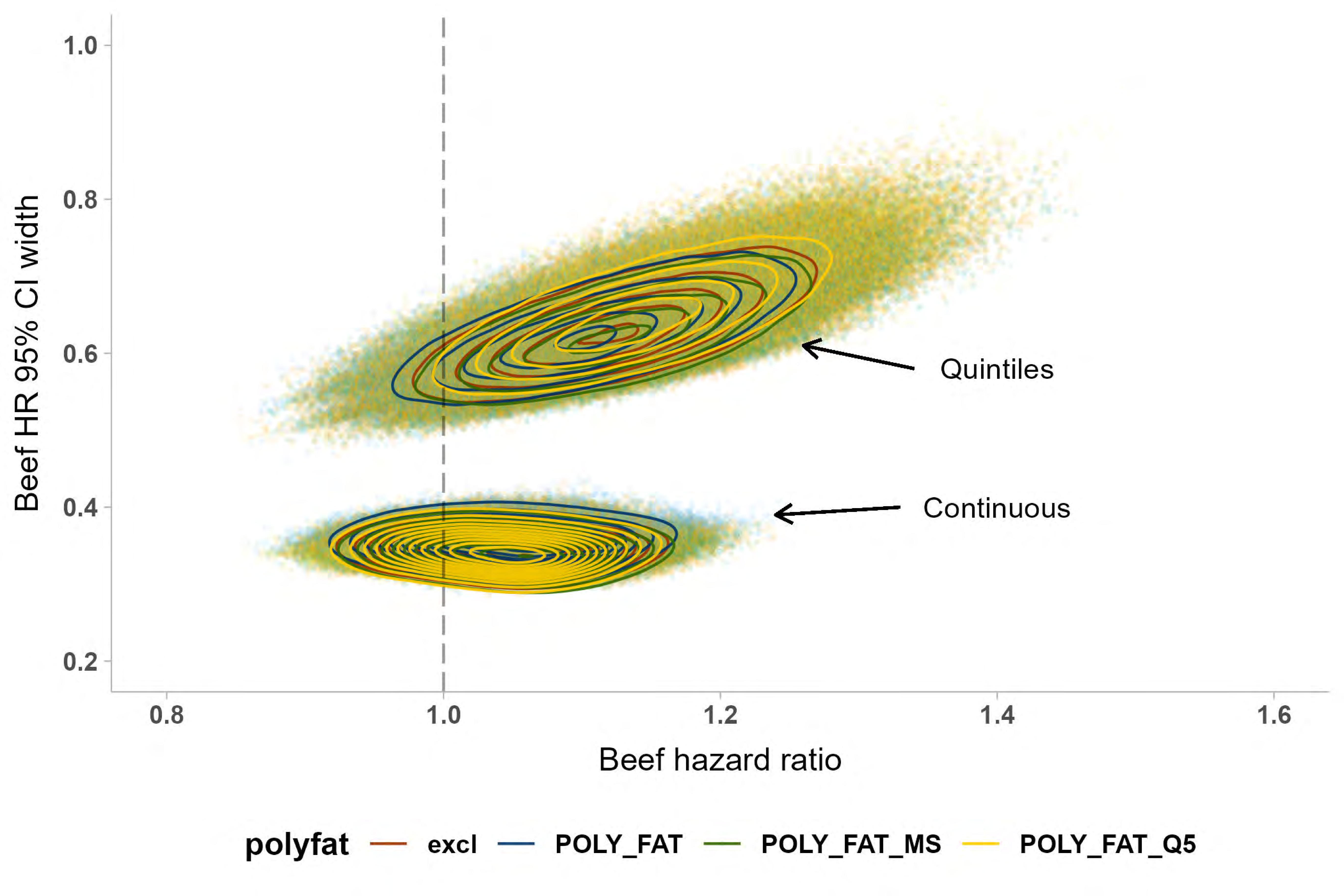

Beef HR 95% CI width
1.0
0.8
0.6
0.4
0.2
0.8
1.0
1.2
1.4
1.6
Beef hazard ratio
Quintiles
Continuous
polyfat
excl
POLY_FAT
POLY_FAT_MS
POLY_FAT_Q5

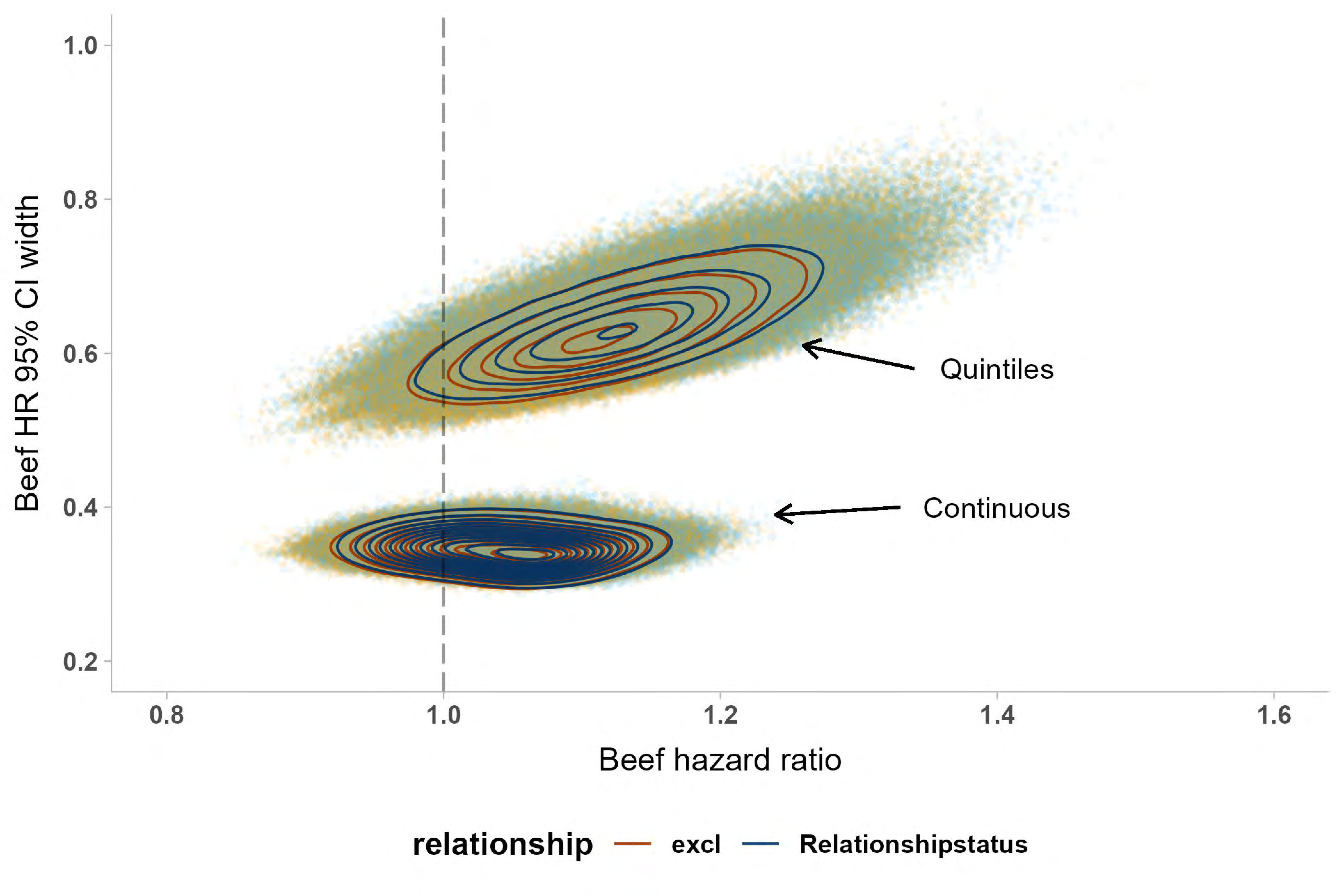

Beef HR 95% CI width
1.0
0.8
0.6
0.4
0.2
0.8
1.0
1.2
1.4
1.6
Beef hazard ratio
Quintiles
Continuous
relationship
excl
Relationshipstatus

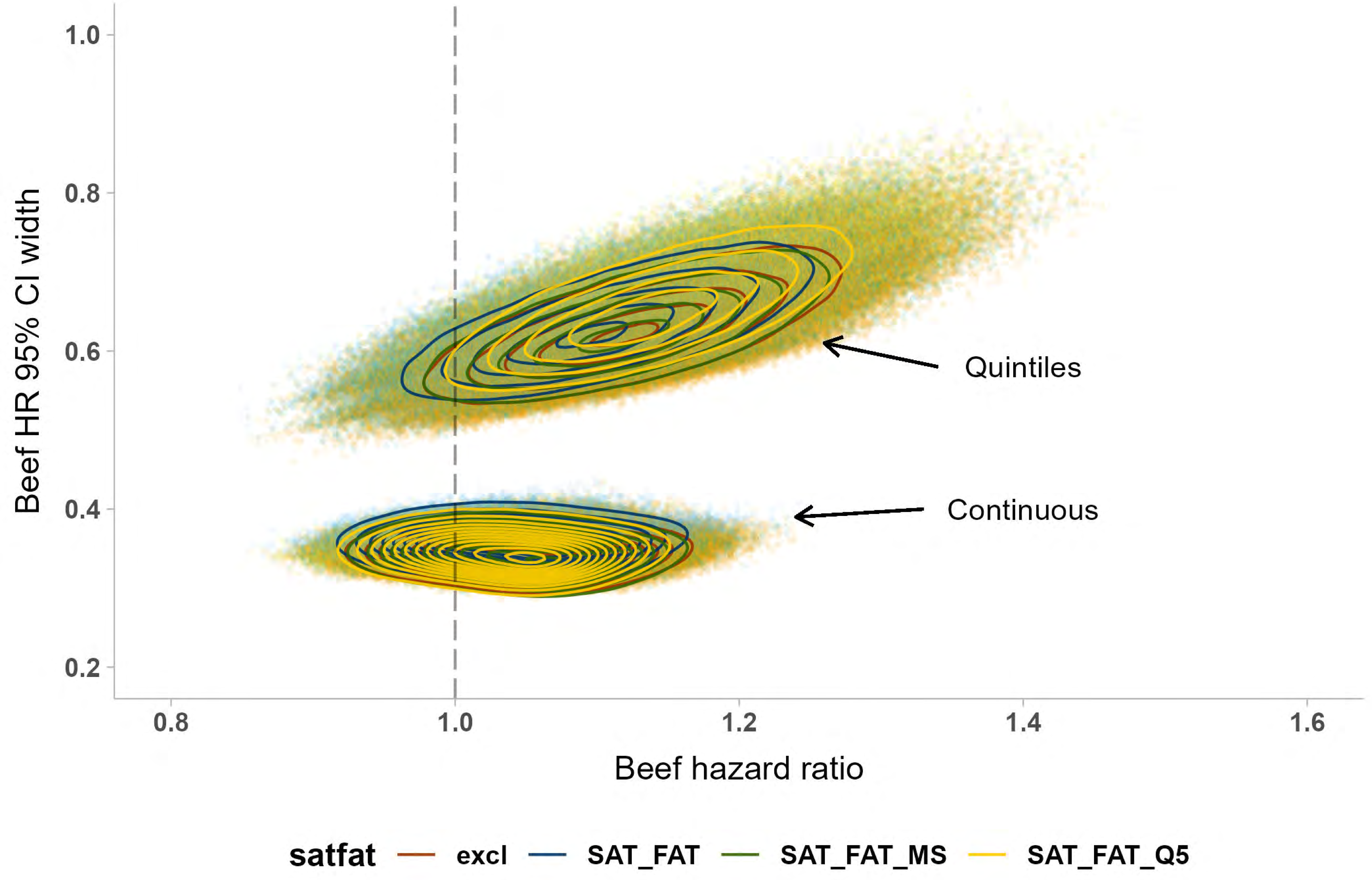

Beef HR 95% CI width
1.0
0.8
0.6
0.4
0.2
0.8
1.0
1.2
1.4
1.6
Beef hazard ratio
Quintiles
Continuous
satfat
excl
SAT_FAT
SAT_FAT_MS
SAT_FAT_Q5

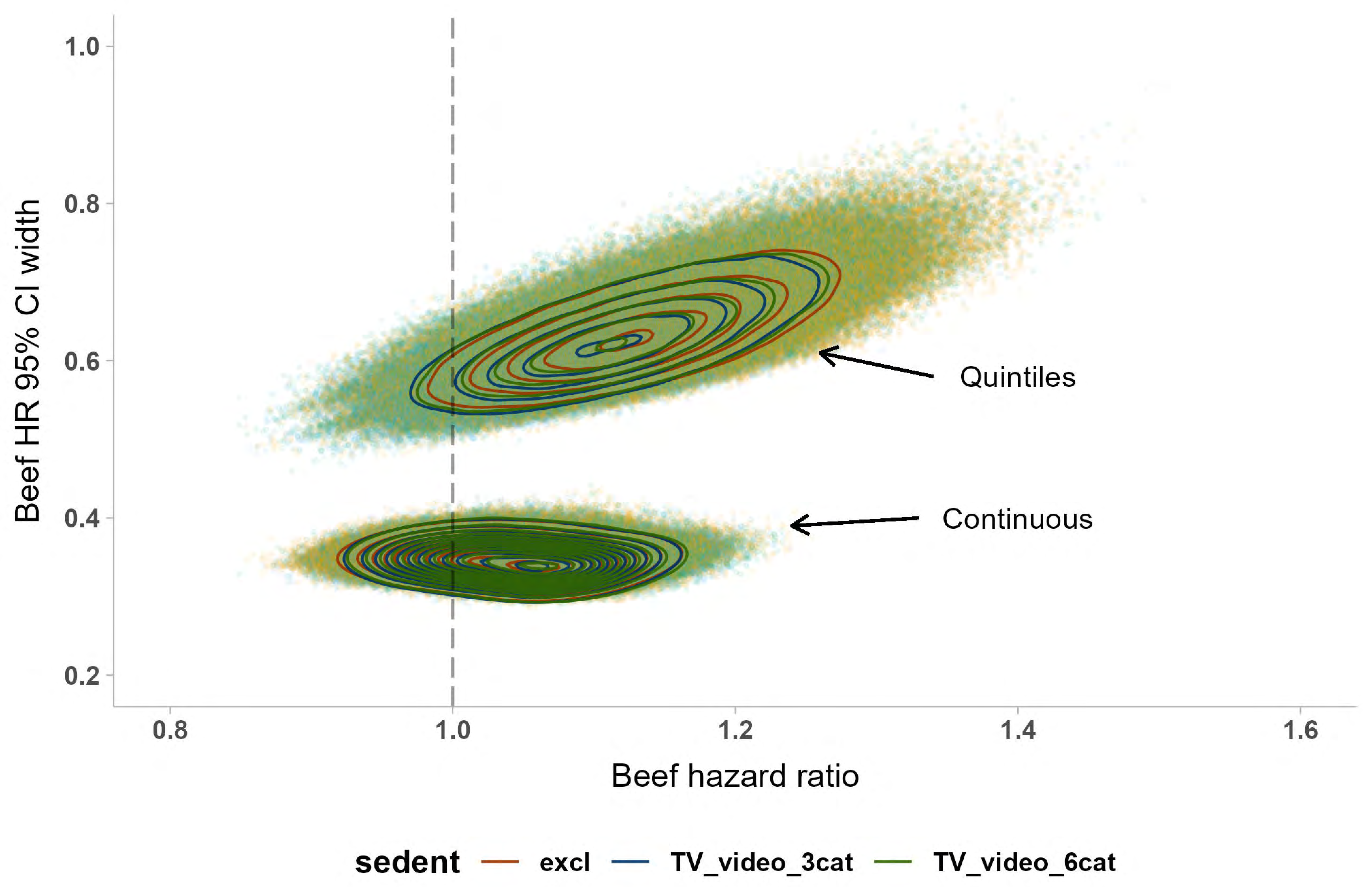

1.0
0.8
0.6
0.4
0.2
Beef HR 95% CI width
0.8
1.0
1.2
1.4
1.6
Beef hazard ratio
Quintiles
Continuous
sedent
excl
TV_video_3cat
TV_video_6cat

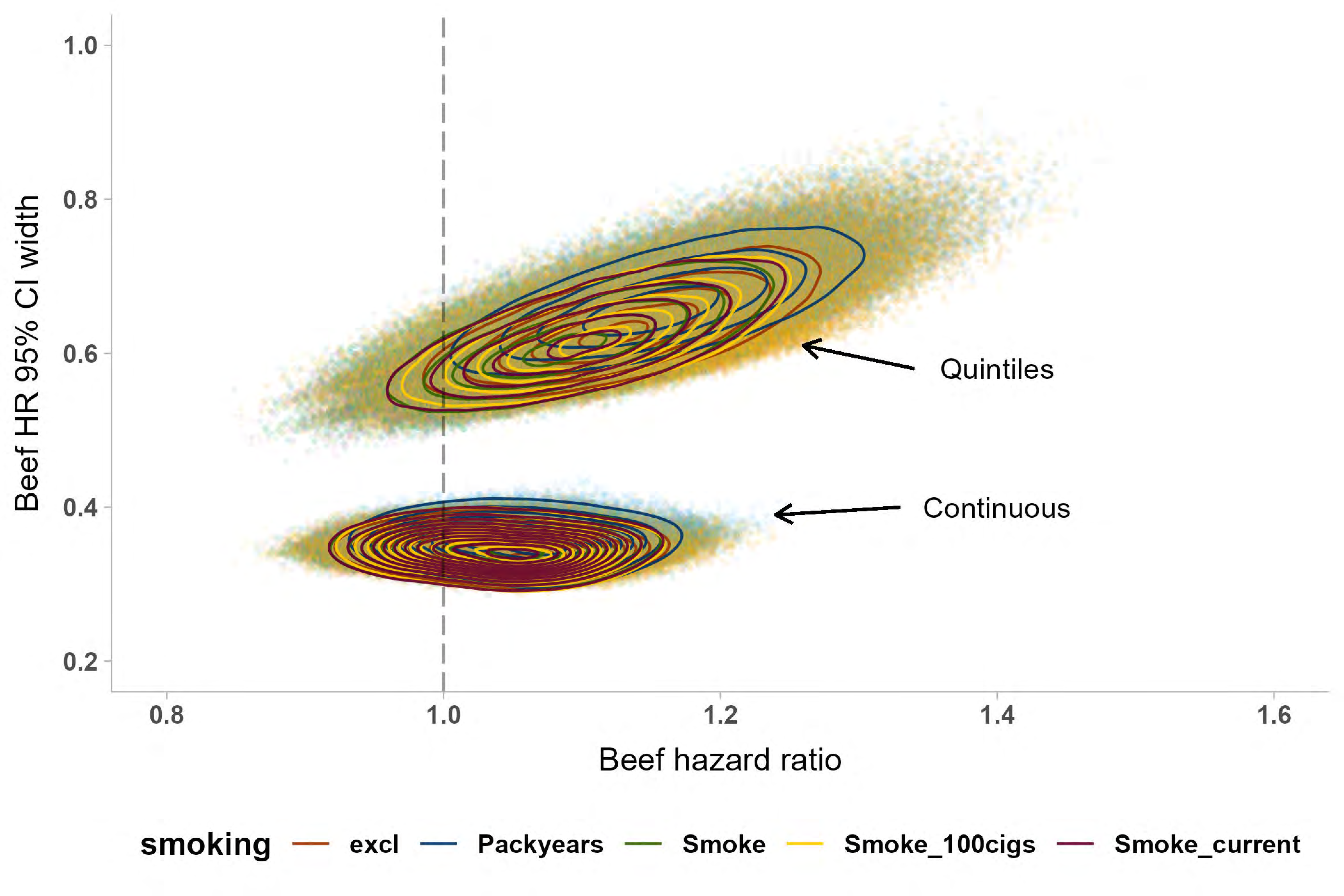

Beef HR 95% CI width
1.0
0.8
0.6
0.4
0.2
Beef hazard ratio
0.8
1.0
1.2
1.4
1.6
Quintiles
Continuous
smoking
excl
Packyears
Smoke
Smoke_100cigs
Smoke_current

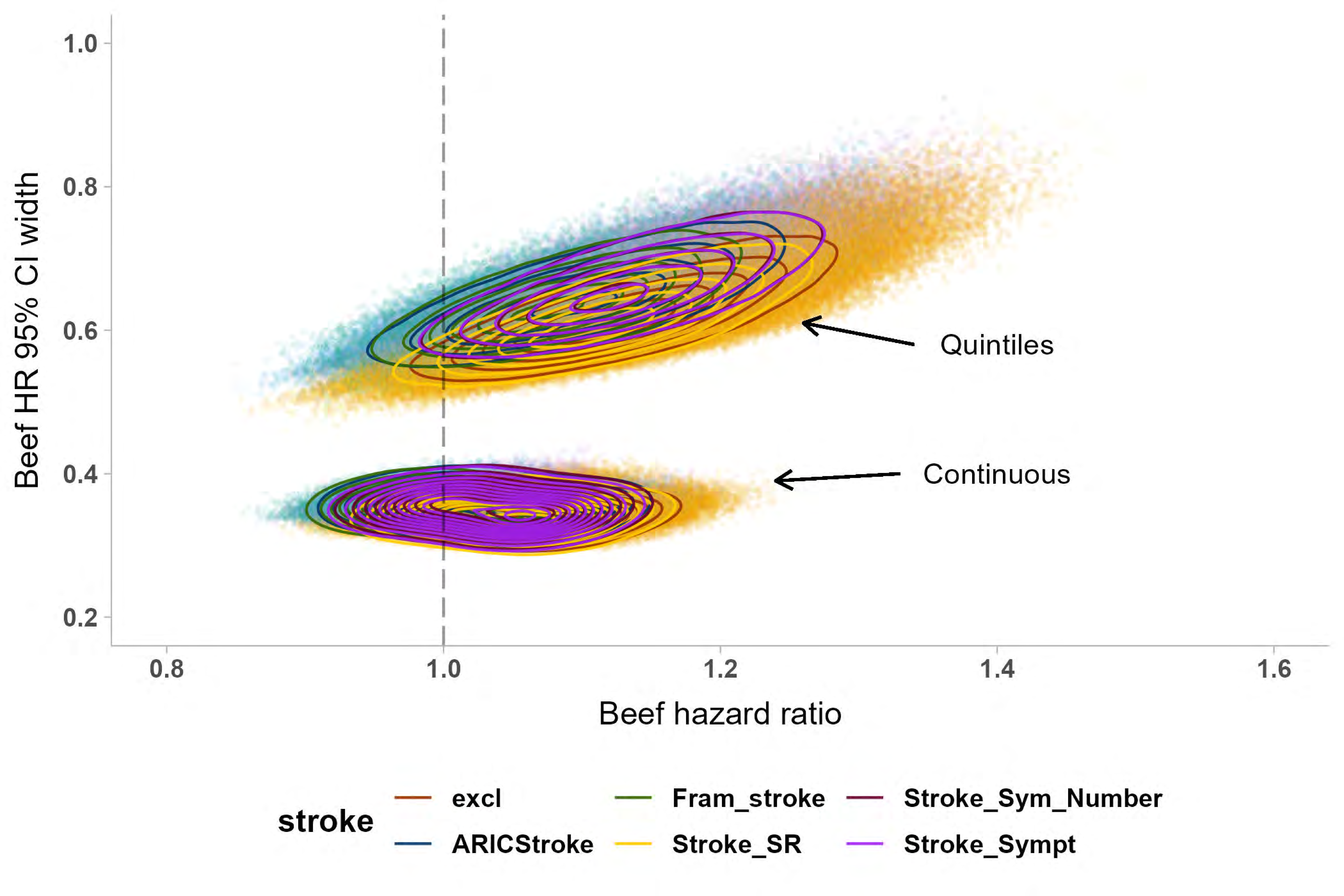
1.0
0.8
0.6
0.4
0.2
Beef HR 95% CI width
0.8
1.0
1.2
1.4
1.6
Beef hazard ratio
Quintiles
Continuous
stroke
excl
ARICStroke
Fram_stroke
Stroke_SR
Stroke_Sym_Number
Stroke_Sympt

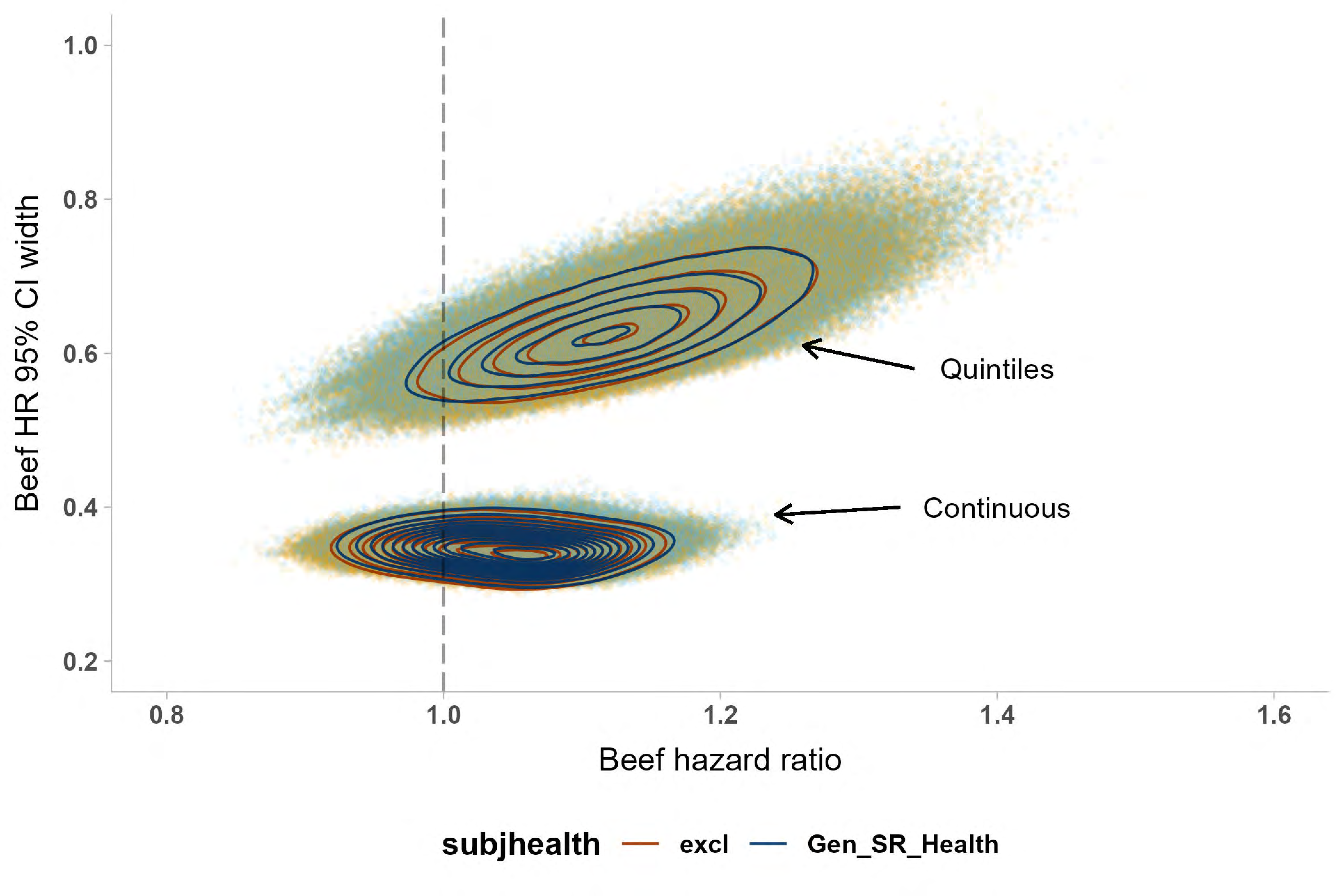

Beef HR 95% CI width
1.0
0.8
0.6
0.4
0.2
Quintiles
Continuous
0.8
1.0
1.2
1.4
1.6
Beef hazard ratio
subjhealth
excl
Gen_SR_Health

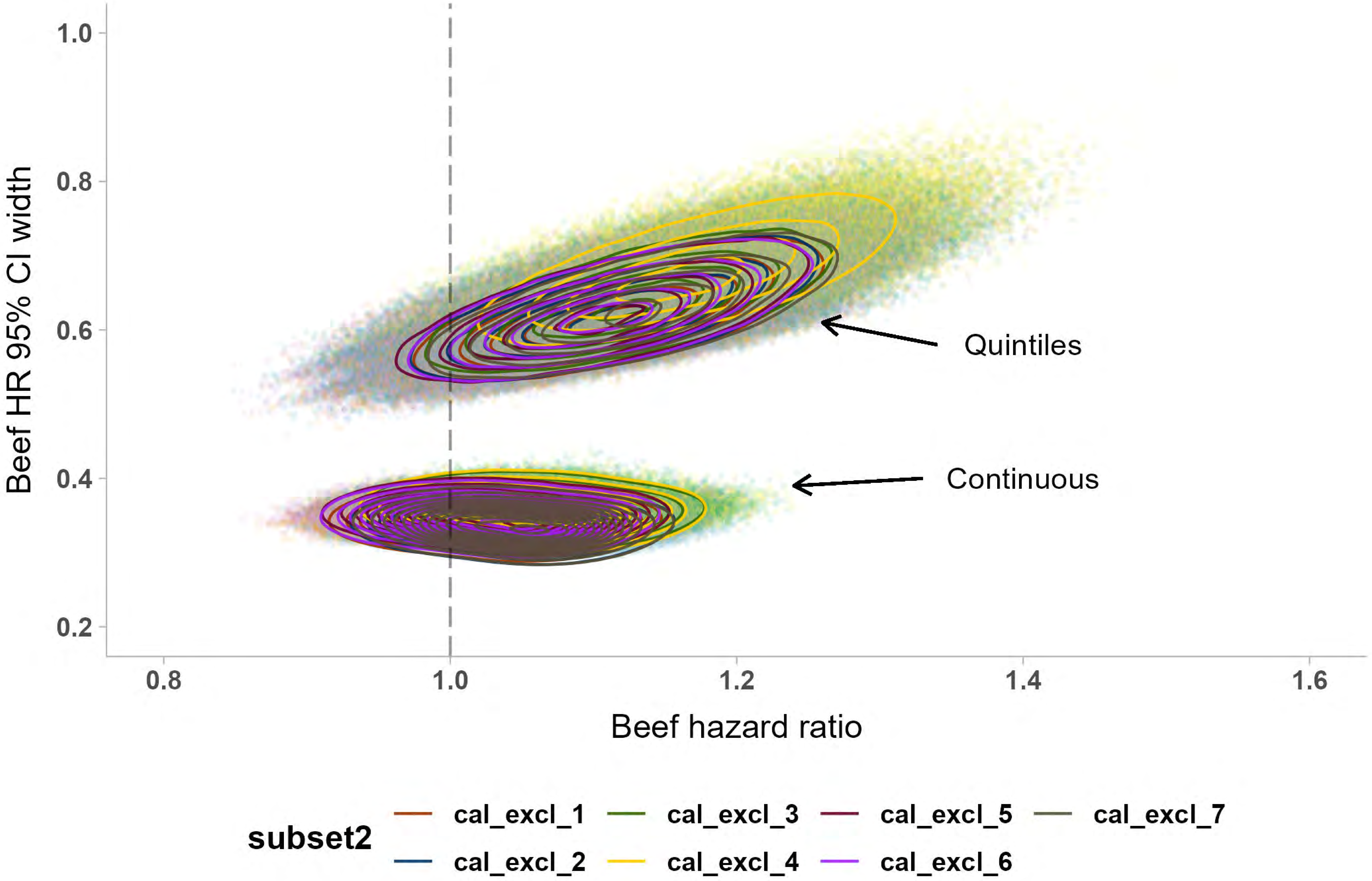

Beef HR 95% CI width
1.0
0.8
0.6
0.4
0.2
0.8
1.0
1.2
1.4
1.6
Beef hazard ratio
Quintiles
Continuous
subset2
cal_excl_1
cal_excl_2
cal_excl_3
cal_excl_4
cal_excl_5
cal_excl_6
cal_excl_7

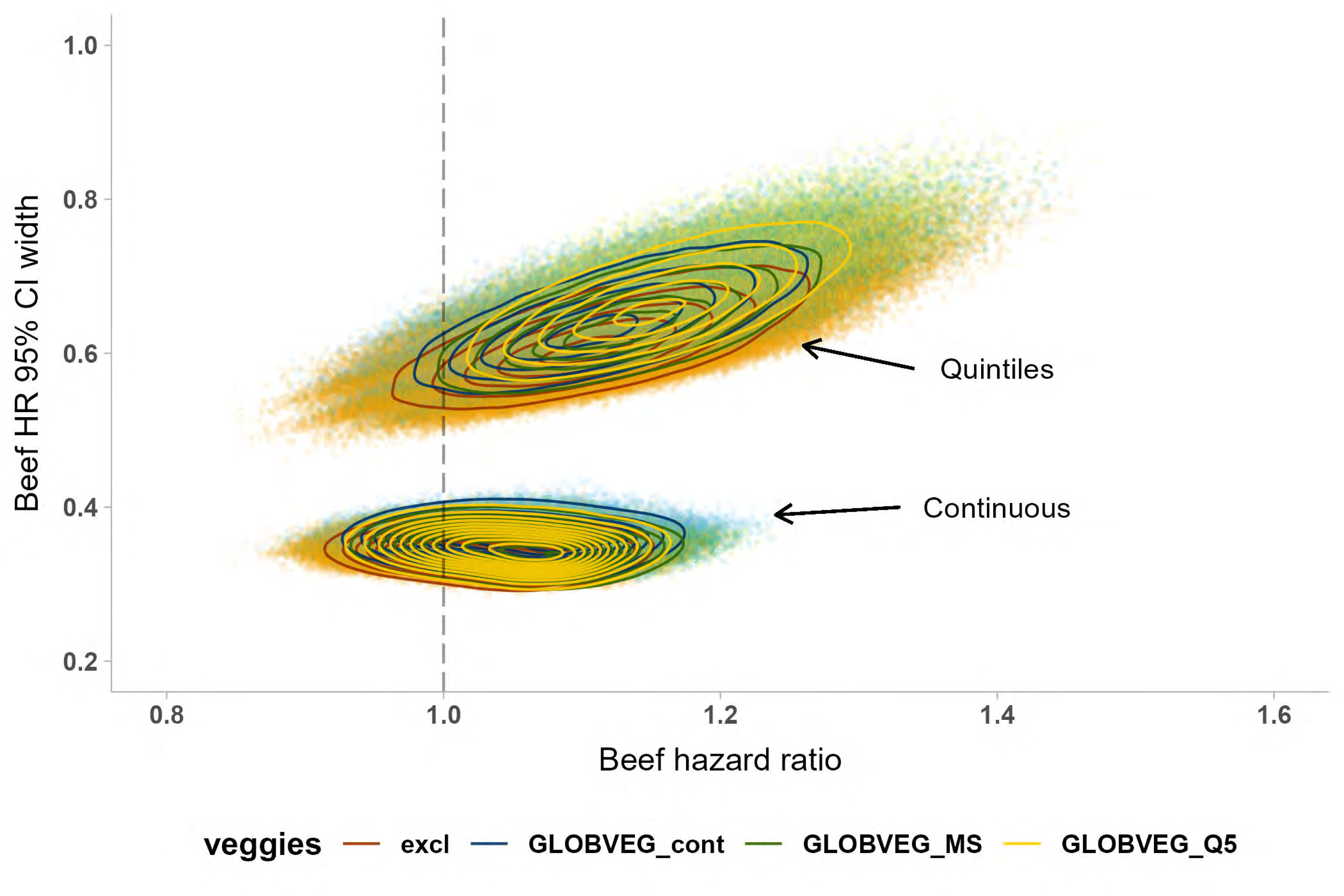

Beef HR 95% CI width
1.0
0.8
0.6
0.4
0.2
0.8
1.0
1.2
1.4
1.6
Beef hazard ratio
Quintiles
Continuous
veggies
excl
GLOBVEG_cont
GLOBVEG_MS
GLOBVEG_Q5

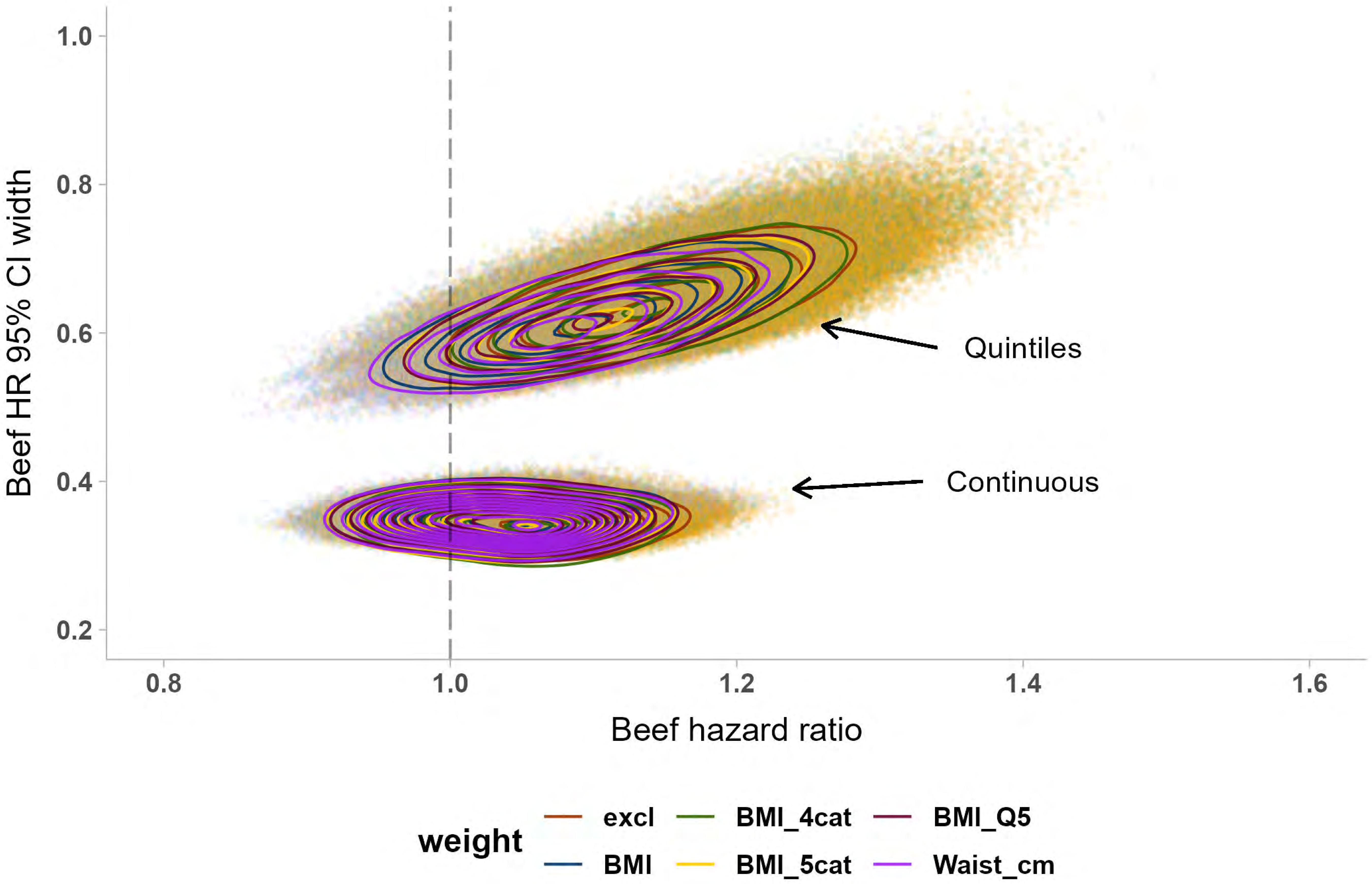

1.0
0.8
0.6
0.4
0.2
Beef HR 95% CI width
0.8
1.0
1.2
1.4
1.6
Beef hazard ratio
Quintiles
Continuous
weight
excl
BMI
BMI_4cat
BMI_5cat
BMI_Q5
Waist_cm

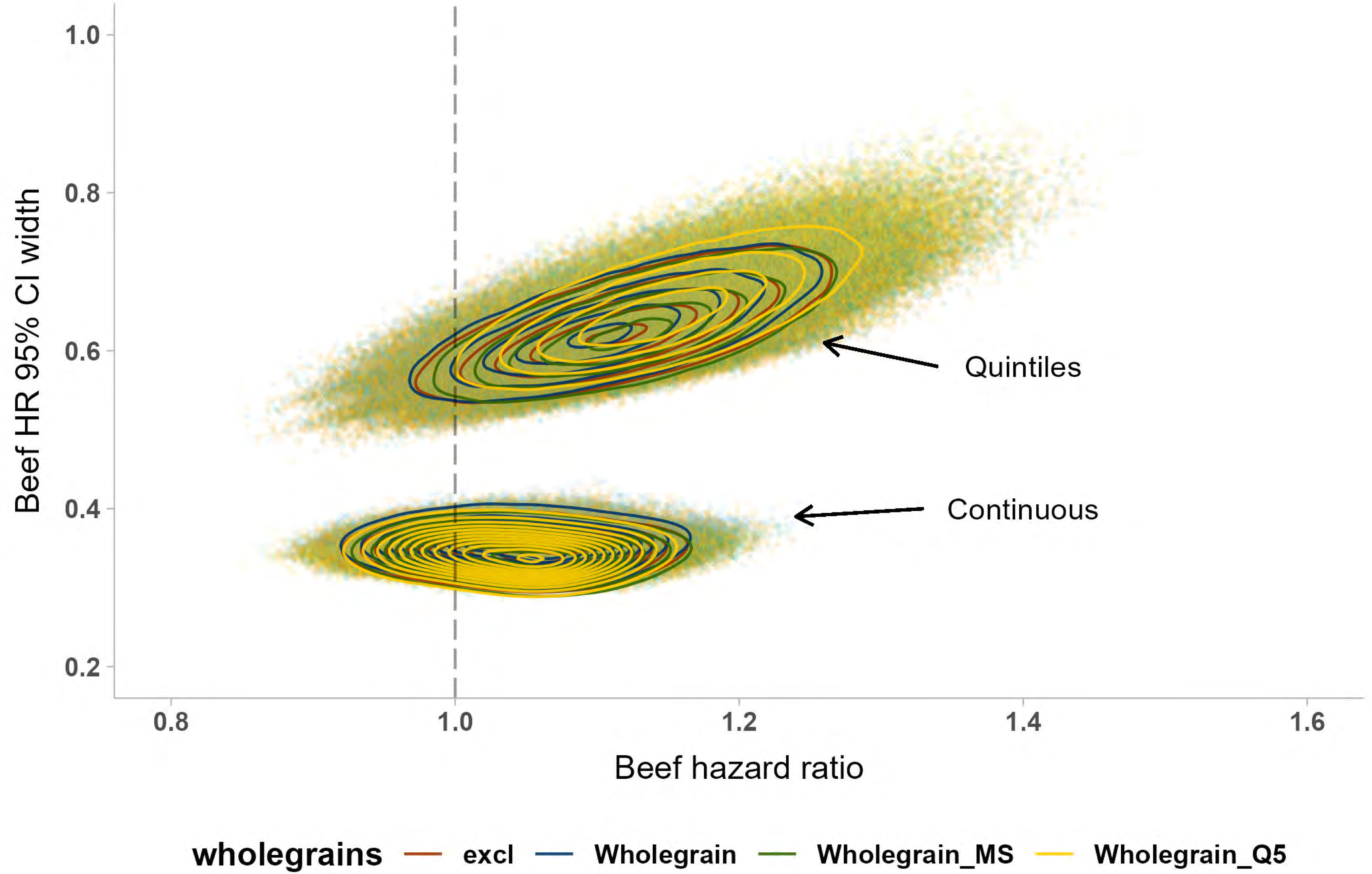

1.0
0.8
0.6
0.4
0.2
Beef HR 95% CI width
Quintiles
Continuous
0.8
1.0
1.2
1.4
1.6
Beef hazard ratio
wholegrains
excl
Wholegrain
Wholegrain_MS
Wholegrain_Q5
