## Supplemental File 2 for "‘Shaking the Ladder’ reveals how analytic choices can influence associations in nutrition epidemiology: beef intake and coronary heart disease as a case study"

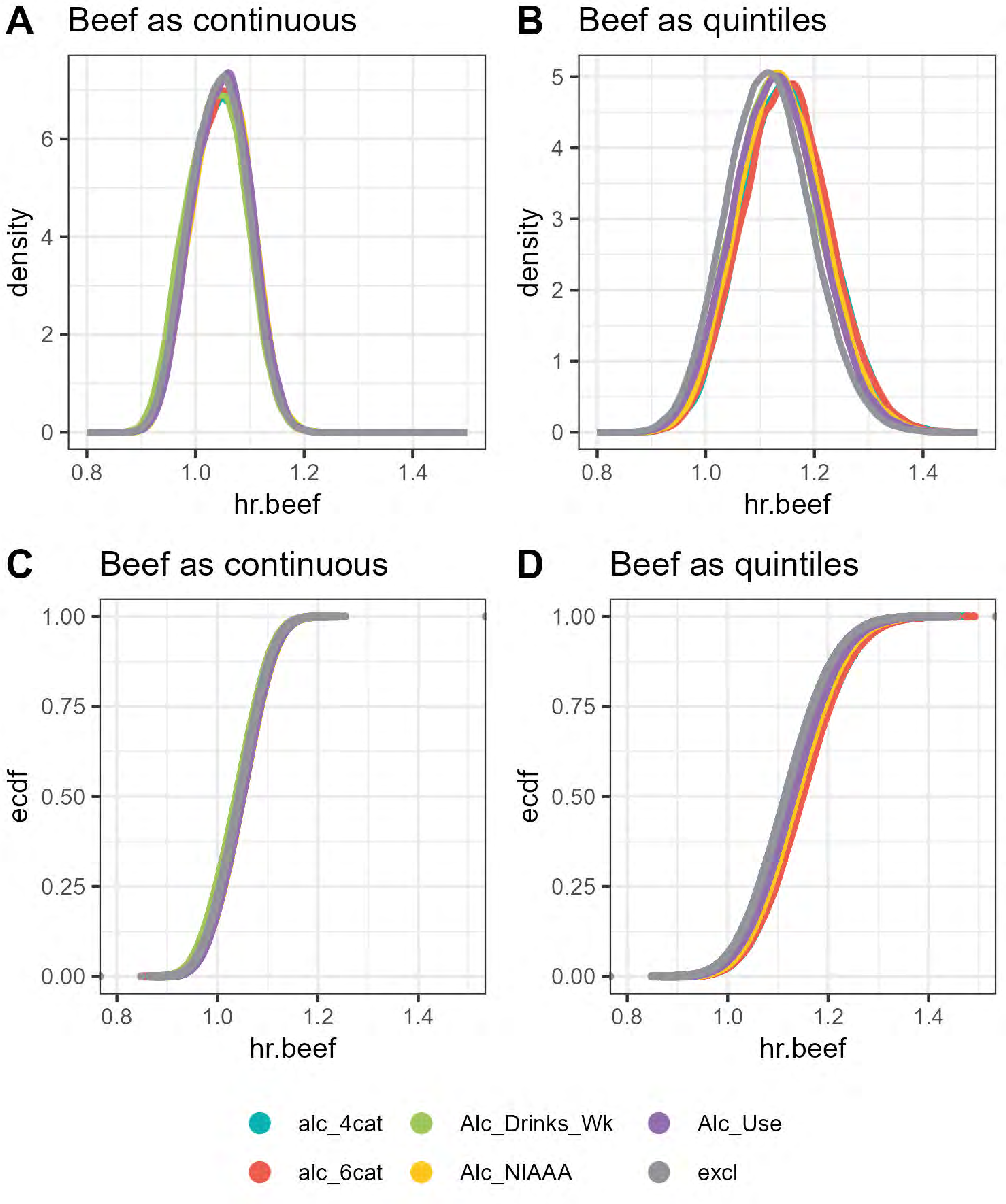

A
Beef as continuous
density
hr.beef
B
Beef as quintiles
density
hr.beef
C
Beef as continuous
ecdf
hr.beef
D
Beef as quintiles
ecdf
hr.beef
alc_4cat
Alc_Drinks_Wk
Alc_Use
alc_6cat
Alc_NIAAA
excl

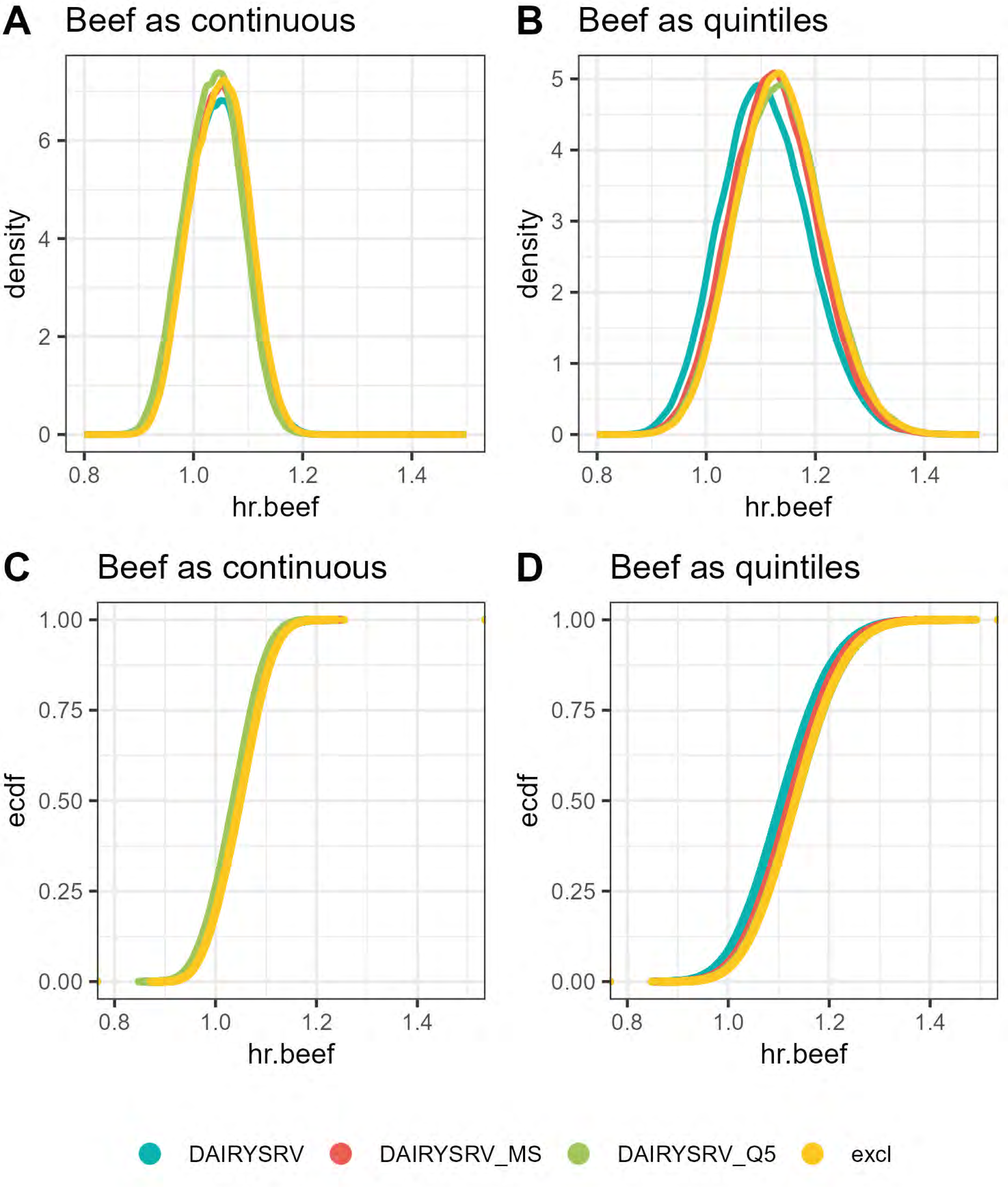

A Beef as continuous
density
hr.beef
B Beef as quintiles
density
hr.beef
C Beef as continuous
ecdf
hr.beef
D Beef as quintiles
ecdf
hr.beef
DAIRYSRV
DAIRYSRV_MS
DAIRYSRV_Q5
excl

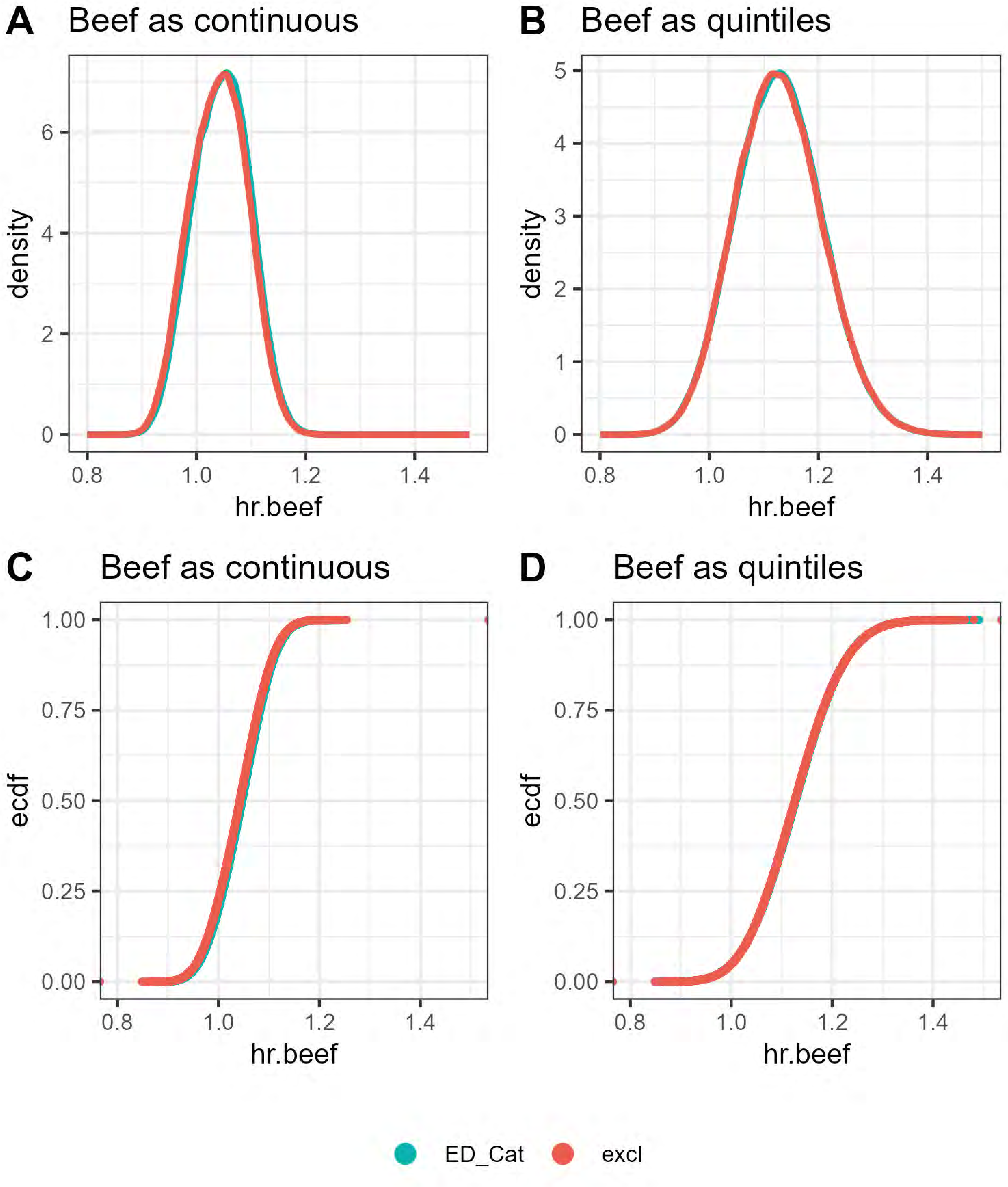

A
Beef as continuous
density
hr.beef
B
Beef as quintiles
density
hr.beef
C
Beef as continuous
ecdf
hr.beef
D
Beef as quintiles
ecdf
hr.beef
ED_Cat
excl

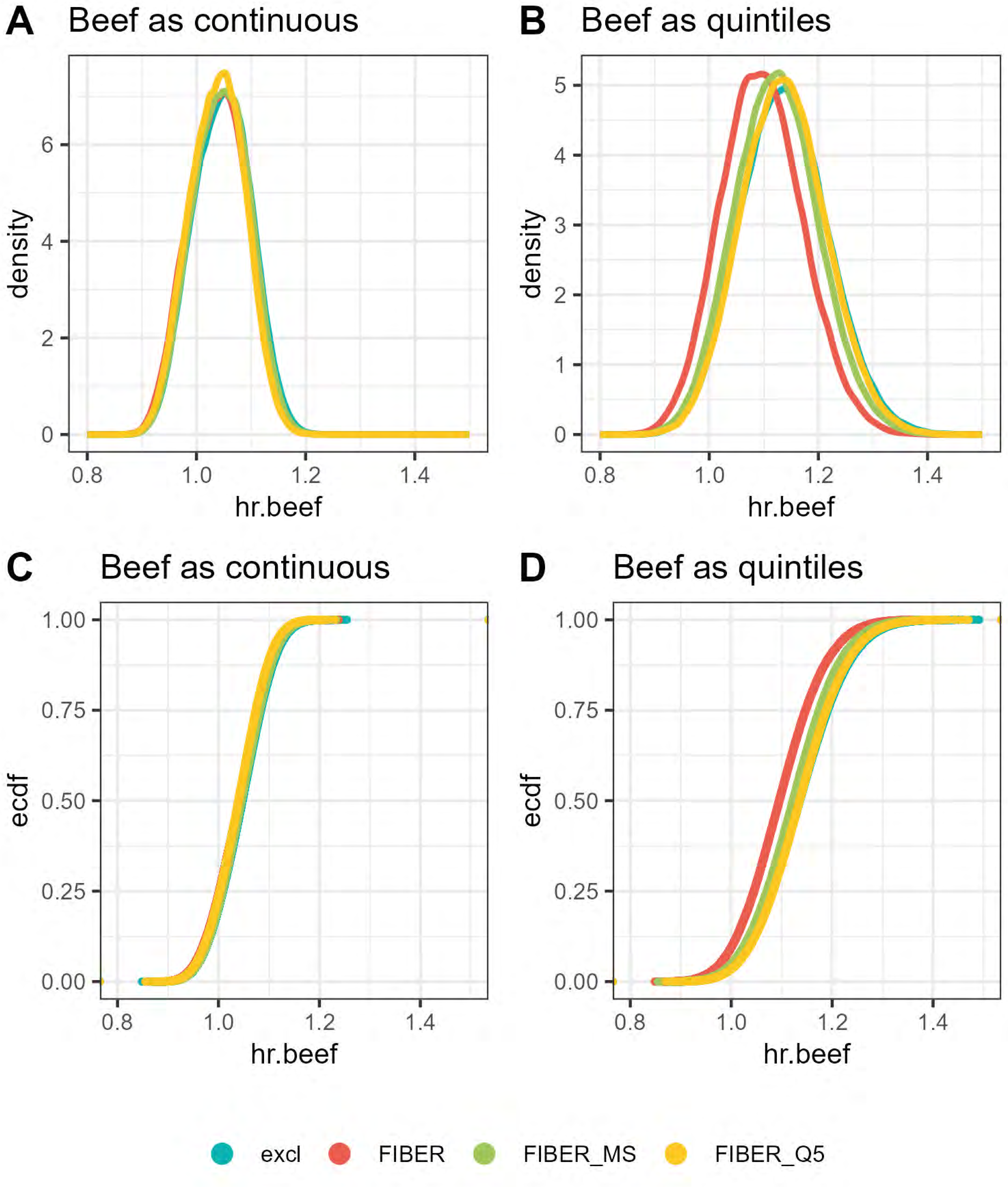

A
Beef as continuous
density
hr.beef
0
2
4
6
0.8
1.0
1.2
1.4
B
Beef as quintiles
density
hr.beef
0
1
2
3
4
5
0.8
1.0
1.2
1.4
C
Beef as continuous
ecdf
hr.beef
0.00
0.25
0.50
0.75
1.00
0.8
1.0
1.2
1.4
D
Beef as quintiles
ecdf
hr.beef
0.00
0.25
0.50
0.75
1.00
0.8
1.0
1.2
1.4
excl
FIBER
FIBER_MS
FIBER_Q5

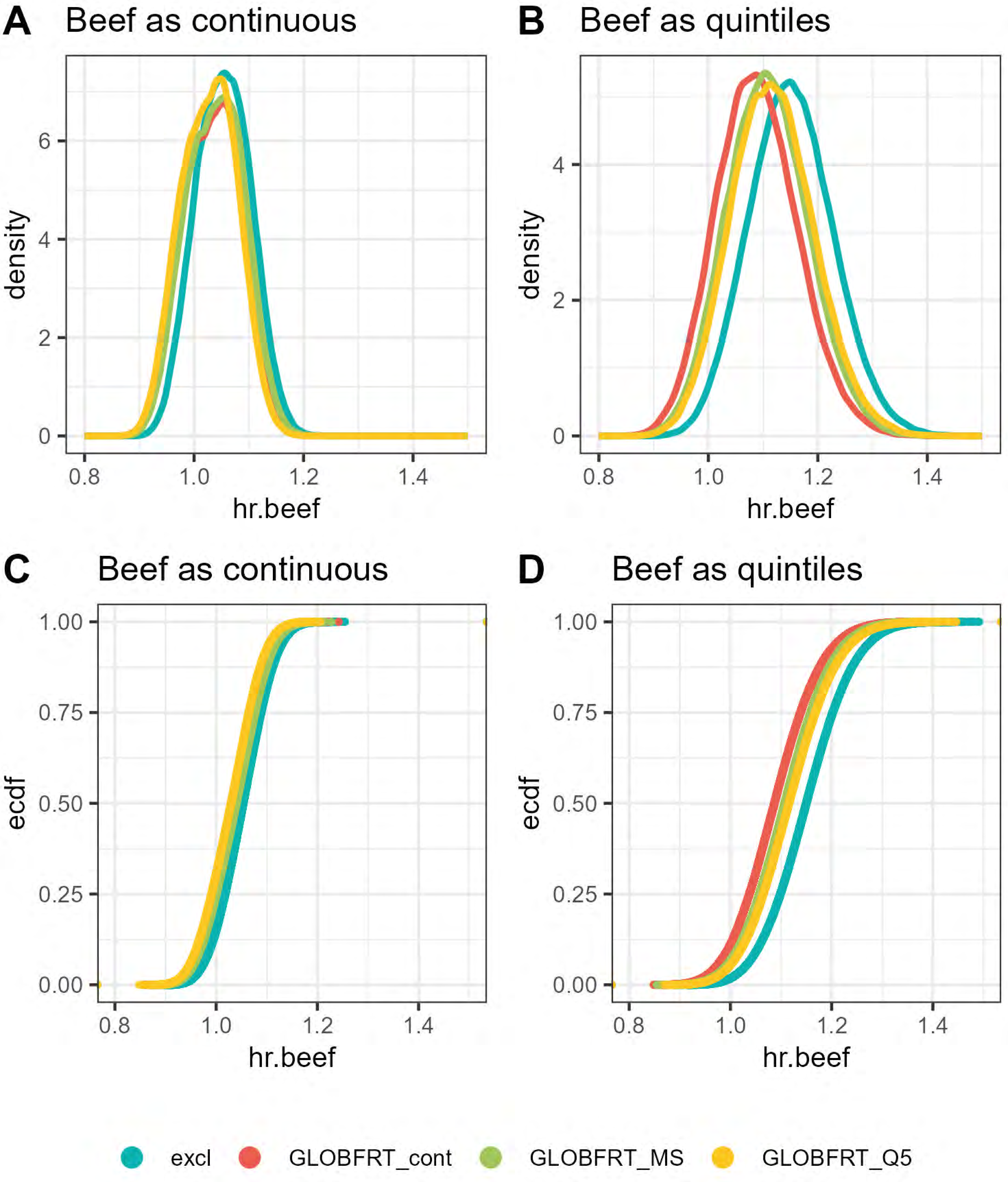
A
Beef as continuous
B
Beef as quintiles
C
Beef as continuous
D
Beef as quintiles
density
ecdf
hr.beef
0
2
4
6
0.00
0.25
0.50
0.75
1.00
0.8
1.0
1.2
1.4
excl
GLOBFRT_cont
GLOBFRT_MS
GLOBFRT_Q5

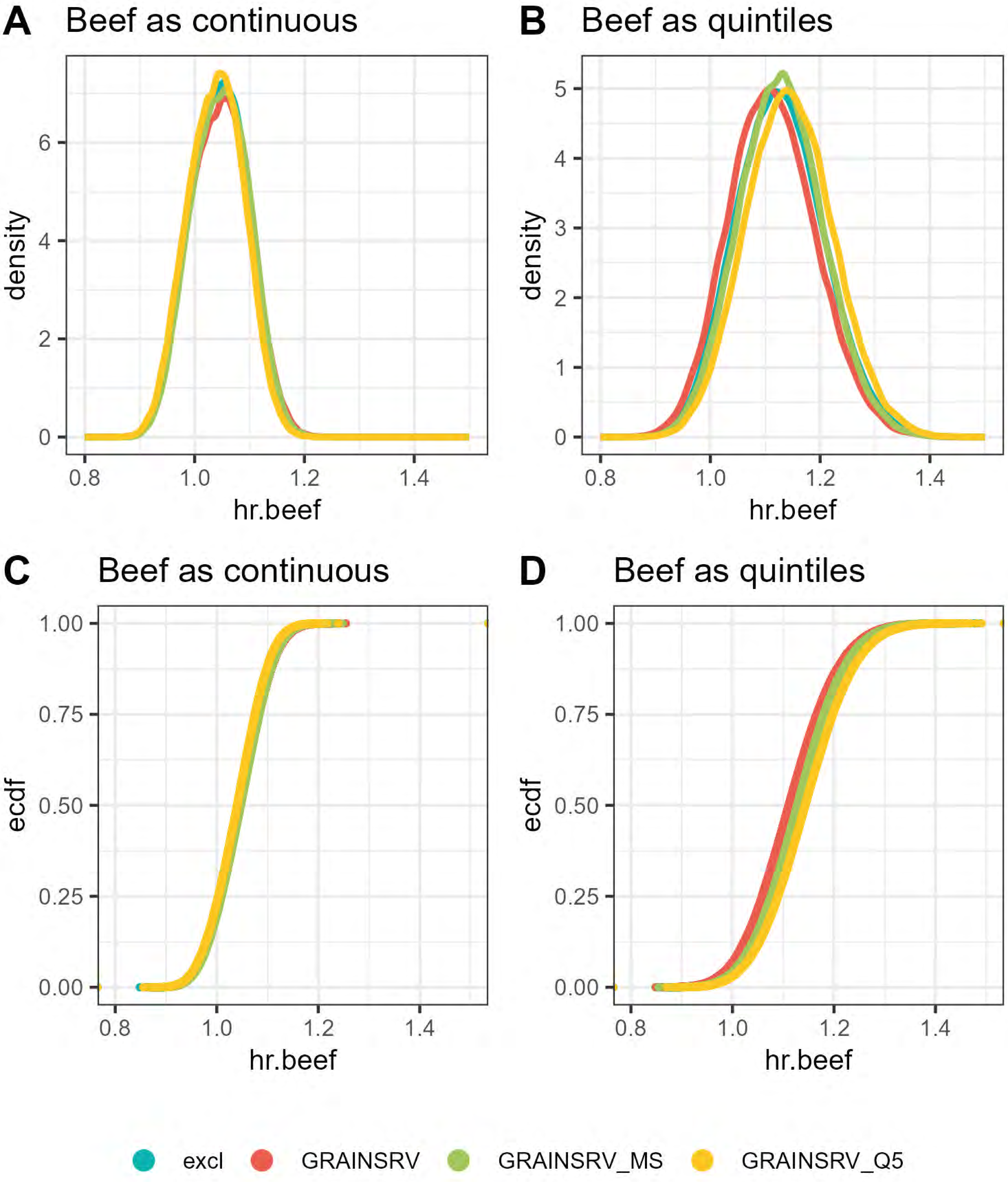
A
Beef as continuous
B
Beef as quintiles
C
Beef as continuous
D
Beef as quintiles
density
ecdf
hr.beef
0
2
4
6
1
3
5
0.00
0.25
0.50
0.75
1.00
0.8
1.0
1.2
1.4
excl
GRAINSRV
GRAINSRV_MS
GRAINSRV_Q5

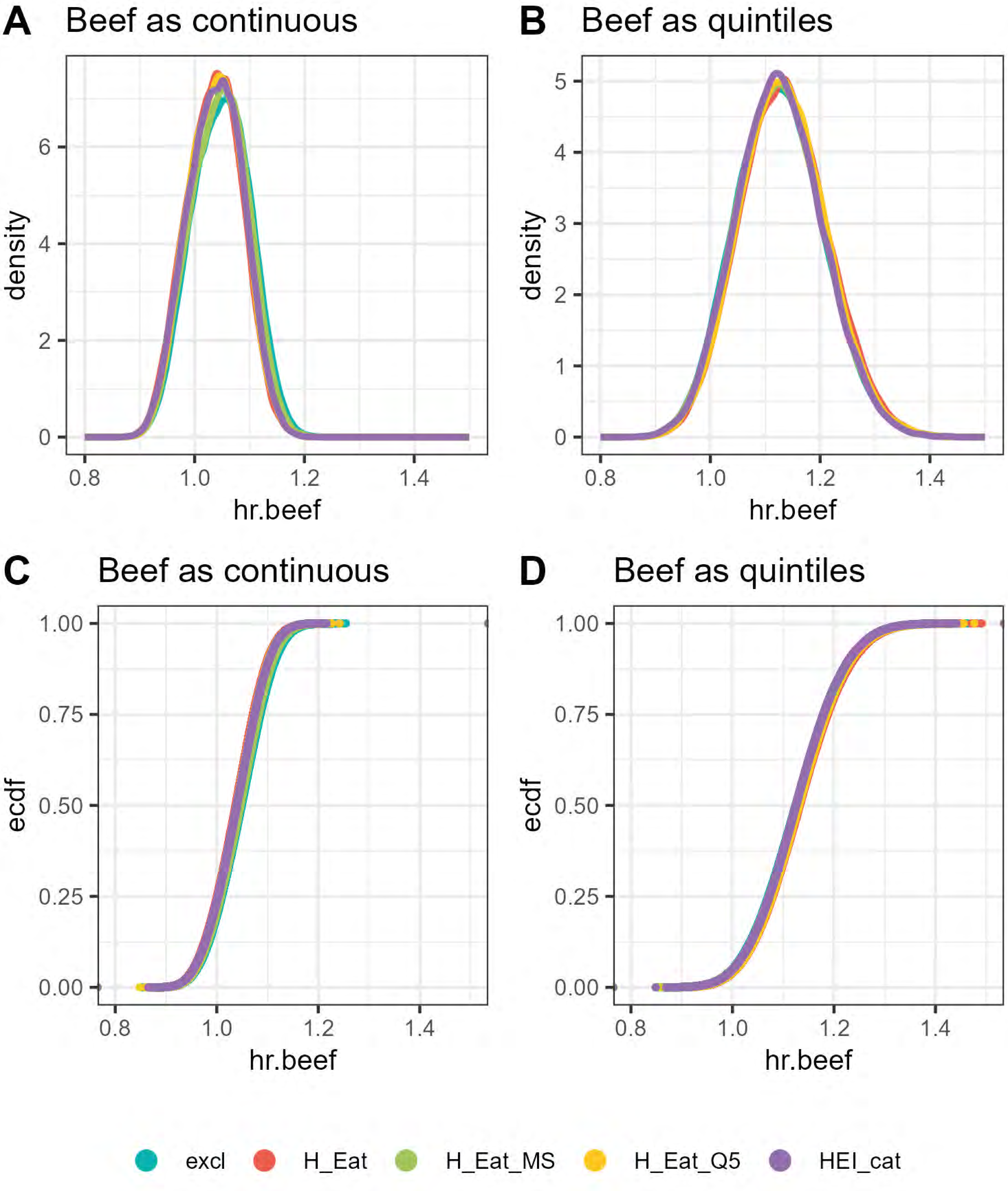

A
Beef as continuous
density
hr.beef
B
Beef as quintiles
density
hr.beef
C
Beef as continuous
ecdf
hr.beef
D
Beef as quintiles
ecdf
hr.beef
excl
H_Eat
H_Eat_MS
H_Eat_Q5
HEI_cat

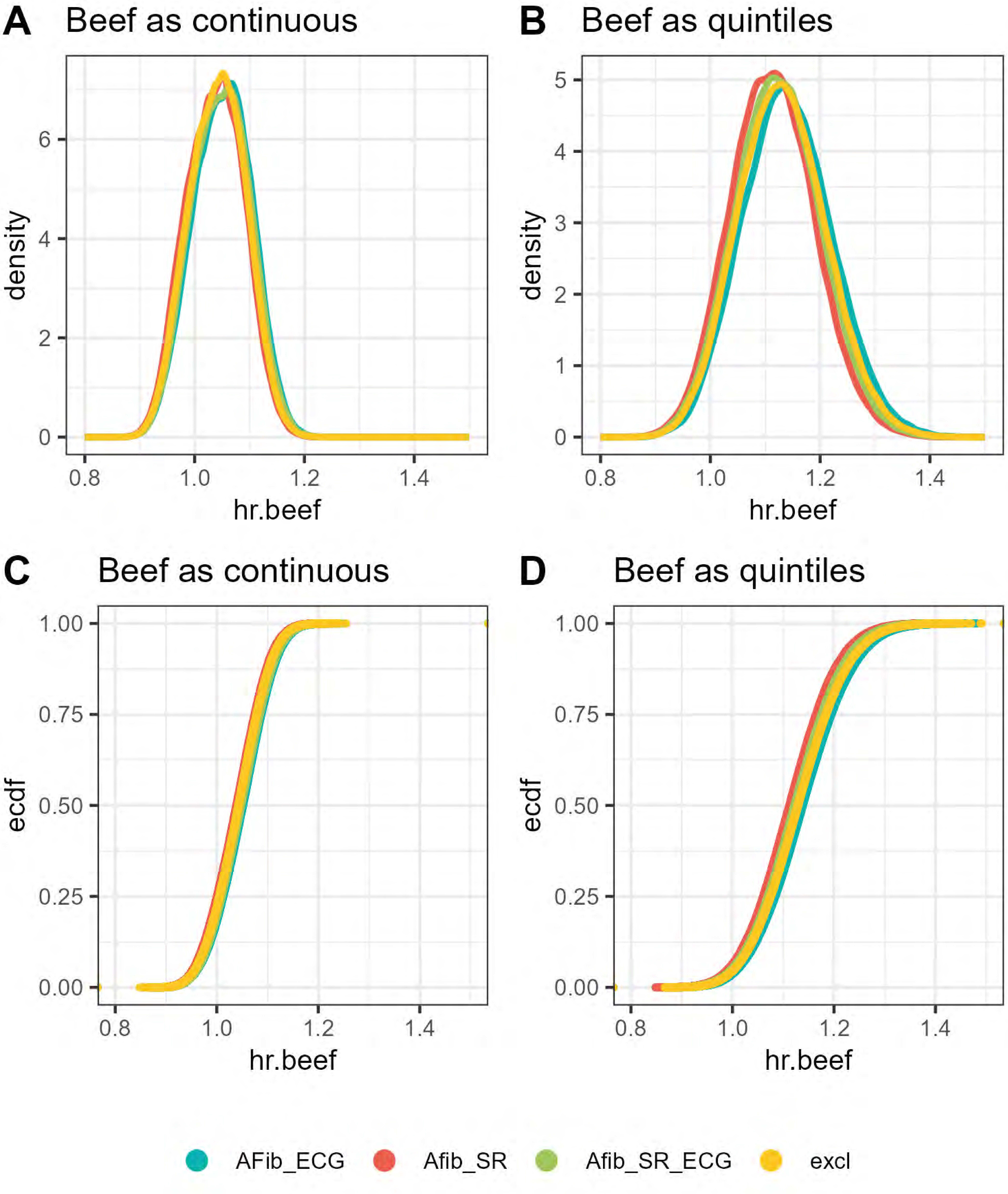
A Beef as continuous
density
hr.beef
0
2
4
6
0.8
1.0
1.2
1.4
B Beef as quintiles
density
hr.beef
0
1
2
3
4
5
C Beef as continuous
ecdf
0.00
0.25
0.50
0.75
1.00
D Beef as quintiles
AFib_ECG
Afib_SR
Afib_SR_ECG
excl

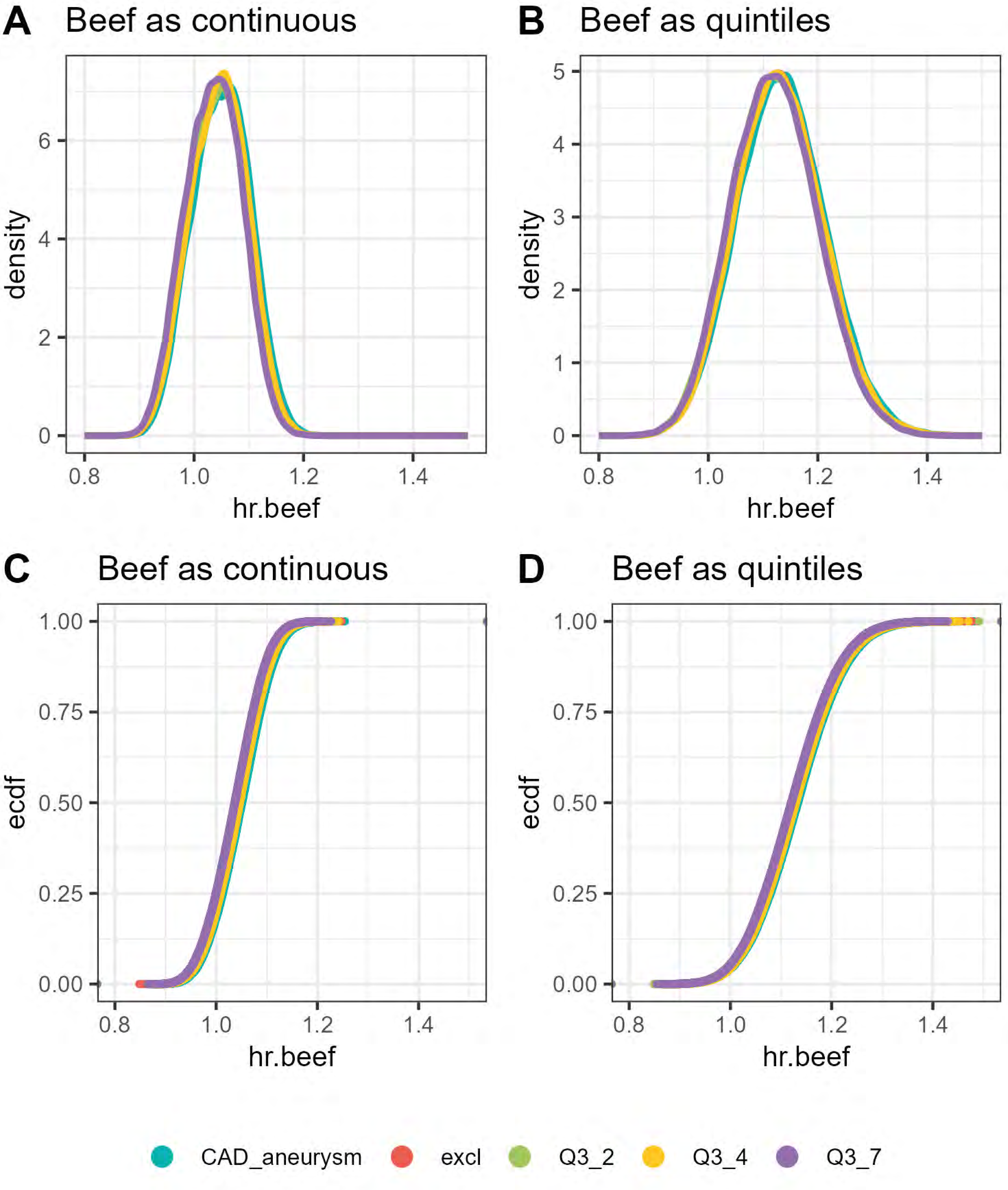
A
Beef as continuous
density
hr.beef
B
Beef as quintiles
density
hr.beef
C
Beef as continuous
ecdf
hr.beef
D
Beef as quintiles
ecdf
hr.beef
CAD_aneurysm
excl
Q3_2
Q3_4
Q3_7

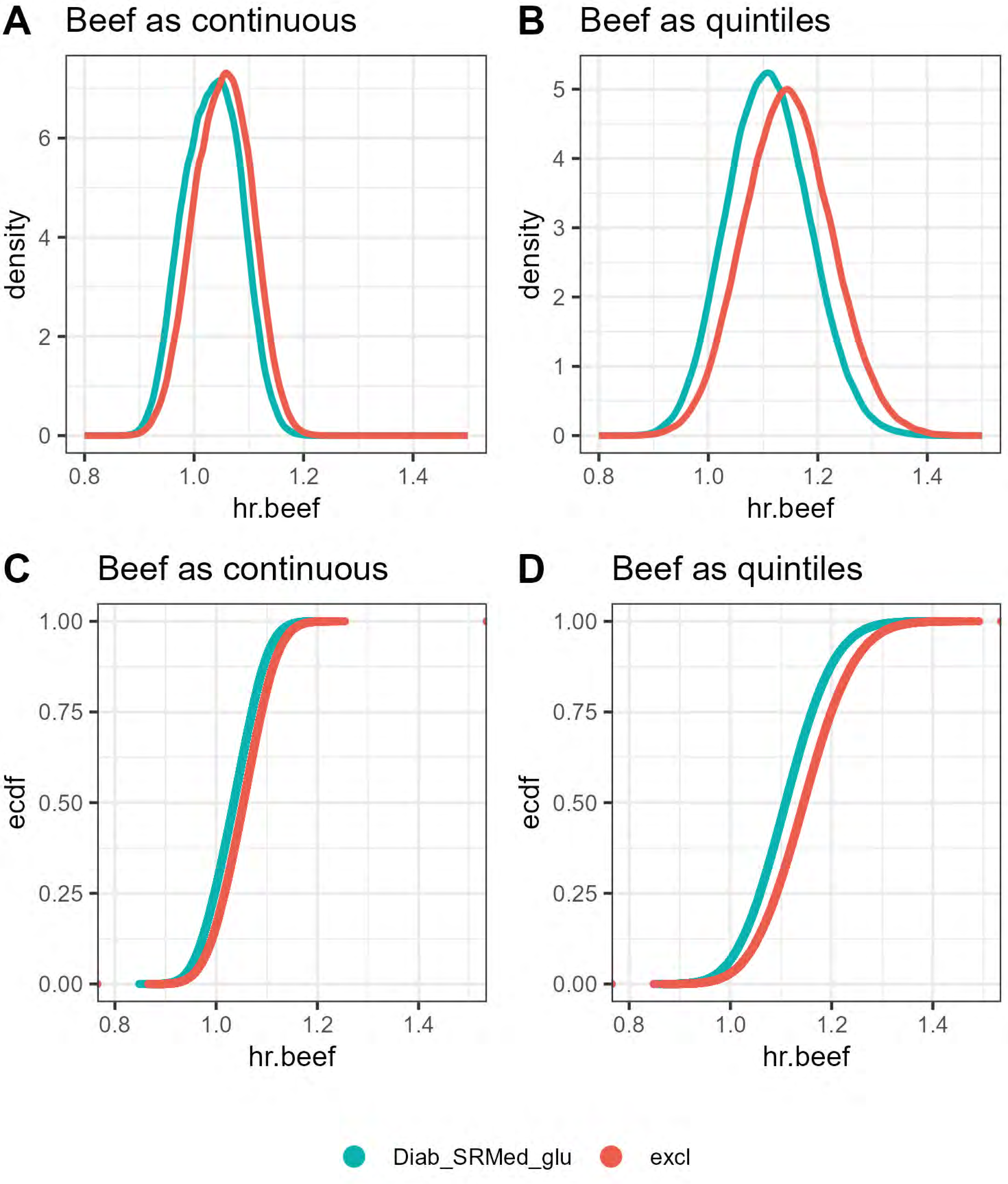
A
Beef as continuous
density
hr.beef
B
Beef as quintiles
density
hr.beef
C
Beef as continuous
ecdf
hr.beef
D
Beef as quintiles
ecdf
hr.beef
Diab_SRMed_glu
excl

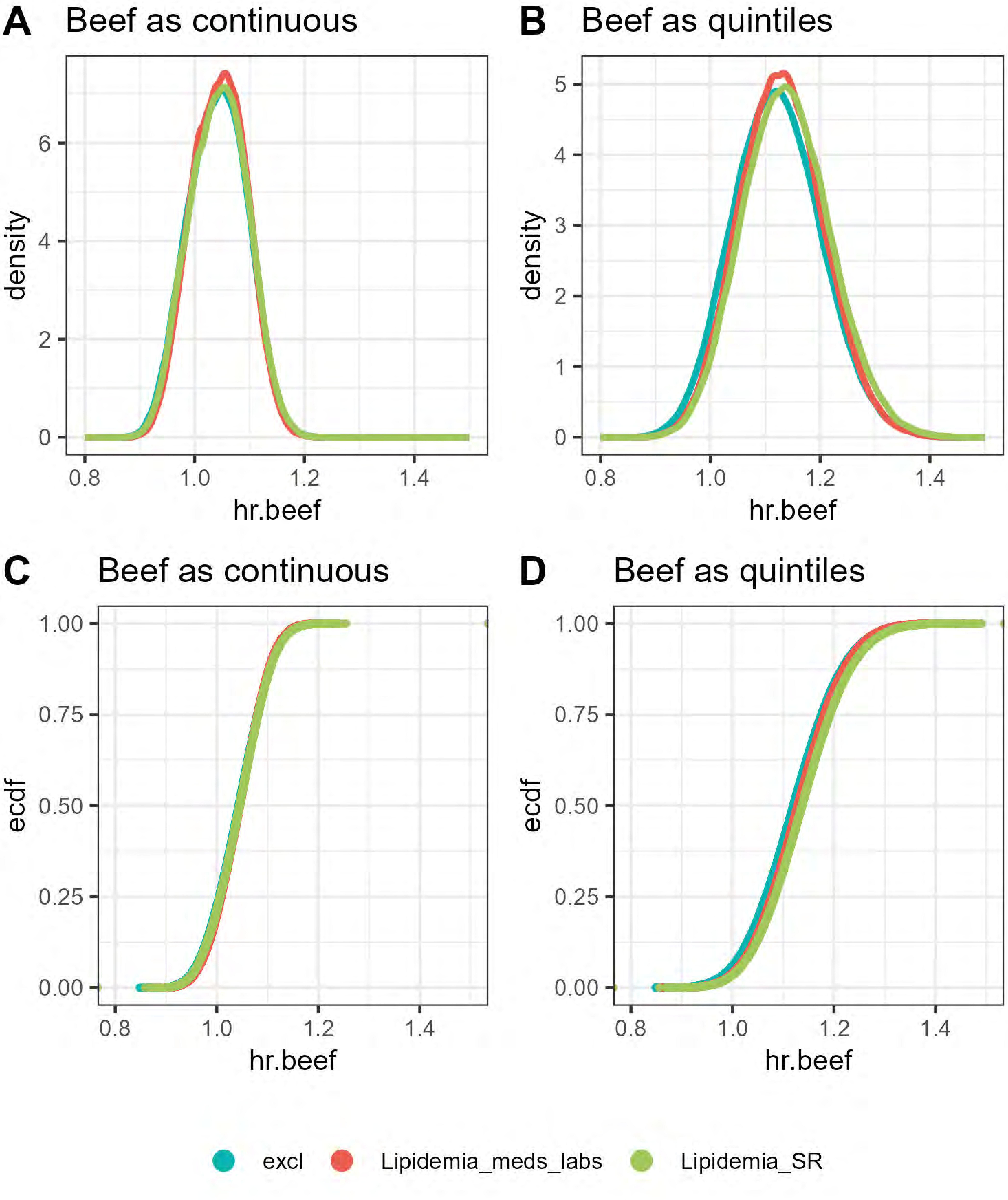
A
Beef as continuous
B
Beef as quintiles
C
Beef as continuous
D
Beef as quintiles
density
ecdf
hr.beef
0
2
4
6
1
3
5
0.00
0.25
0.50
0.75
1.00
0.8
1.0
1.2
1.4
excl
Lipidemia_meds_labs
Lipidemia_SR

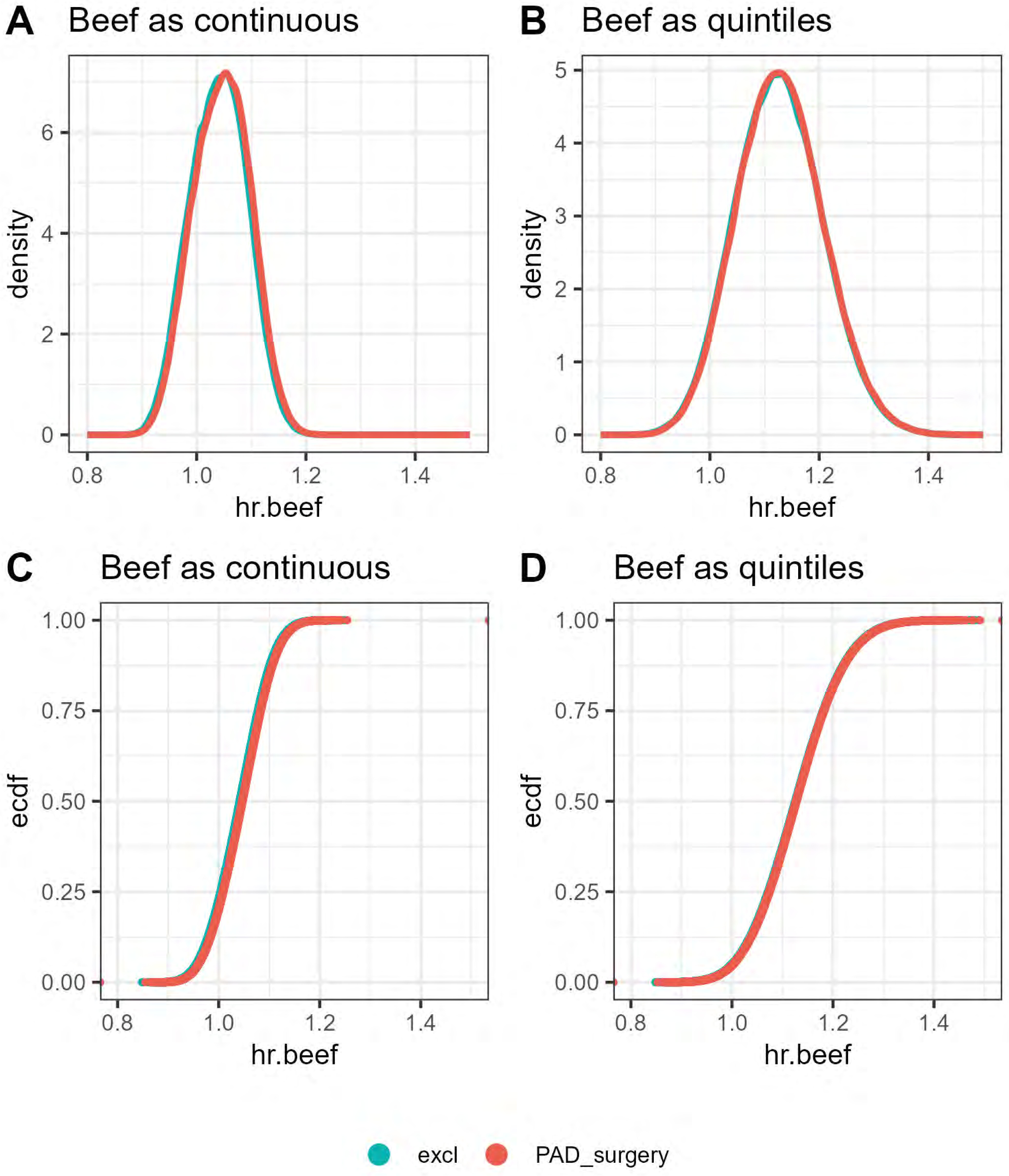

A Beef as continuous
B Beef as quintiles
density
hr.beef
C Beef as continuous
D Beef as quintiles
ecdf
hr.beef
excl
PAD_surgery

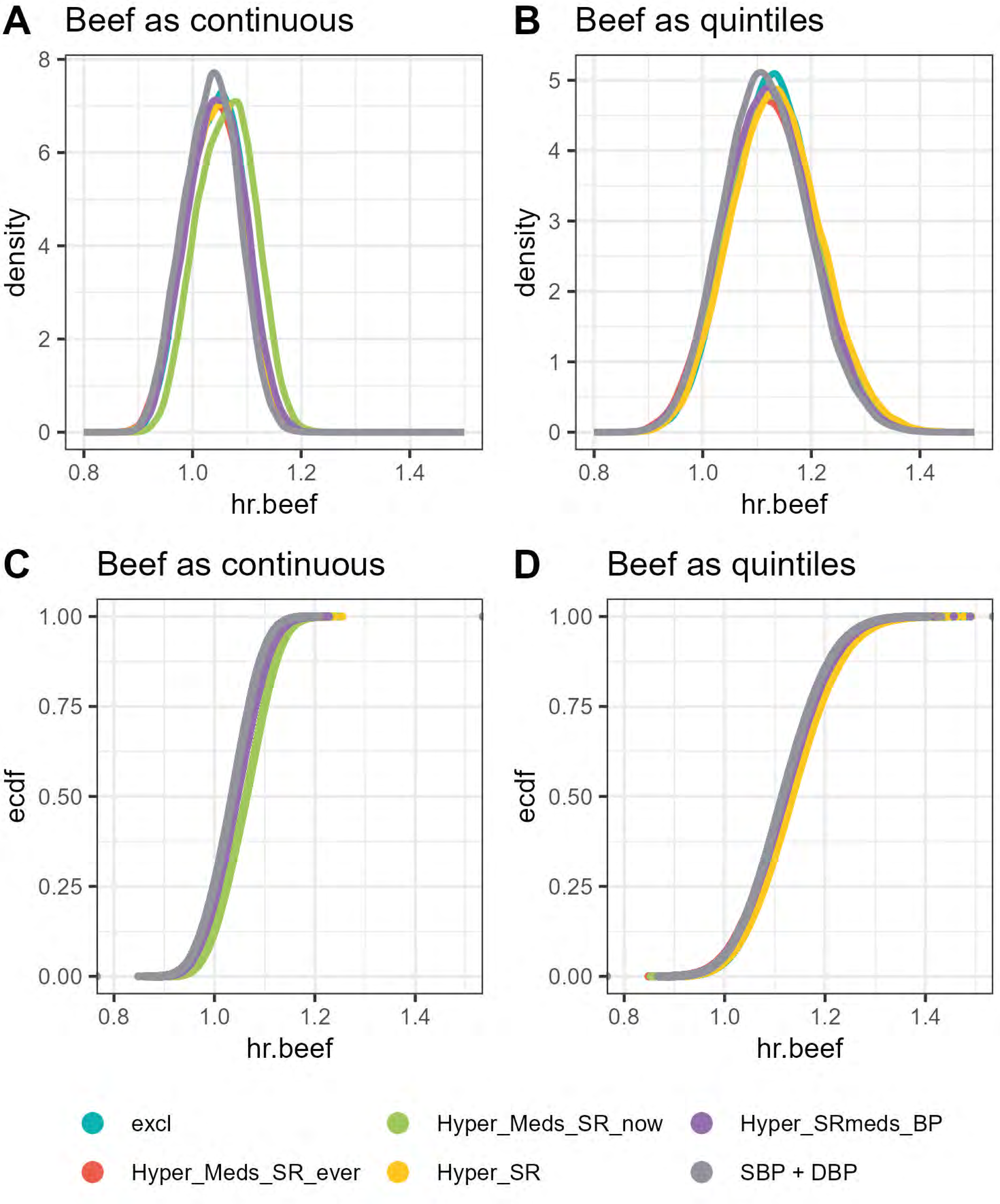
A
Beef as continuous
B
Beef as quintiles
C
Beef as continuous
D
Beef as quintiles
density
ecdf
hr.beef
0
2
4
6
8
1
3
5
0.00
0.25
0.50
0.75
1.00
0.8
1.0
1.2
1.4
excl
Hyper_Meds_SR_ever
Hyper_Meds_SR_now
Hyper_SR
Hyper_SRmeds_BP
SBP + DBP

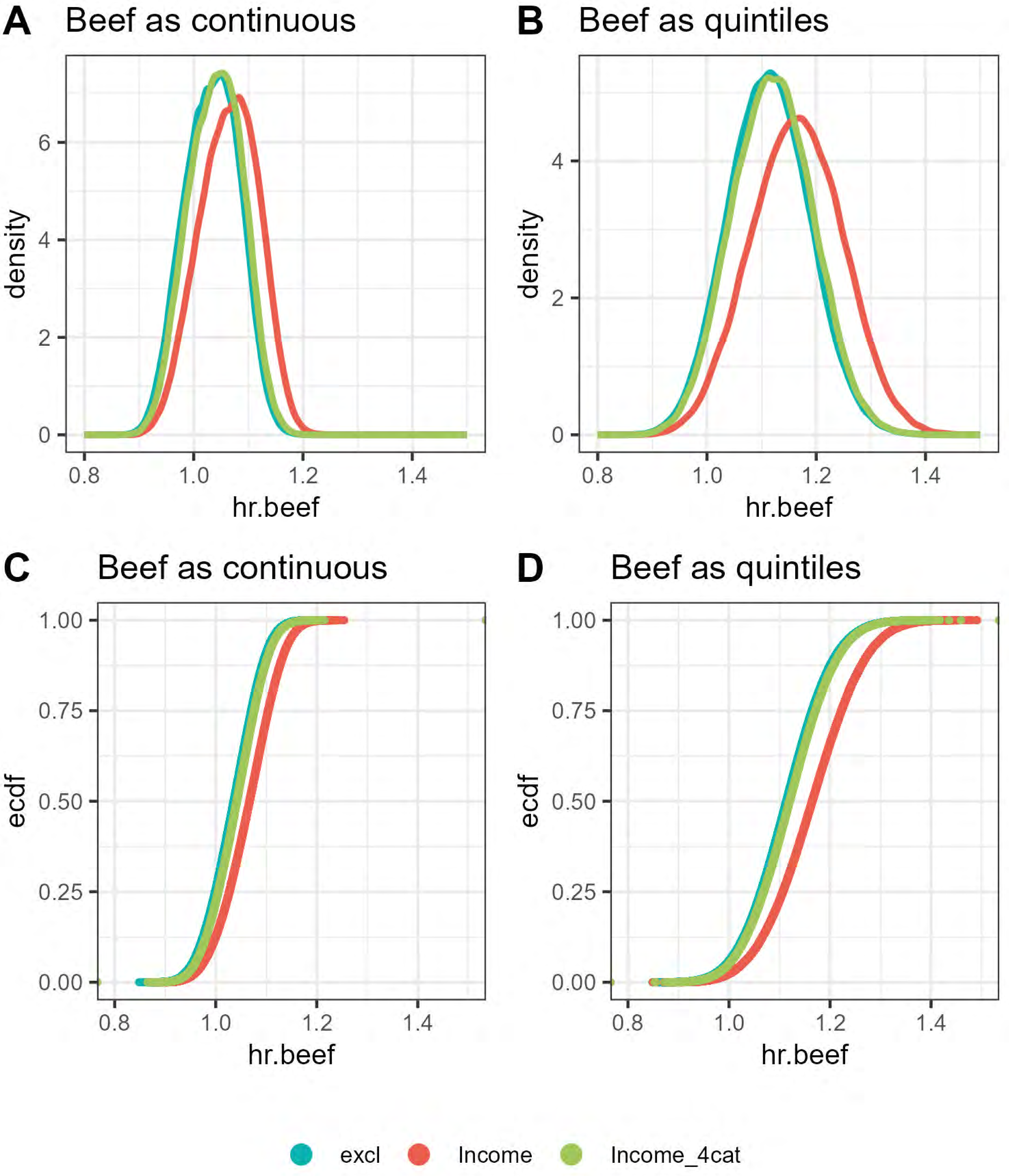
A
Beef as continuous
density
hr.beef
B
Beef as quintiles
density
hr.beef
C
Beef as continuous
ecdf
hr.beef
D
Beef as quintiles
ecdf
hr.beef
excl
Income
Income_4cat

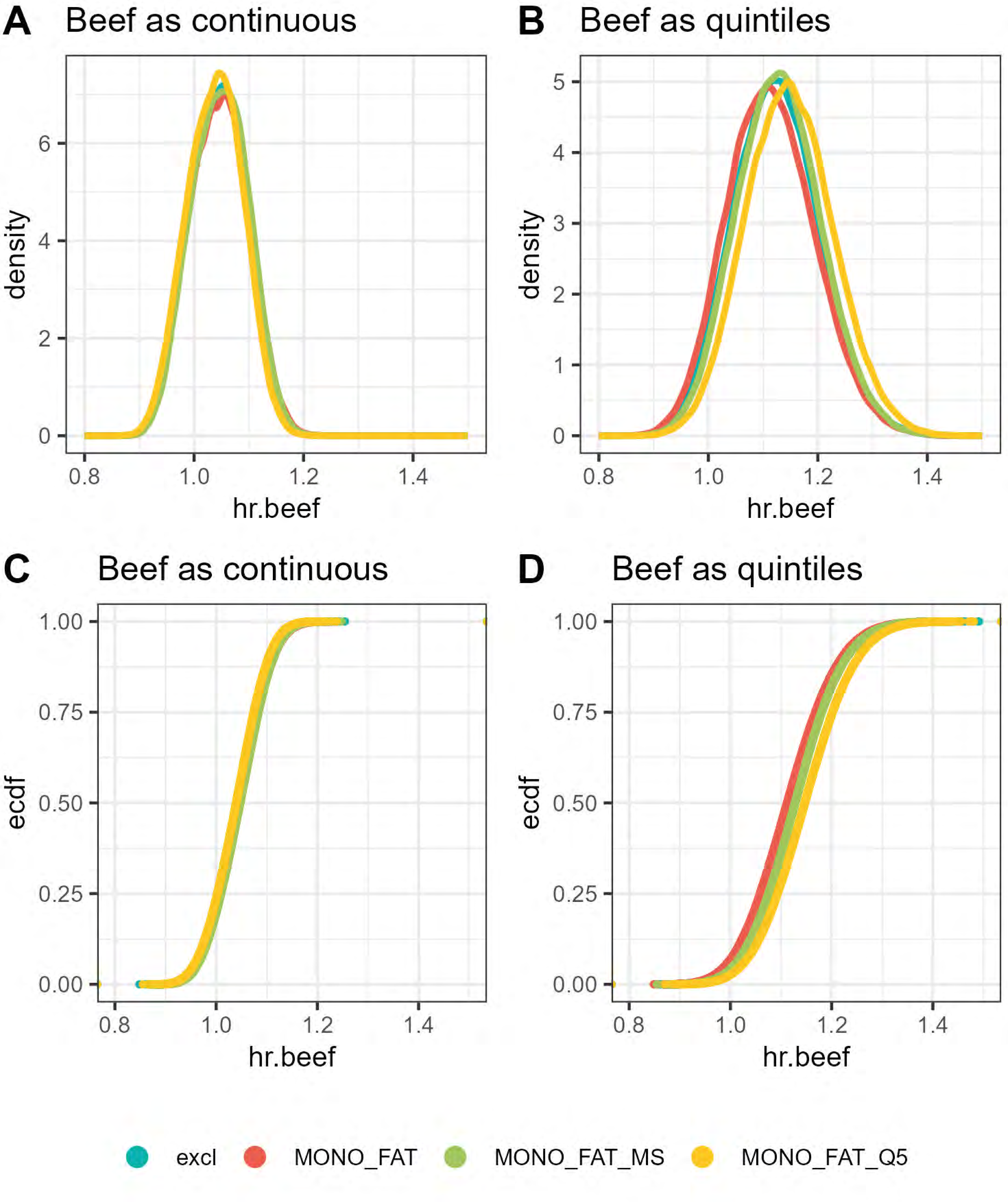

A
Beef as continuous
density
hr.beef
B
Beef as quintiles
density
hr.beef
C
Beef as continuous
ecdf
hr.beef
D
Beef as quintiles
ecdf
hr.beef
excl
MONO_FAT
MONO_FAT_MS
MONO_FAT_Q5

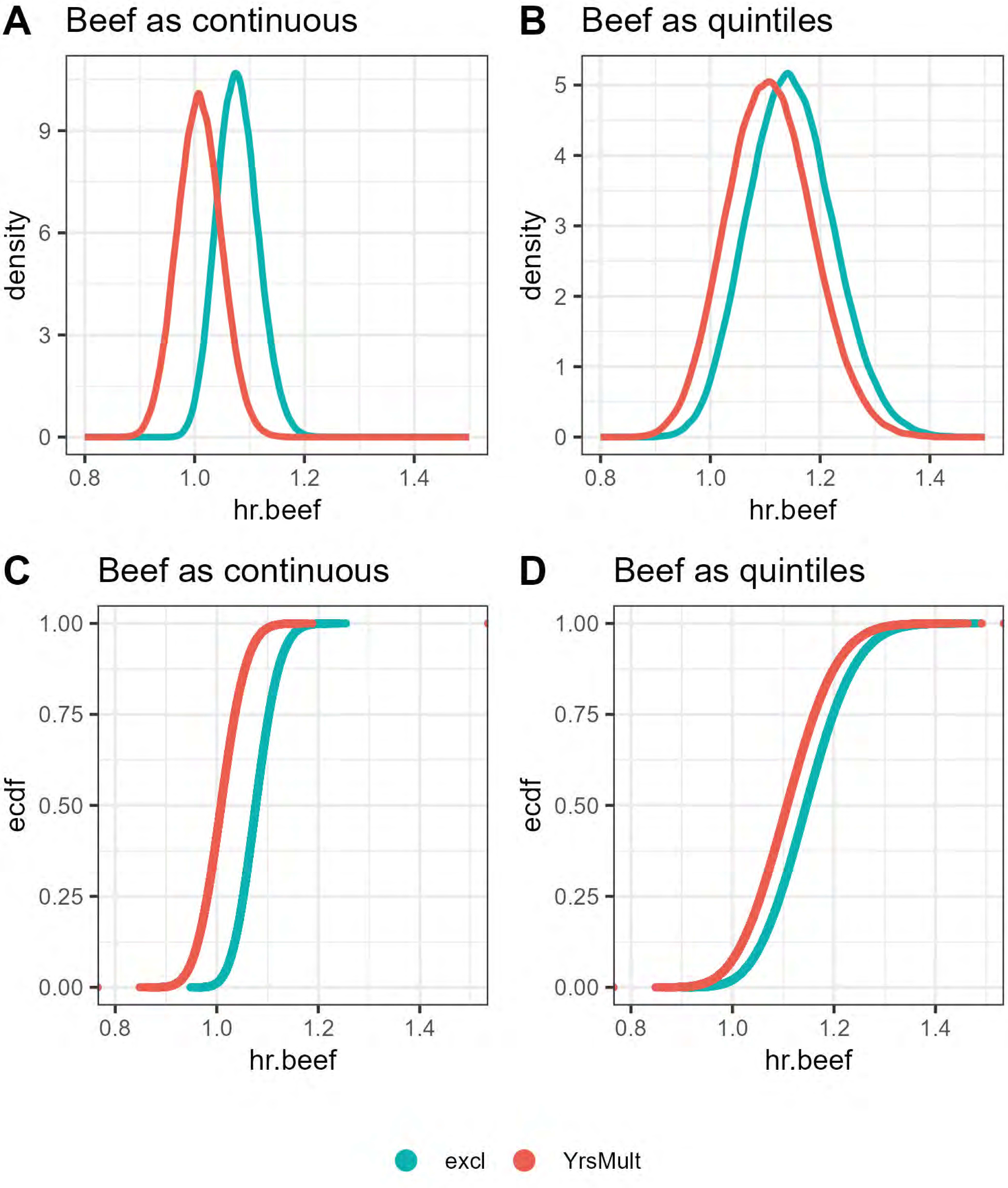
A
Beef as continuous
density
hr.beef
B
Beef as quintiles
density
hr.beef
C
Beef as continuous
ecdf
hr.beef
D
Beef as quintiles
ecdf
hr.beef
excl
YrsMult

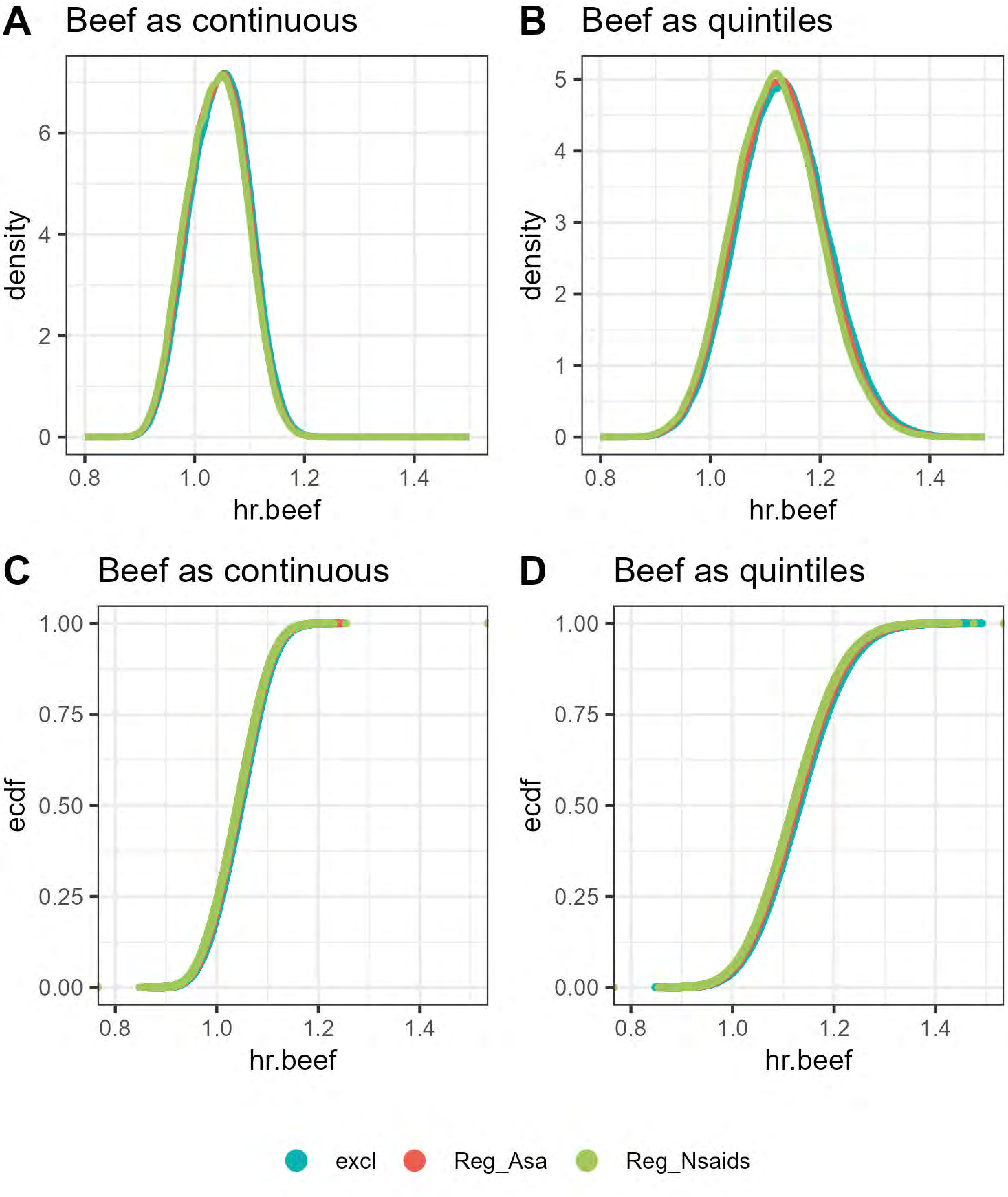
A
Beef as continuous
density
hr.beef
B
Beef as quintiles
density
hr.beef
C
Beef as continuous
ecdf
hr.beef
D
Beef as quintiles
ecdf
hr.beef
excl
Reg_Asa
Reg_Nsaids

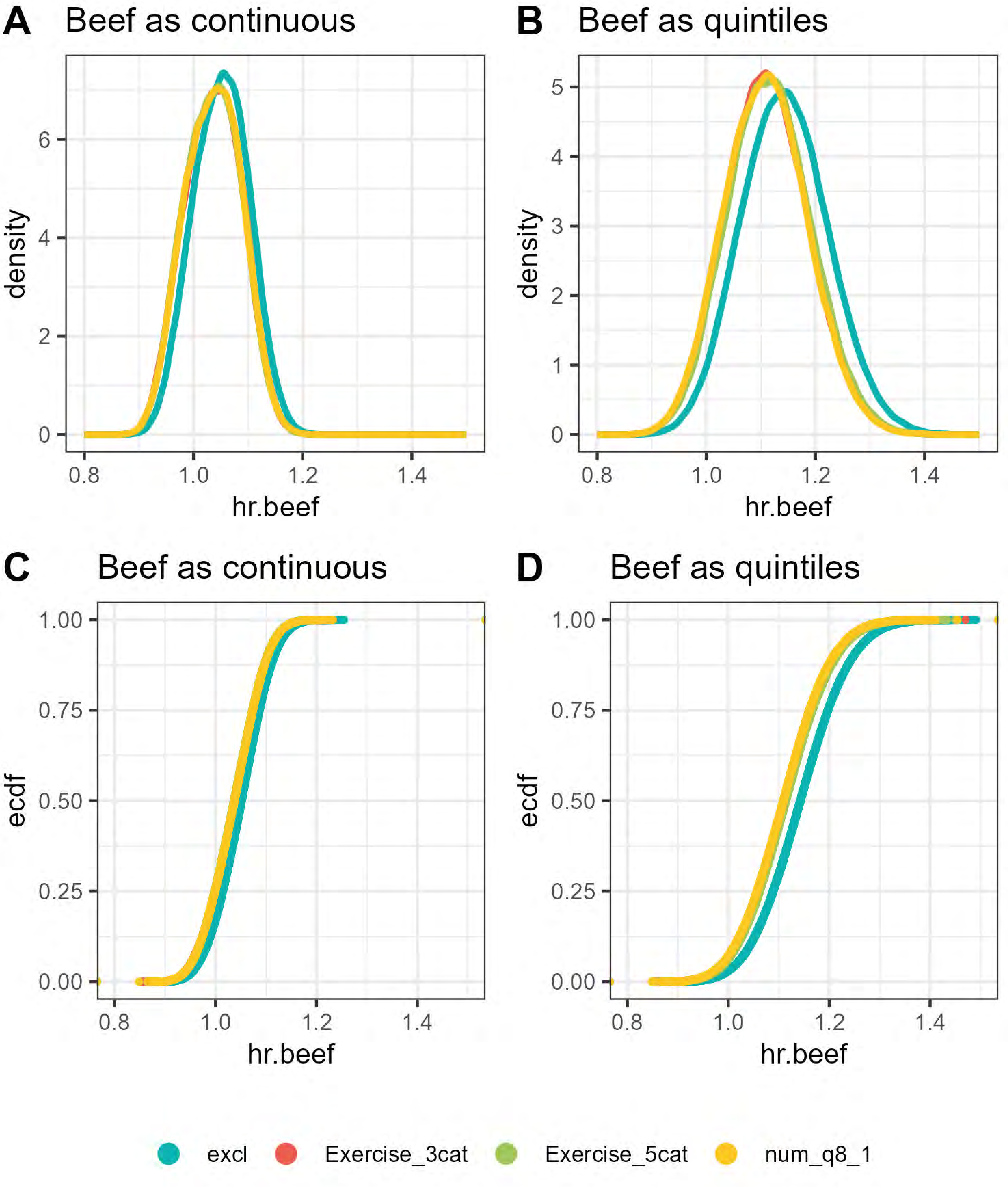
A
Beef as continuous
density
hr.beef
B
Beef as quintiles
density
hr.beef
C
Beef as continuous
ecdf
hr.beef
D
Beef as quintiles
ecdf
hr.beef
excl
Exercise_3cat
Exercise_5cat
num_q8_1

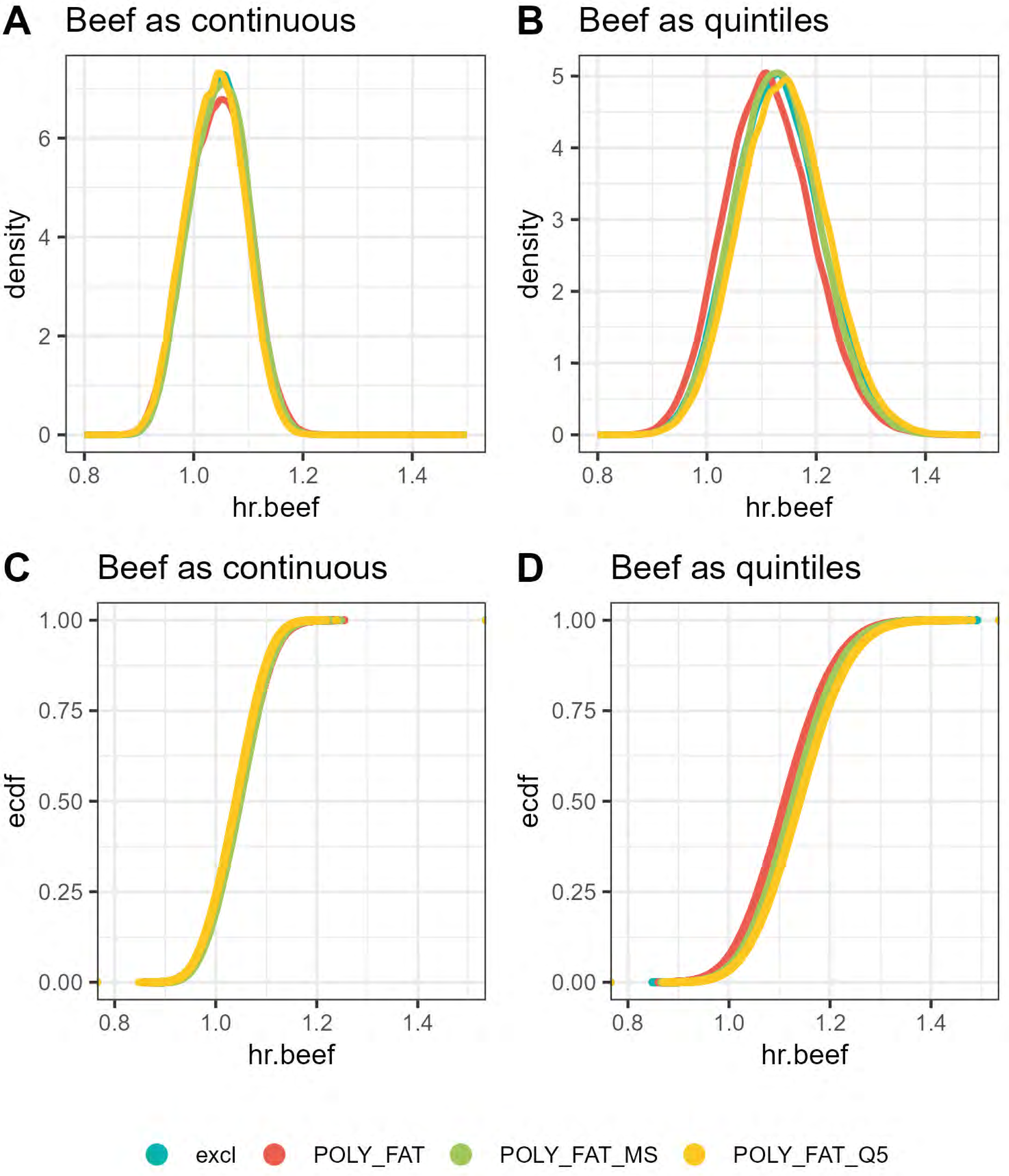
A
Beef as continuous
density
hr.beef
0
2
4
6
0.8
1.0
1.2
1.4
B
Beef as quintiles
density
hr.beef
0
1
2
3
4
5
0.8
1.0
1.2
1.4
C
Beef as continuous
ecdf
hr.beef
0.00
0.25
0.50
0.75
1.00
0.8
1.0
1.2
1.4
D
Beef as quintiles
ecdf
hr.beef
0.00
0.25
0.50
0.75
1.00
0.8
1.0
1.2
1.4
excl
POLY_FAT
POLY_FAT_MS
POLY_FAT_Q5

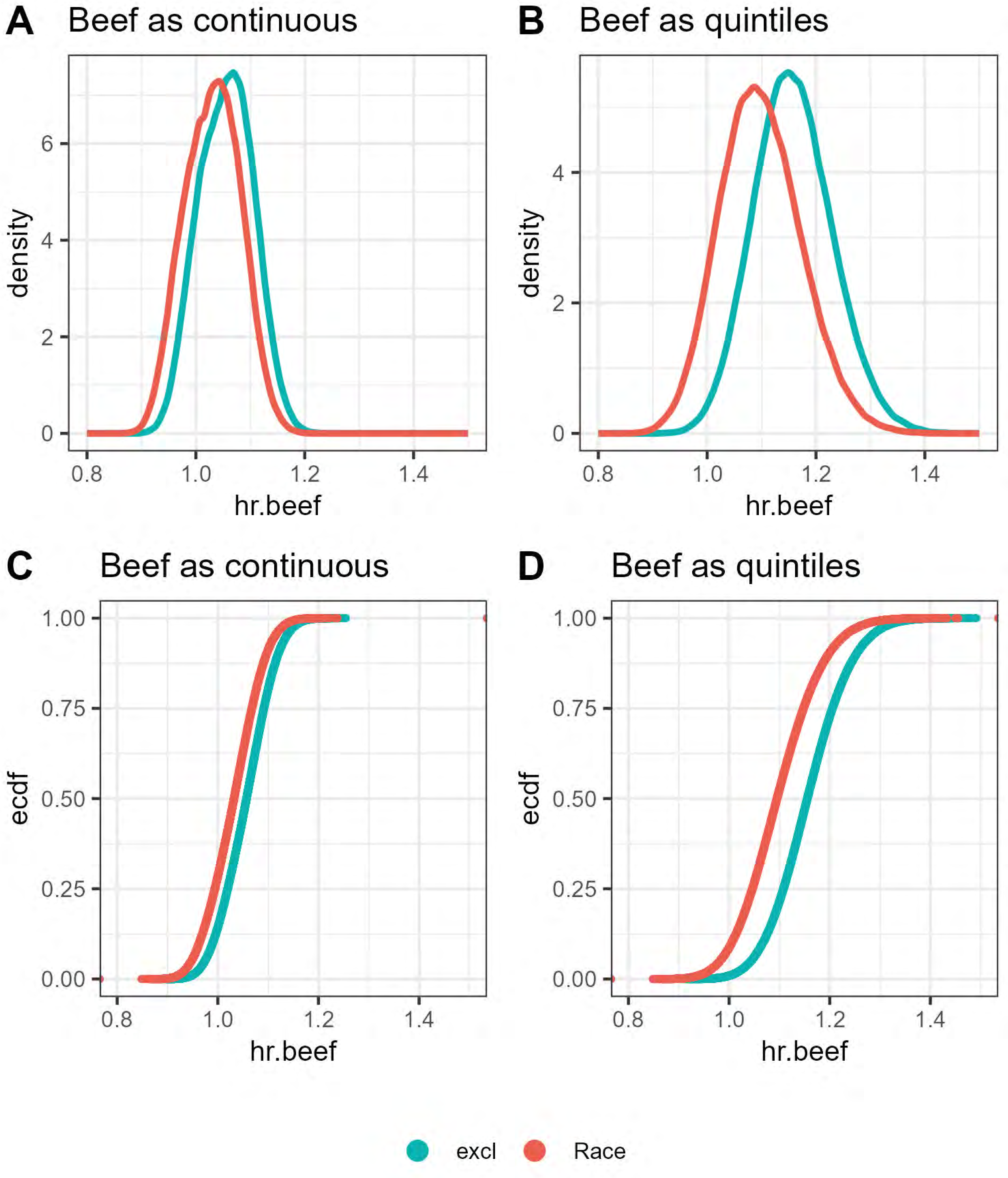
A
Beef as continuous
density
hr.beef
B
Beef as quintiles
density
hr.beef
C
Beef as continuous
ecdf
hr.beef
D
Beef as quintiles
ecdf
hr.beef
excl
Race

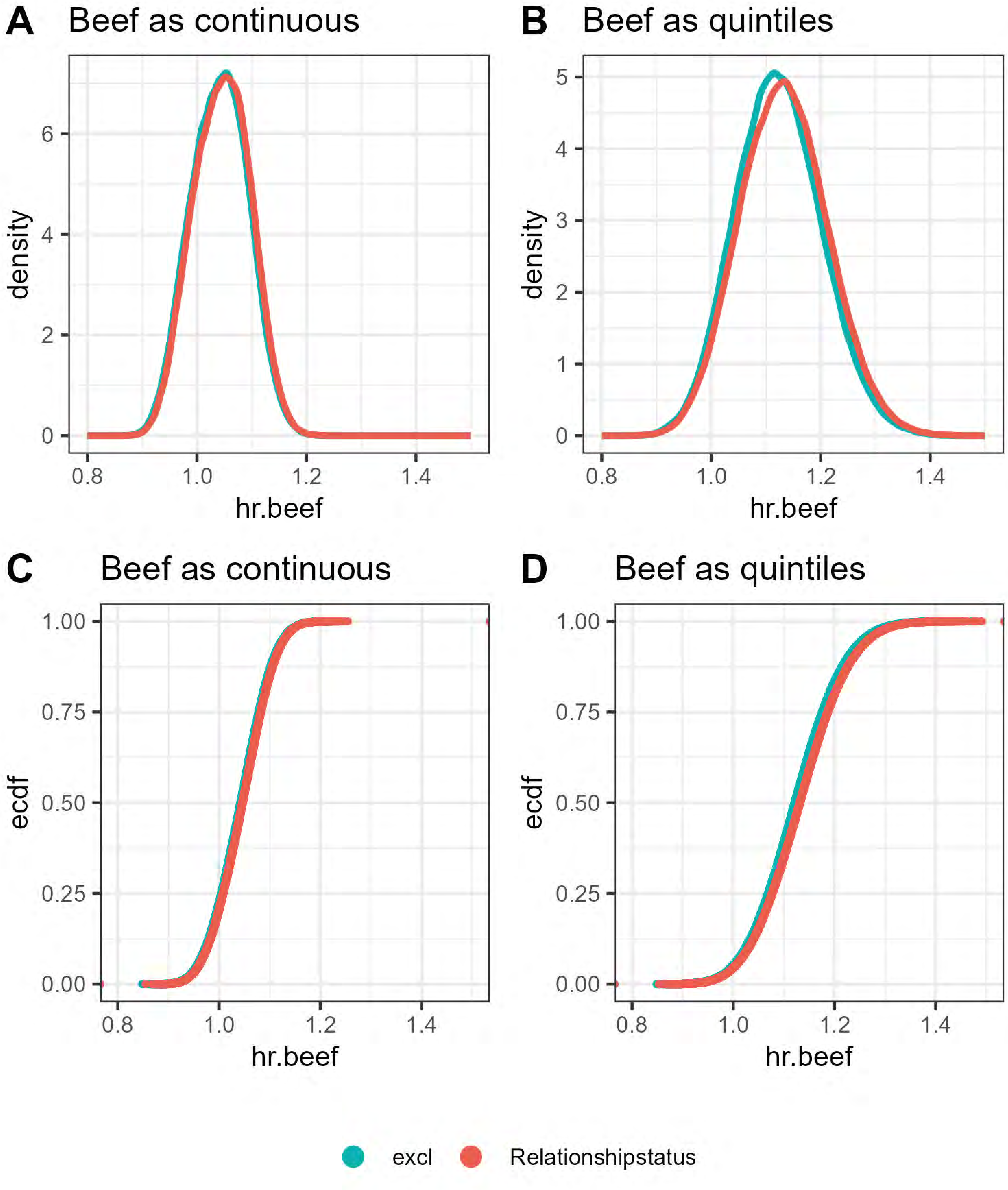

A
Beef as continuous
density
hr.beef
B
Beef as quintiles
density
hr.beef
C
Beef as continuous
ecdf
hr.beef
D
Beef as quintiles
ecdf
hr.beef
excl
Relationshipstatus

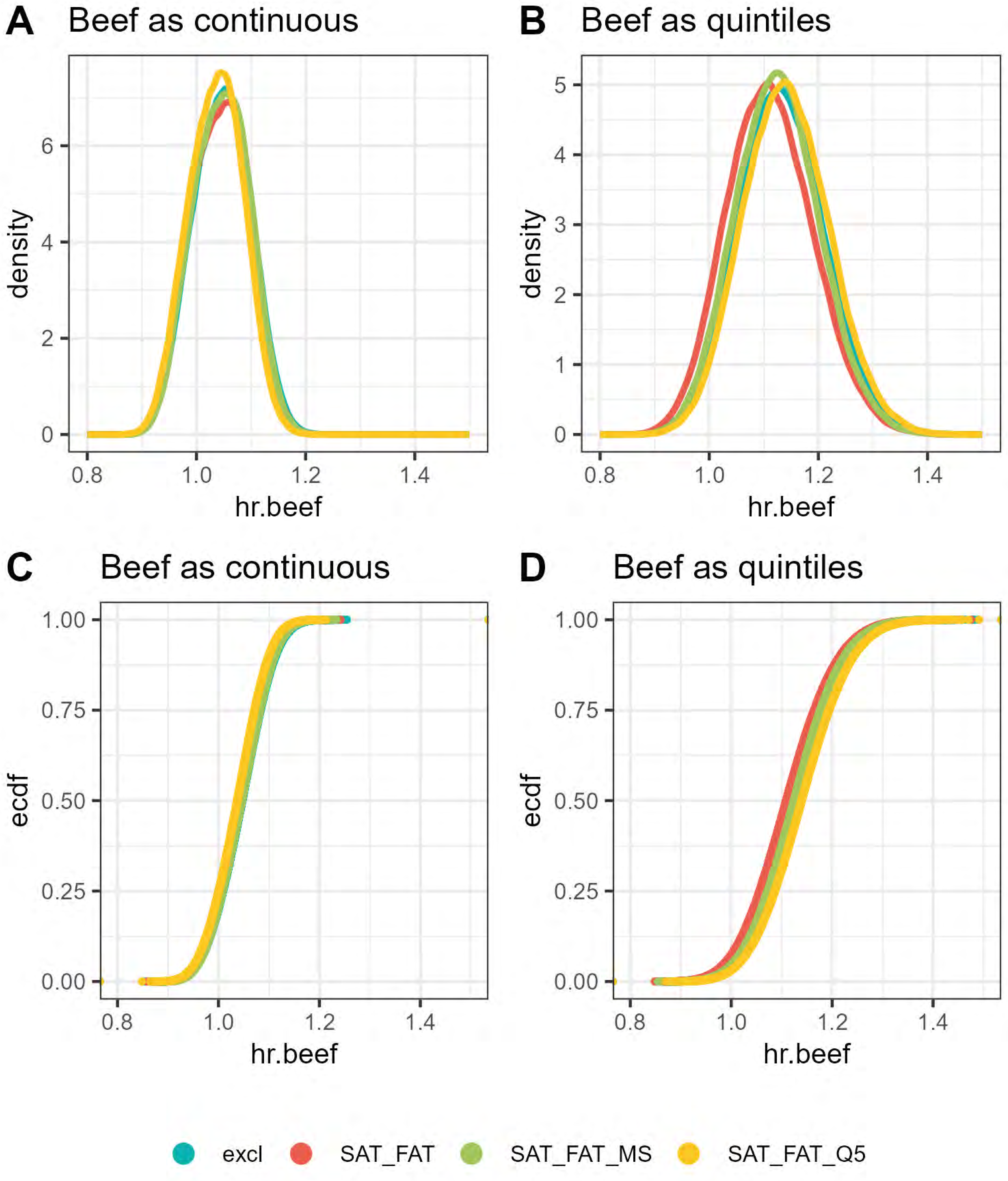

A
Beef as continuous
density
hr.beef
B
Beef as quintiles
density
hr.beef
C
Beef as continuous
ecdf
hr.beef
D
Beef as quintiles
ecdf
hr.beef
excl
SAT_FAT
SAT_FAT_MS
SAT_FAT_Q5

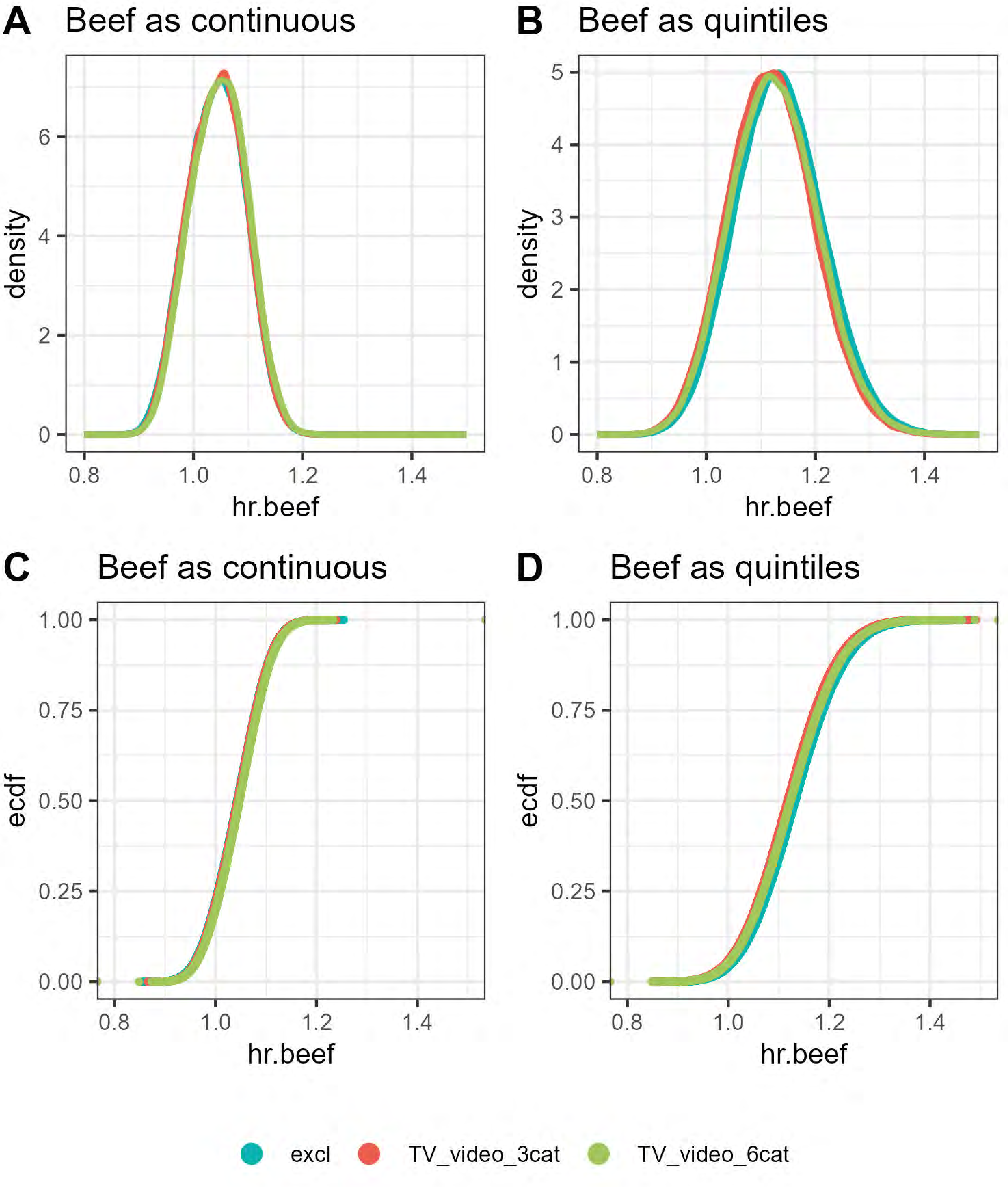
A
Beef as continuous
density
hr.beef
B
Beef as quintiles
density
hr.beef
C
Beef as continuous
ecdf
hr.beef
D
Beef as quintiles
ecdf
hr.beef
excl
TV_video_3cat
TV_video_6cat

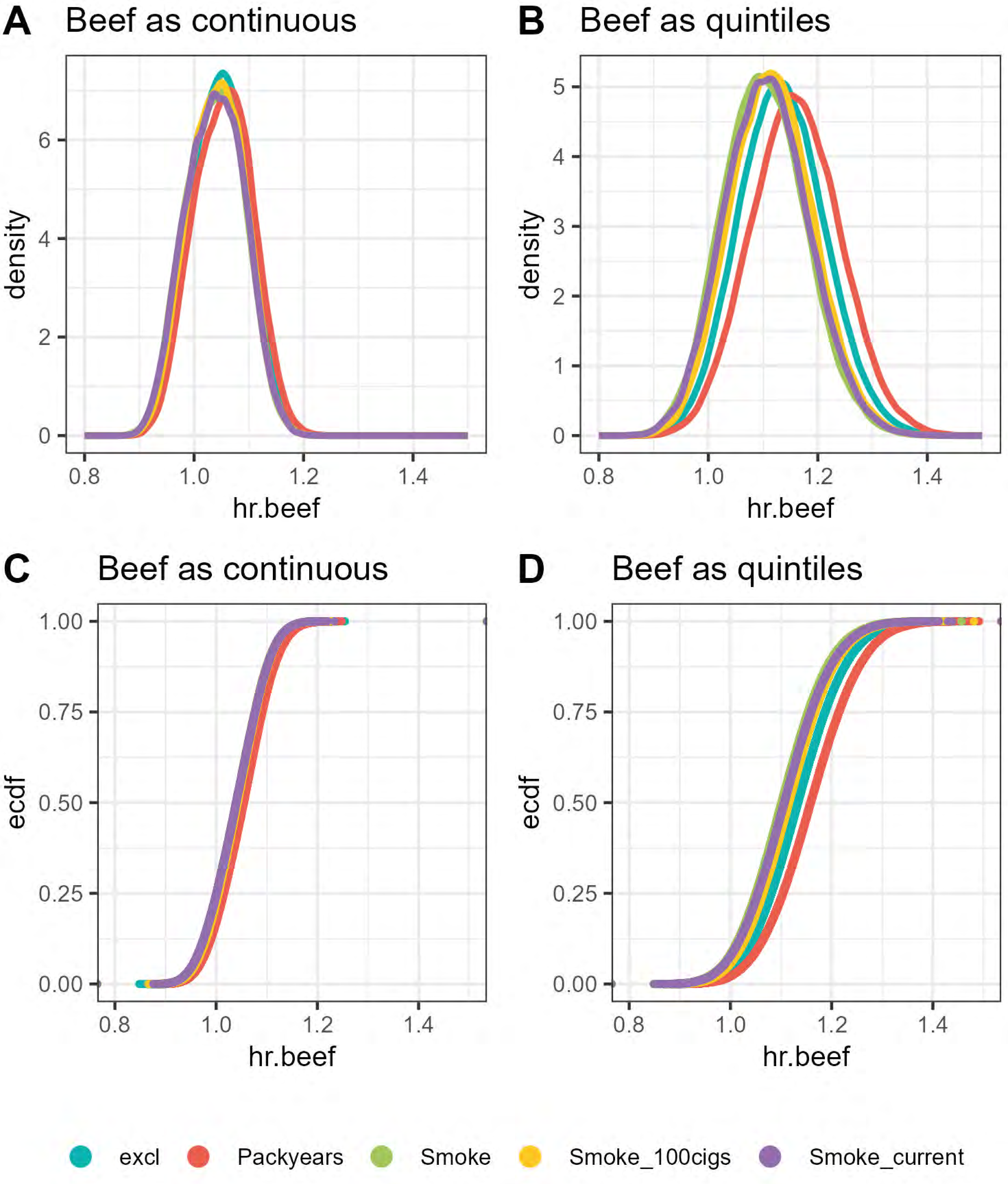

A
Beef as continuous
density
hr.beef
B
Beef as quintiles
density
hr.beef
C
Beef as continuous
ecdf
hr.beef
D
Beef as quintiles
ecdf
hr.beef
excl
Packyears
Smoke
Smoke_100cigs
Smoke_current

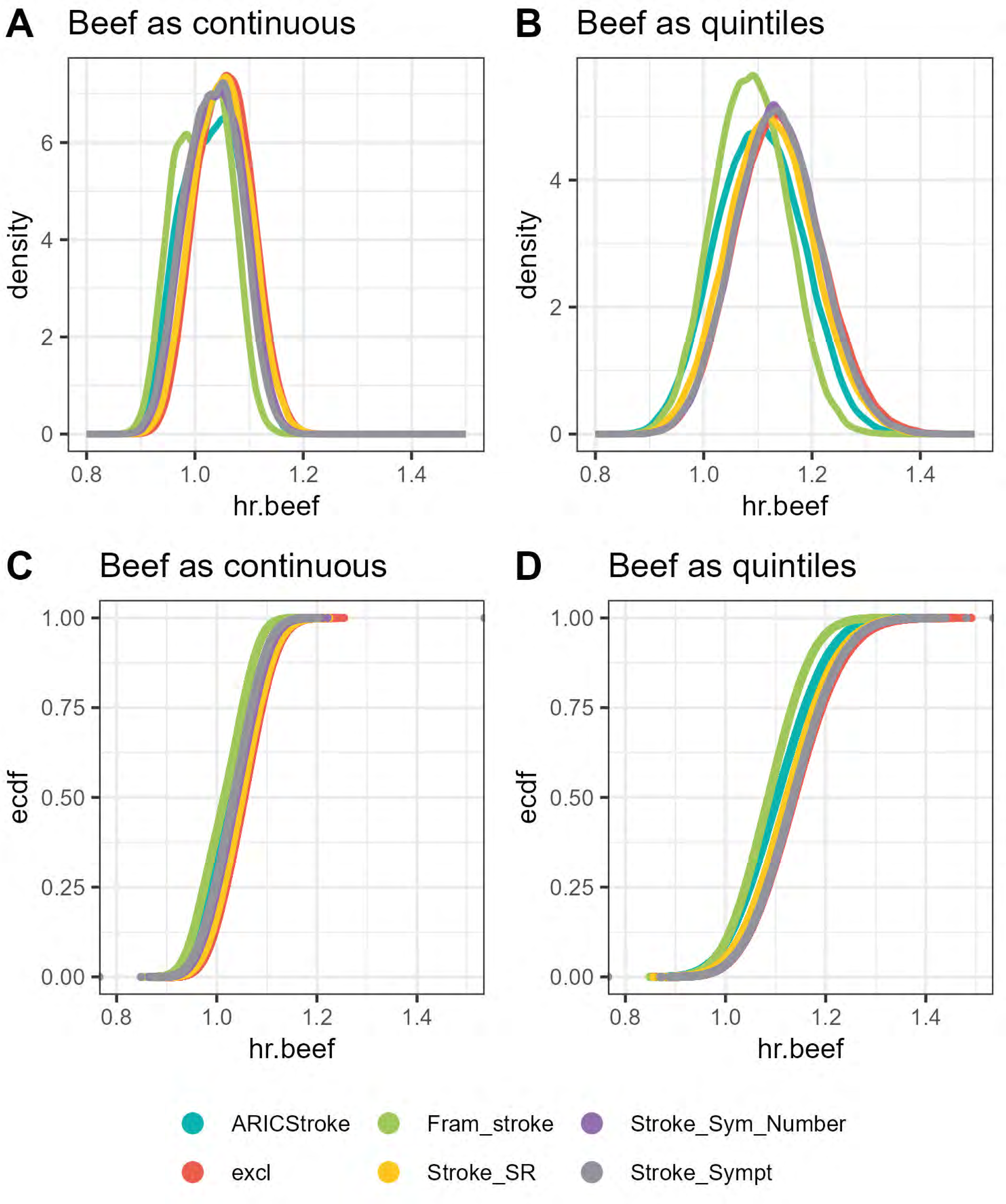

A
Beef as continuous
B
Beef as quintiles
density
hr.beef
C
Beef as continuous
D
Beef as quintiles
ecdf
hr.beef
0.8
1.0
1.2
1.4
0
2
4
6
0.00
0.25
0.50
0.75
1.00
ARICStroke
Fram_stroke
Stroke_Sym_Number
excl
Stroke_SR
Stroke_Sympt

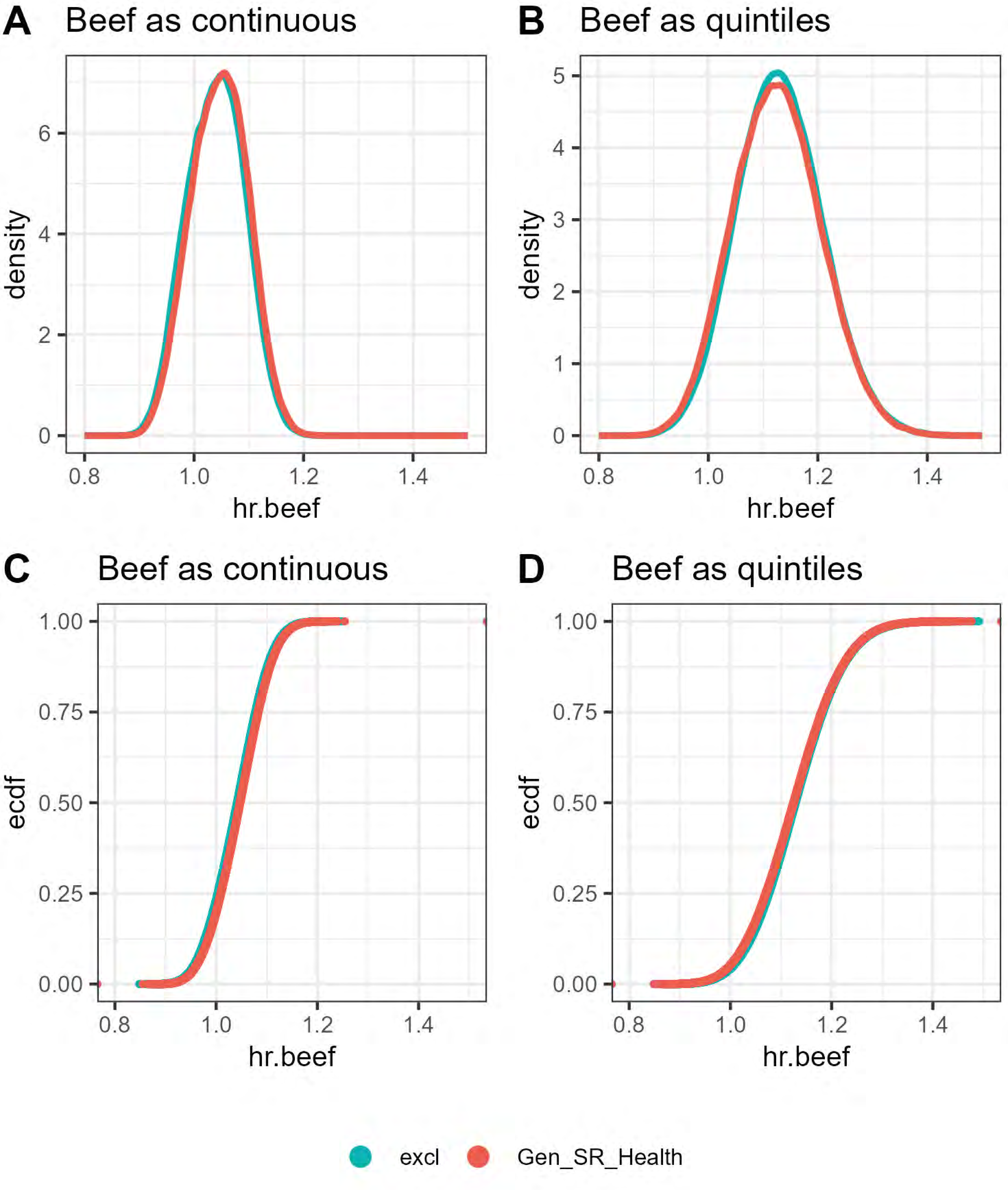

A
Beef as continuous
density
hr.beef
0
2
4
6
0.8
1.0
1.2
1.4
B
Beef as quintiles
density
hr.beef
0
1
2
3
4
5
C
Beef as continuous
ecdf
hr.beef
0.00
0.25
0.50
0.75
1.00
D
Beef as quintiles
ecdf
hr.beef
excl
Gen_SR_Health

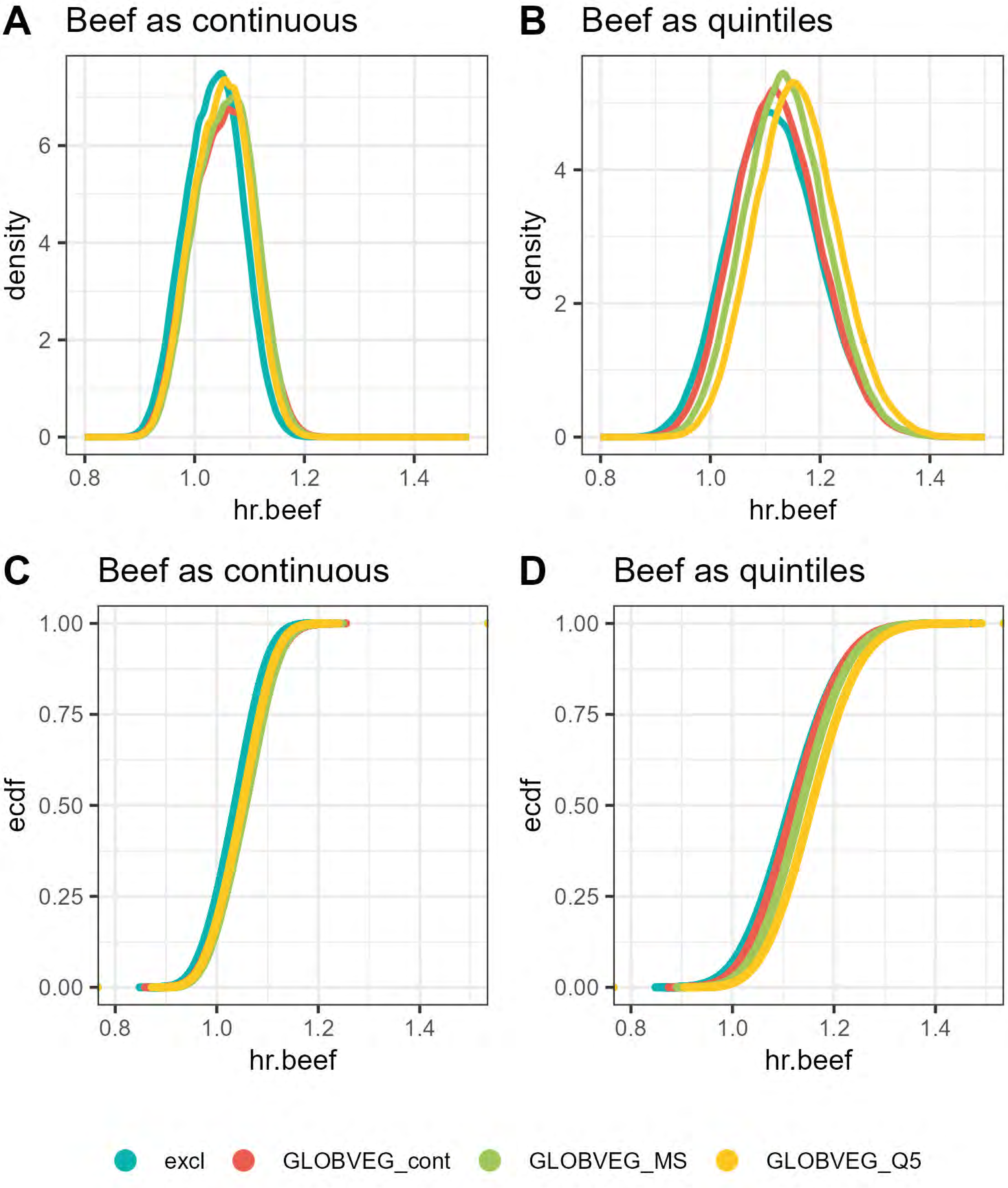
A
Beef as continuous
B
Beef as quintiles
C
Beef as continuous
D
Beef as quintiles
density
ecdf
hr.beef
0
2
4
6
0.8
1.0
1.2
1.4
0.00
0.25
0.50
0.75
1.00
excl
GLOBVEG_cont
GLOBVEG_MS
GLOBVEG_Q5

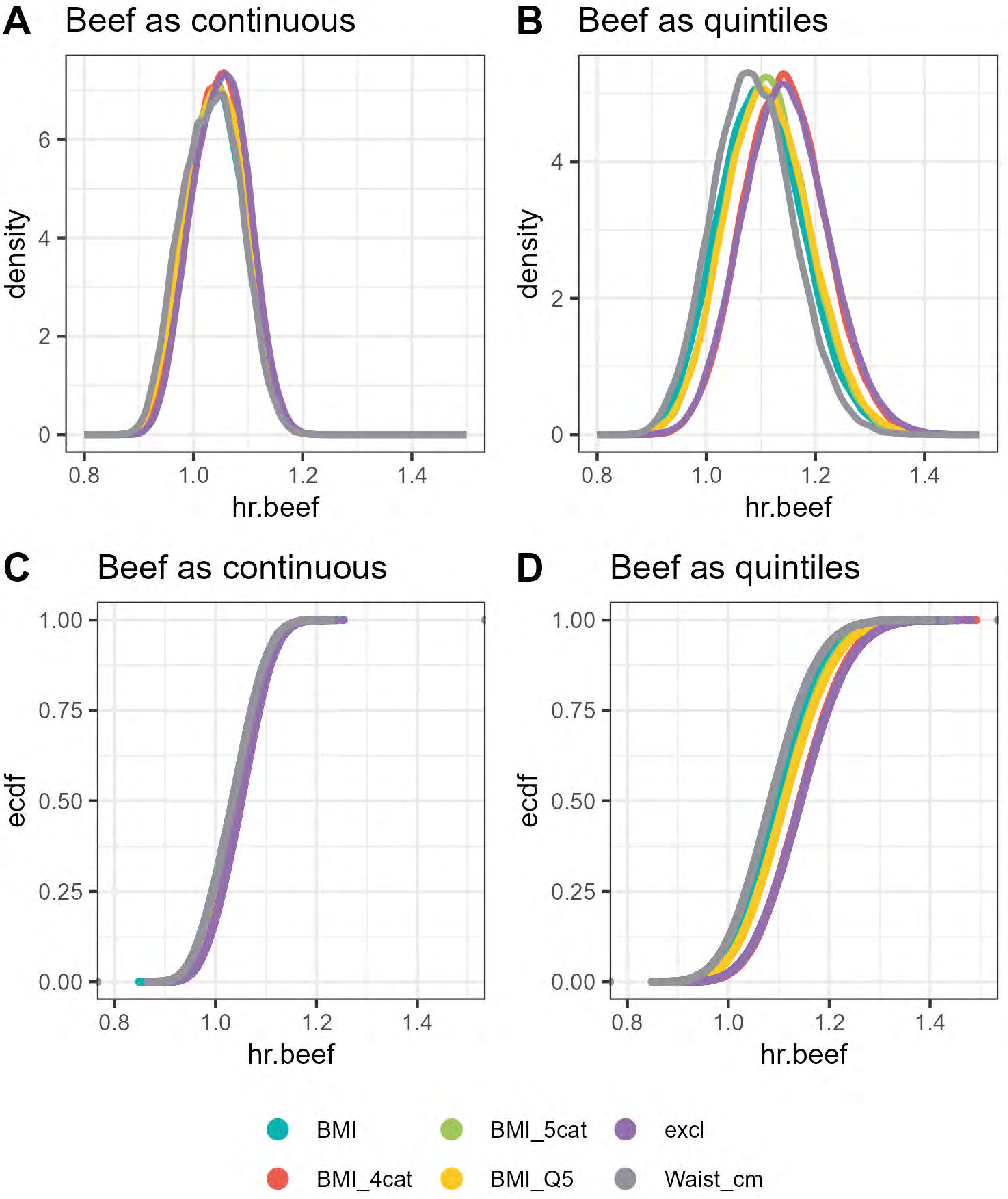
A
Beef as continuous
B
Beef as quintiles
C
Beef as continuous
D
Beef as quintiles
density
ecdf
hr.beef
BMI
BMI_4cat
BMI_5cat
BMI_Q5
excl
Waist_cm

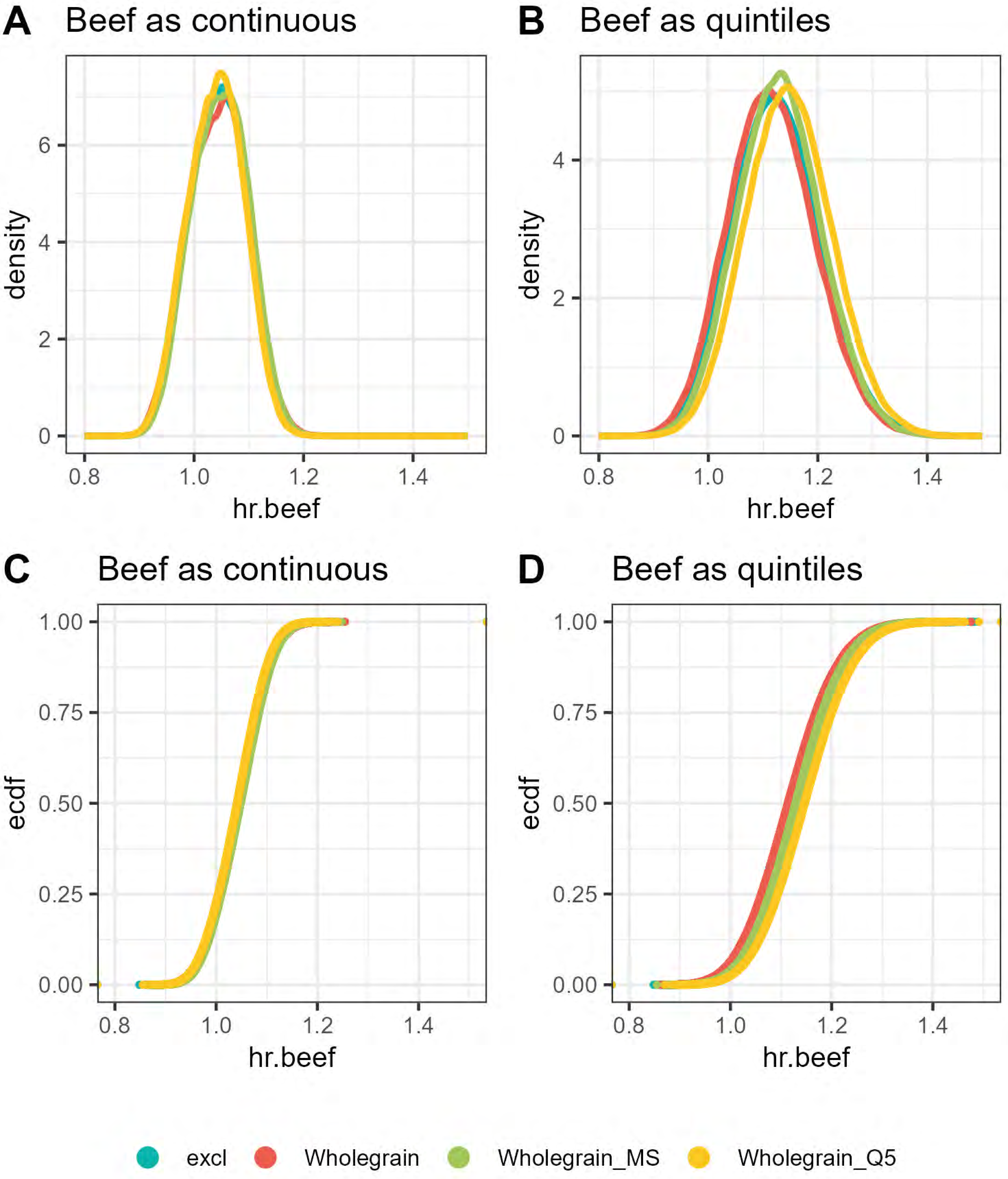
A
Beef as continuous
B
Beef as quintiles
C
Beef as continuous
D
Beef as quintiles
density
ecdf
hr.beef
0
2
4
6
0.00
0.25
0.50
0.75
1.00
0.8
1.0
1.2
1.4
excl
Wholegrain
Wholegrain_MS
Wholegrain_Q5
