## Supplemental File 3 for "‘Shaking the Ladder’ reveals how analytic choices can influence associations in nutrition epidemiology: beef intake and coronary heart disease as a case study"

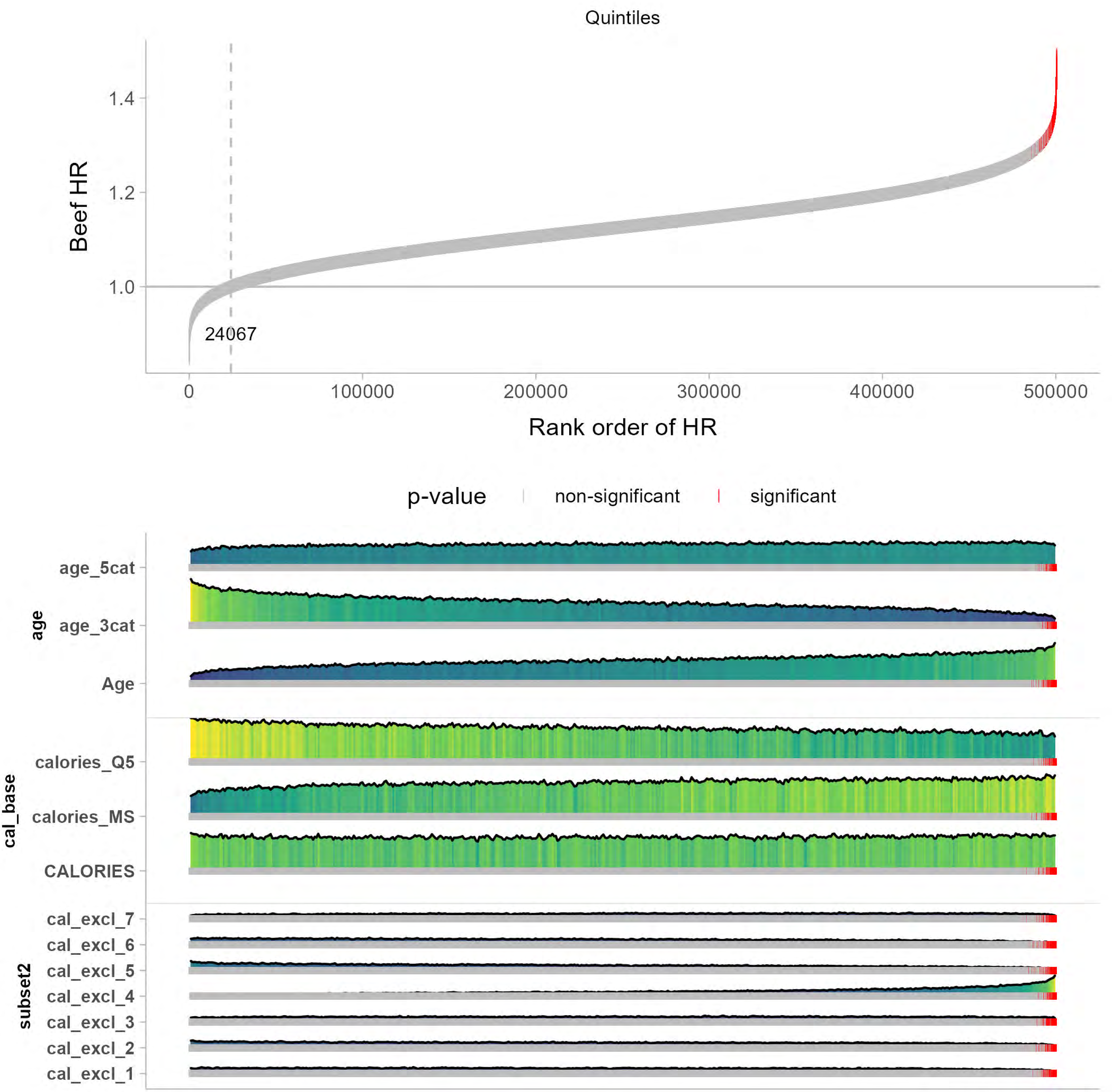

Quintiles
Beef HR
1.4
1.2
1.0
24067
0
100000
200000
300000
400000
500000
Rank order of HR
p-value
non-significant
significant
age
age_5cat
age_3cat
Age
cal_base
calories_Q5
calories_MS
CALORIES
subset2
cal_excl_7
cal_excl_6
cal_excl_5
cal_excl_4
cal_excl_3
cal_excl_2
cal_excl_1

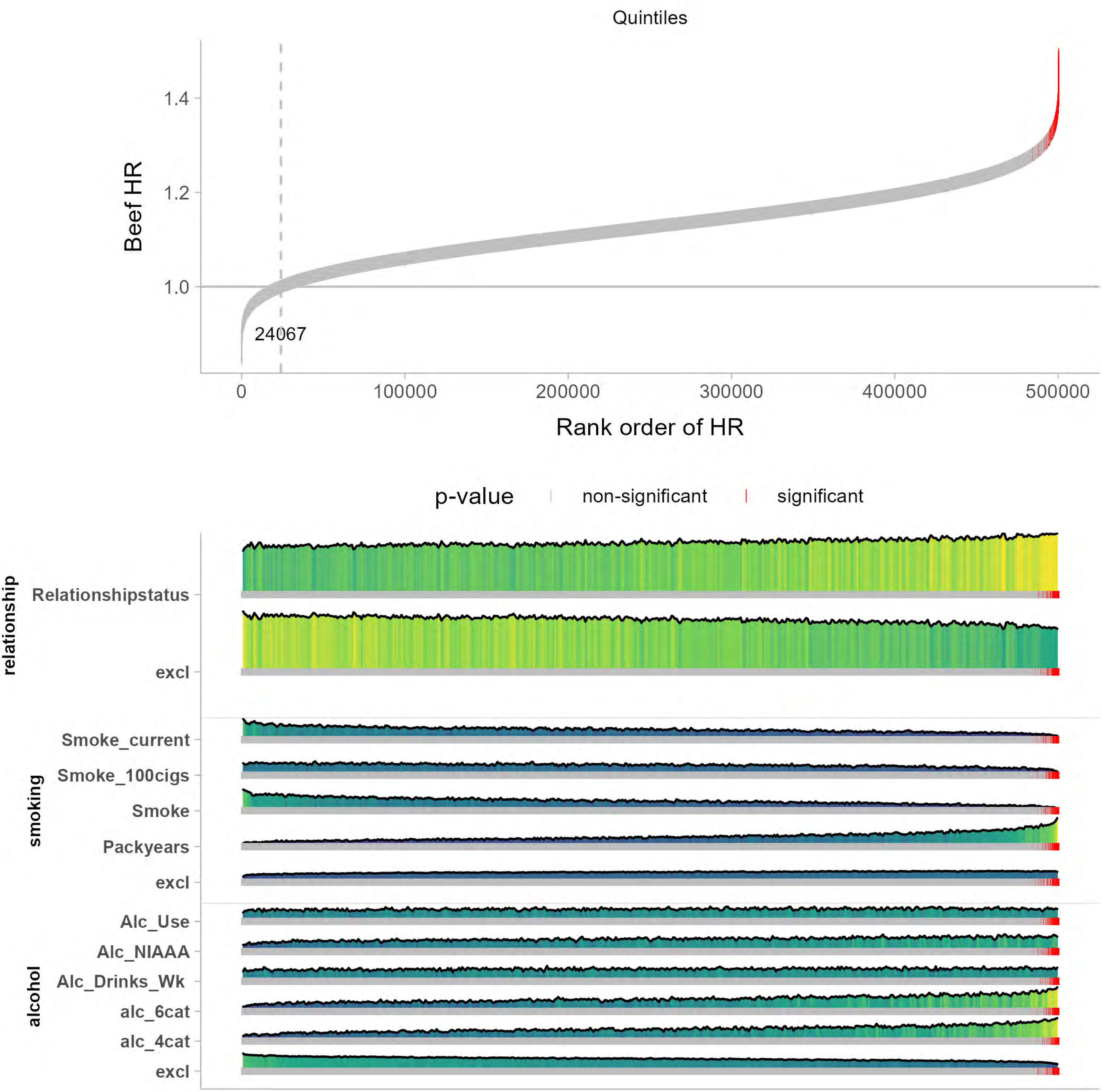

Quintiles
Beef HR
1.4
1.2
1.0
24067
0
100000
200000
300000
400000
500000
Rank order of HR
p-value
non-significant
significant
relationship
Relationshipstatus
excl
smoking
Smoke_current
Smoke_100cigs
Smoke
Packyears
excl
alcohol
Alc_Use
Alc_NIAAA
Alc_Drinks_Wk
alc_6cat
alc_4cat
excl

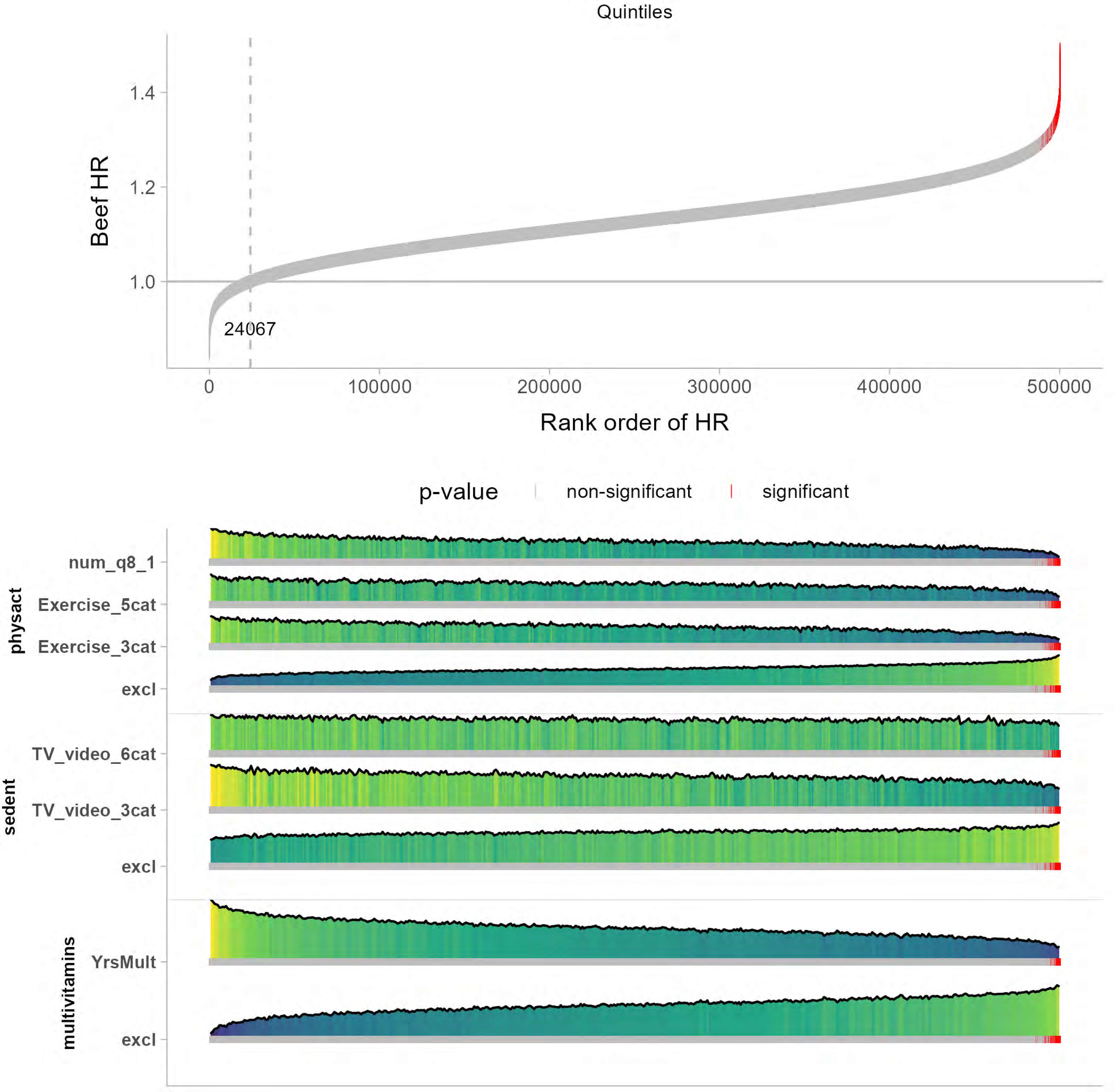

Quintiles
Beef HR
1.4
1.2
1.0
24067
0
100000
200000
300000
400000
500000
Rank order of HR
p-value
non-significant
significant
physact
num_q8_1
Exercise_5cat
Exercise_3cat
excl
sedent
TV_video_6cat
TV_video_3cat
excl
multivitamins
YrsMult
excl

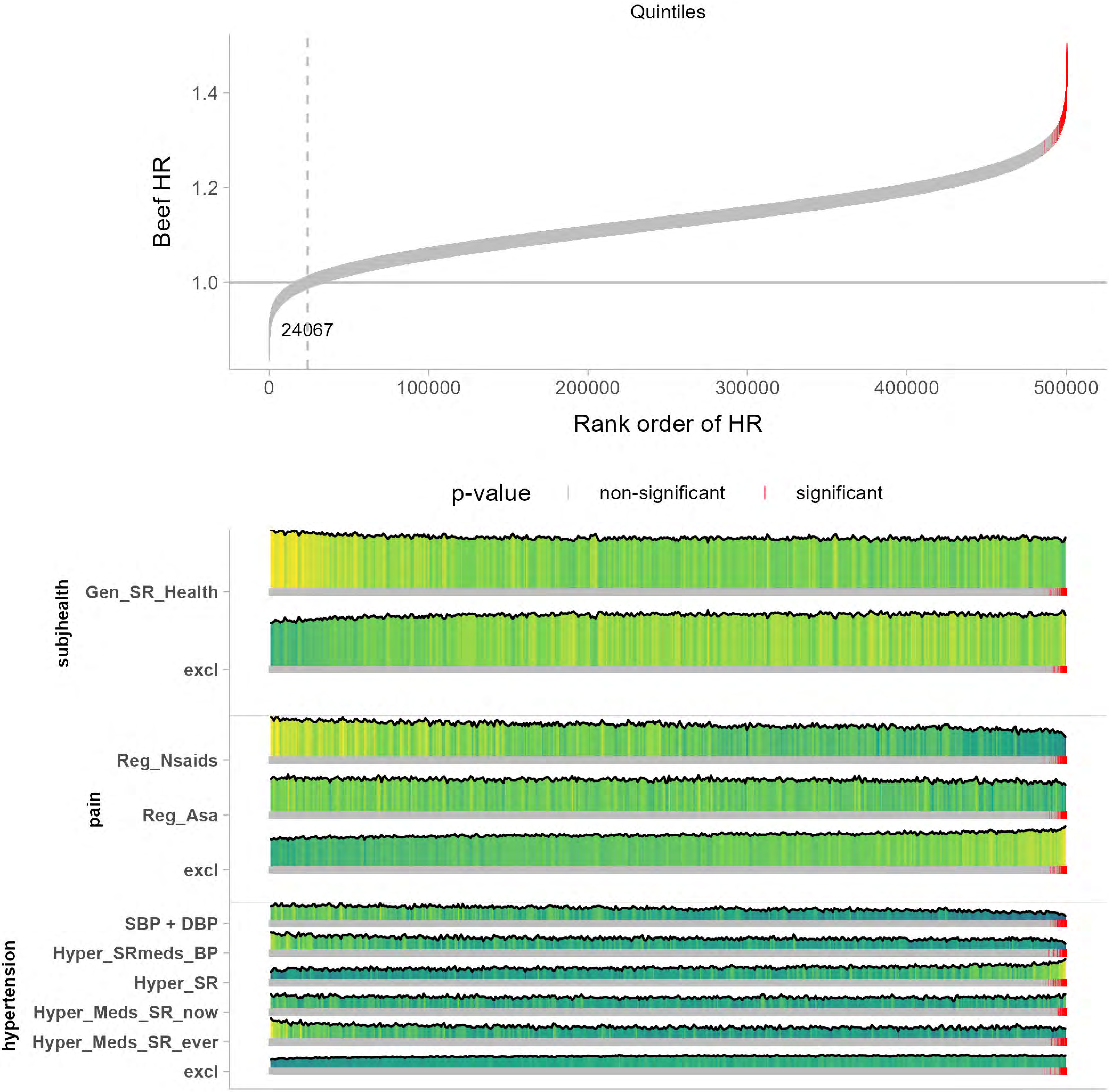
Quintiles
Beef HR
1.4
1.2
1.0
24067
0
100000
200000
300000
400000
500000
Rank order of HR
p-value
non-significant
significant
subjhealth
Gen_SR_Health
excl
pain
Reg_Nsaids
Reg_Asa
excl
hypertension
SBP + DBP
Hyper_SRmeds_BP
Hyper_SR
Hyper_Meds_SR_now
Hyper_Meds_SR_ever
excl

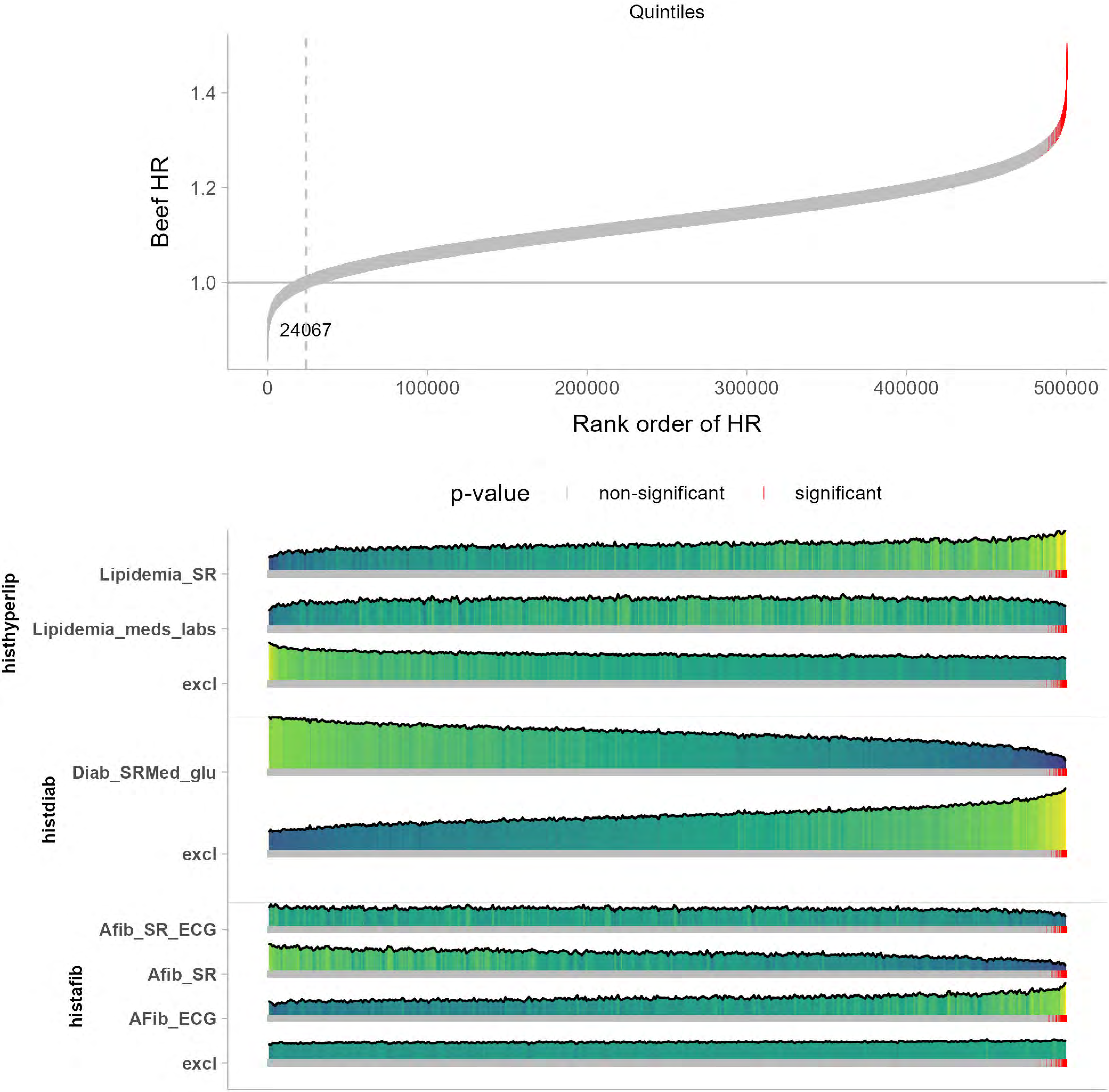

Quintiles
Beef HR
1.4
1.2
1.0
24067
0
100000
200000
300000
400000
500000
Rank order of HR
p-value
non-significant
significant
histhyperlip
Lipidemia_SR
Lipidemia_meds_labs
excl
histdiab
Diab_SRMed_glu
excl
histafib
Afib_SR_ECG
Afib_SR
AFib_ECG
excl

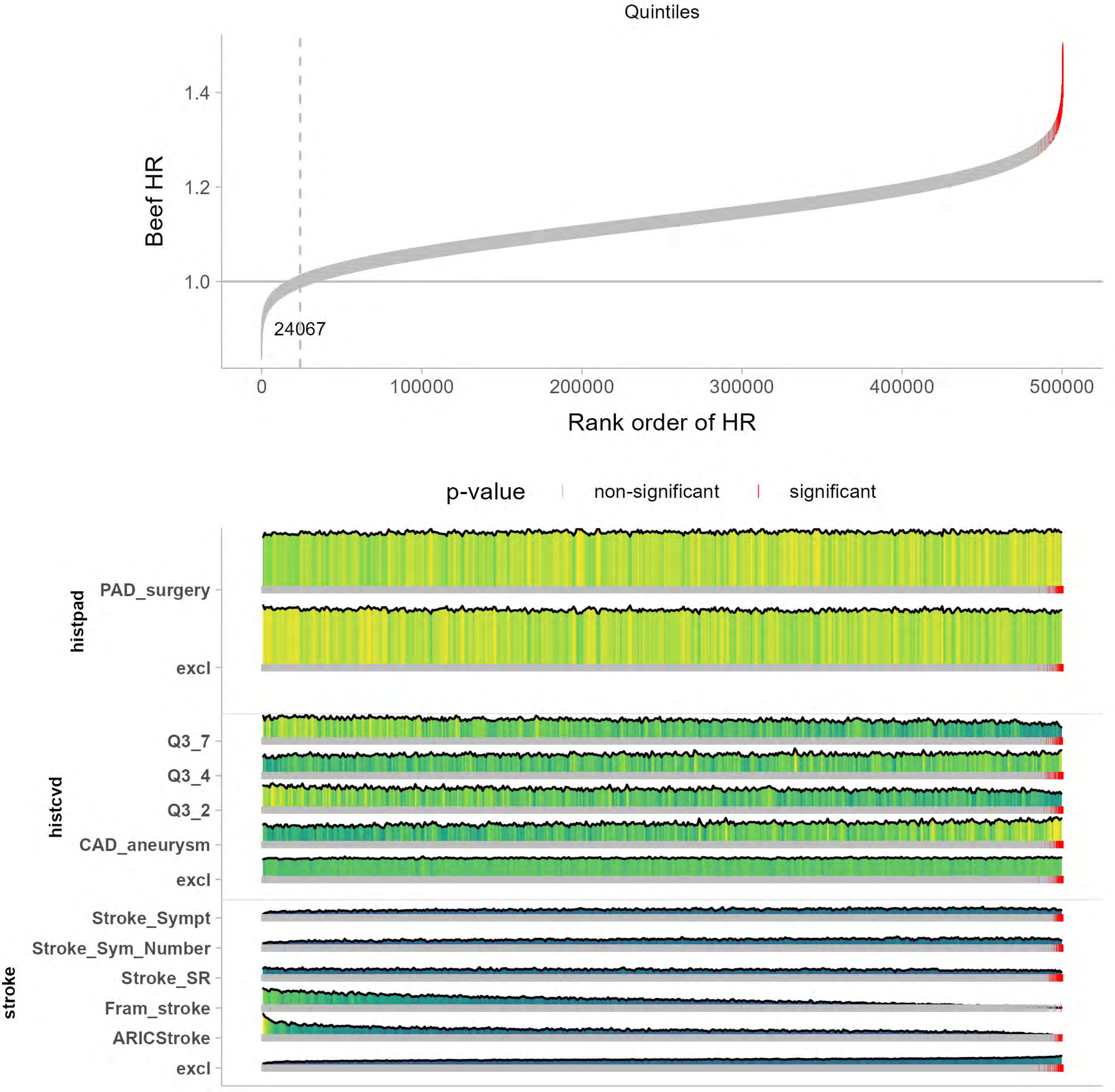

Quintiles
Beef HR
1.4
1.2
1.0
24067
0
100000
200000
300000
400000
500000
Rank order of HR
p-value
non-significant
significant
histpad
PAD_surgery
excl
histcvd
Q3_7
Q3_4
Q3_2
CAD_aneurysm
excl
stroke
Stroke_Sympt
Stroke_Sym_Number
Stroke_SR
Fram_stroke
ARICStroke
excl

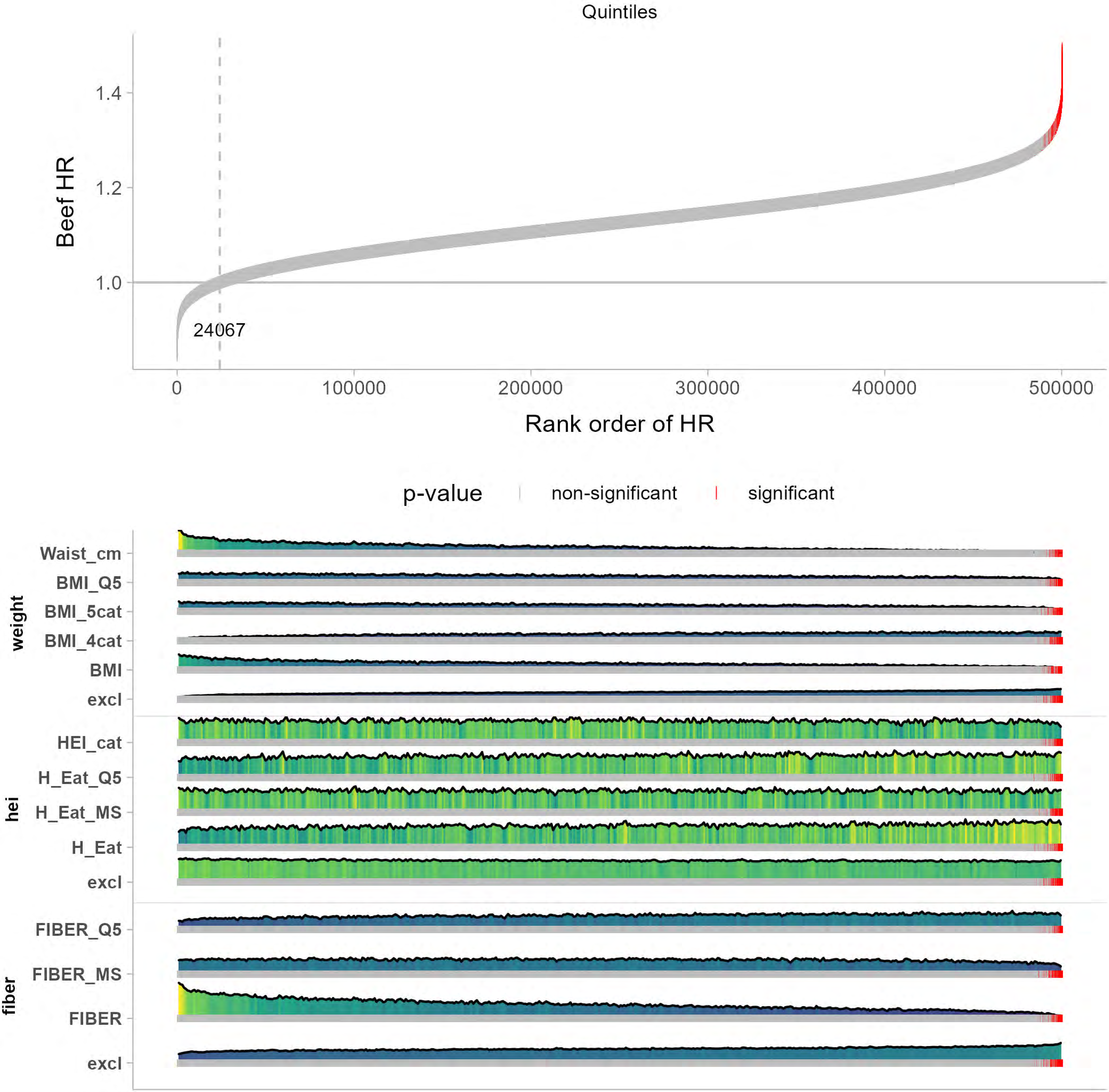
Quintiles
Beef HR
1.4
1.2
1.0
24067
0
100000
200000
300000
400000
500000
Rank order of HR
p-value
non-significant
significant
weight
Waist_cm
BMI_Q5
BMI_5cat
BMI_4cat
BMI
excl
hei
HEI_cat
H_Eat_Q5
H_Eat_MS
H_Eat
excl
fiber
FIBER_Q5
FIBER_MS
FIBER
excl

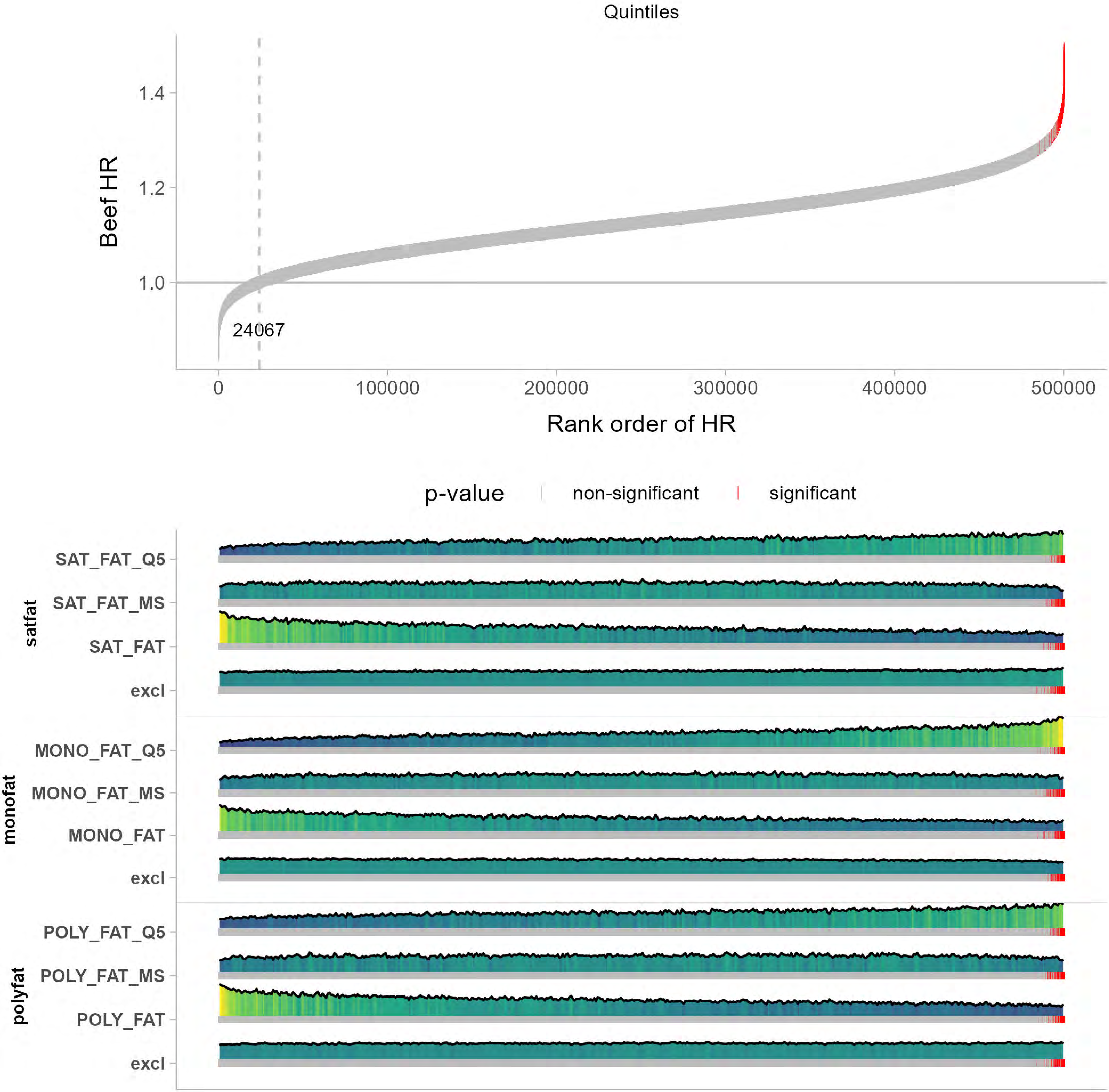
Quintiles
Beef HR
1.4
1.2
1.0
24067
0
100000
200000
300000
400000
500000
Rank order of HR
p-value
non-significant
significant
satfat
SAT_FAT_Q5
SAT_FAT_MS
SAT_FAT
excl
monofat
MONO_FAT_Q5
MONO_FAT_MS
MONO_FAT
excl
polyfat
POLY_FAT_Q5
POLY_FAT_MS
POLY_FAT
excl

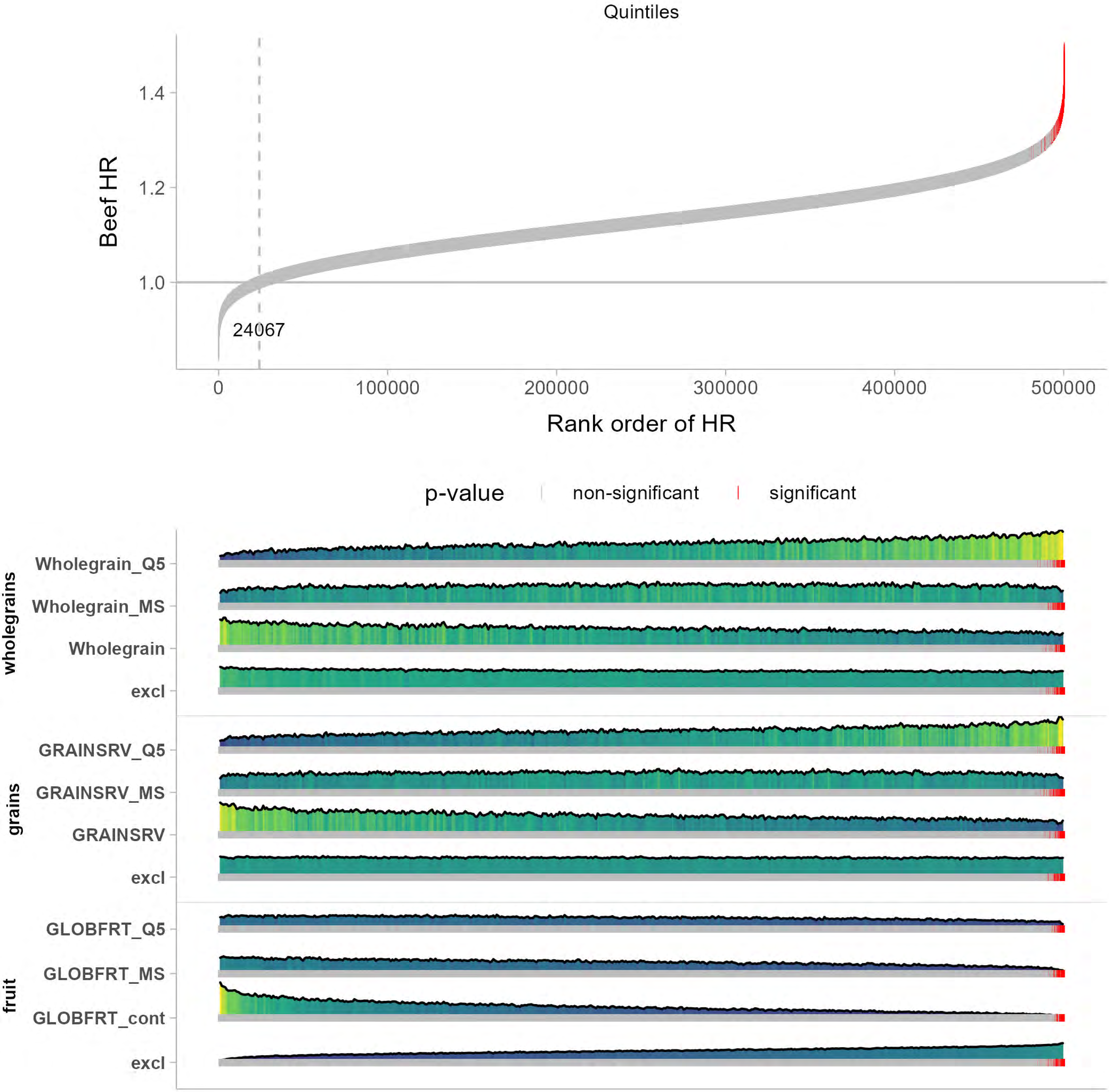
Quintiles
Beef HR
1.4
1.2
1.0
24067
0
100000
200000
300000
400000
500000
Rank order of HR
p-value
non-significant
significant
wholegrains
Wholegrain_Q5
Wholegrain_MS
Wholegrain
excl
grains
GRAINSRV_Q5
GRAINSRV_MS
GRAINSRV
excl
fruit
GLOBFRT_Q5
GLOBFRT_MS
GLOBFRT_cont
excl

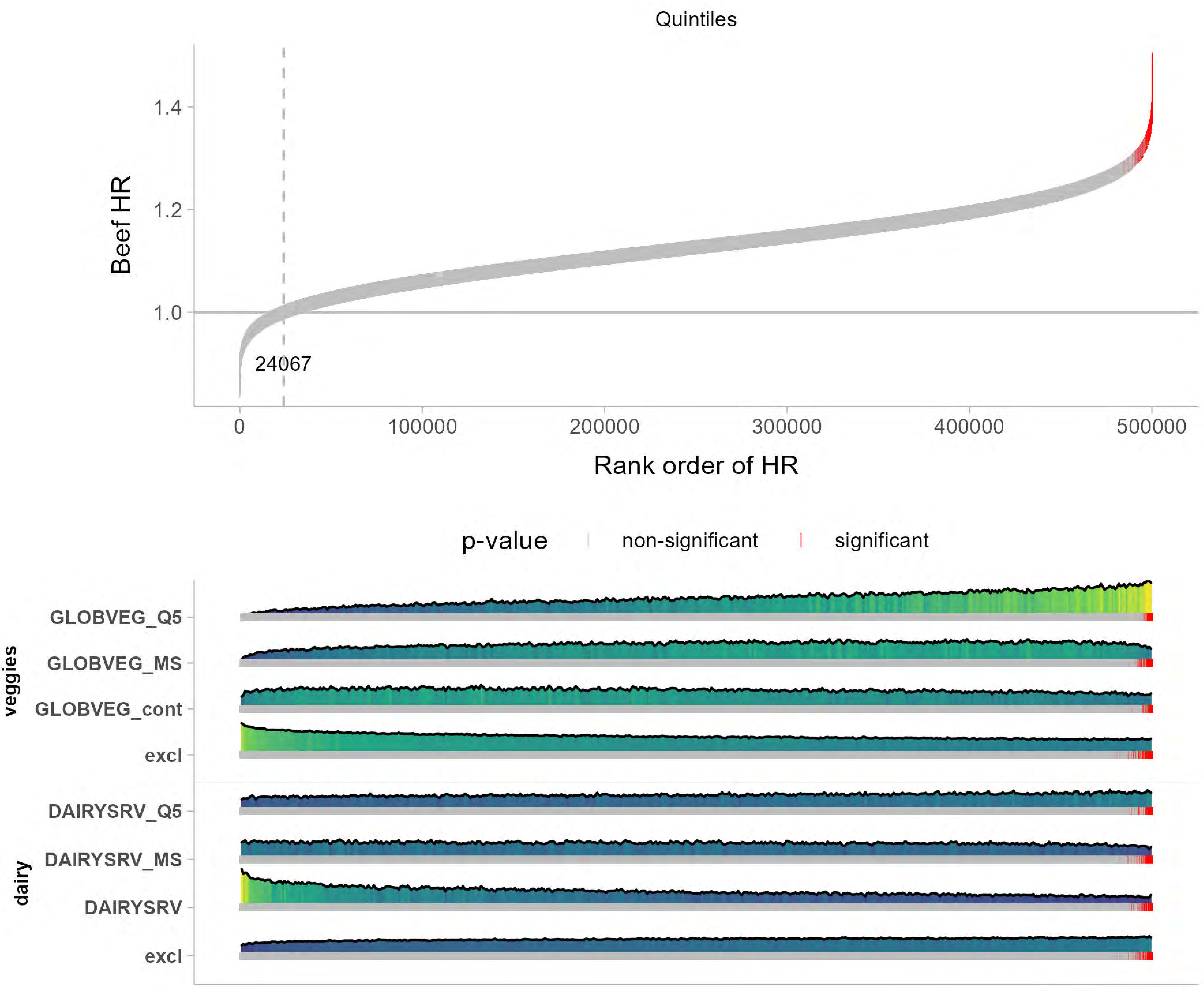

Quintiles
Beef HR
1.4
1.2
1.0
24067
0
100000
200000
300000
400000
500000
Rank order of HR
p-value
non-significant
significant
veggies
GLOBVEG_Q5
GLOBVEG_MS
GLOBVEG_cont
excl
dairy
DAIRYSRV_Q5
DAIRYSRV_MS
DAIRYSRV
excl

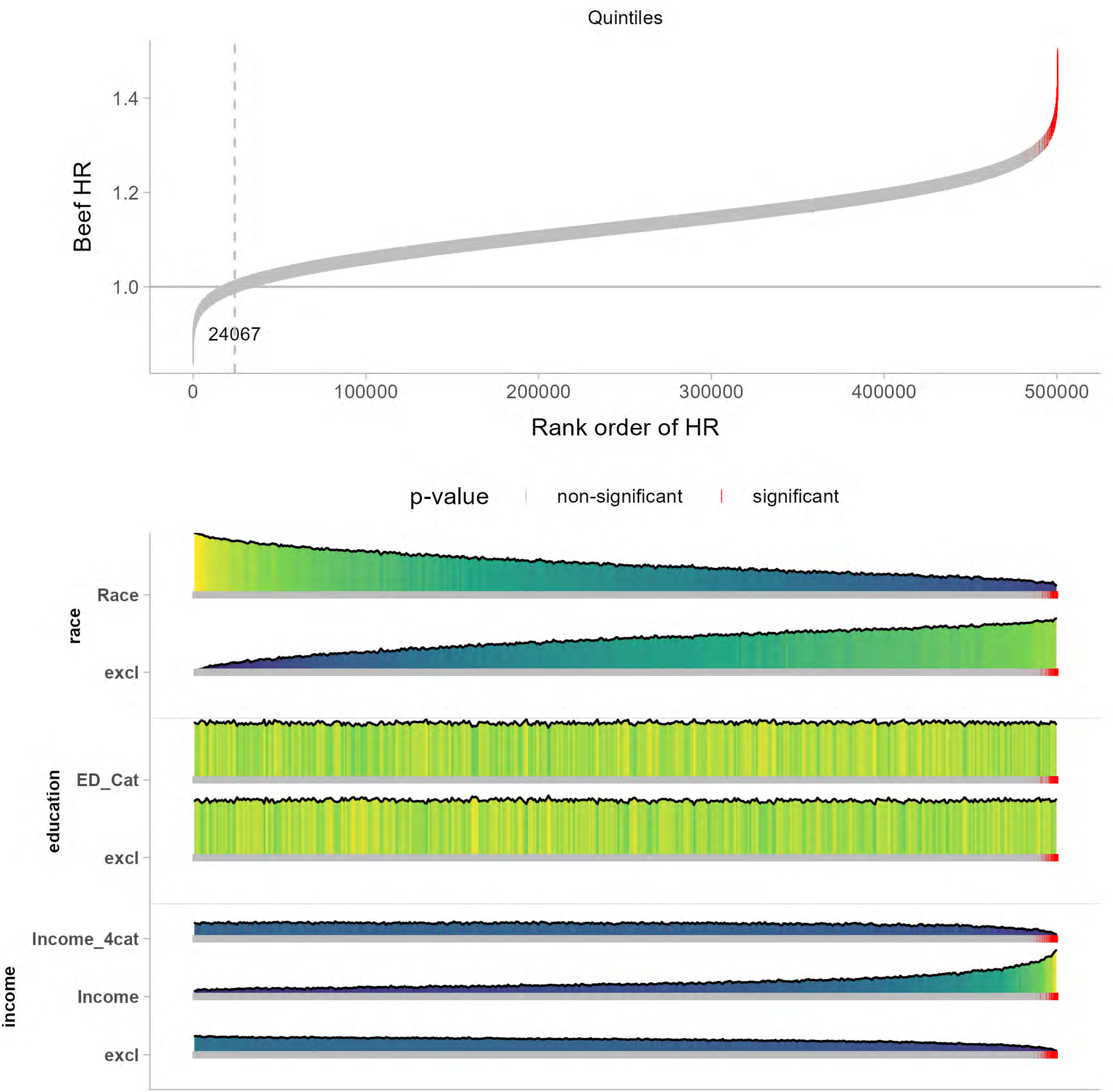

Quintiles
Beef HR
1.4
1.2
1.0
24067
0
100000
200000
300000
400000
500000
Rank order of HR
p-value
non-significant
significant
race
Race
excl
education
ED_Cat
excl
income
Income_4cat
Income
excl
